## Supplemental Appendix for "Cost-effectiveness of leveraging existing HIV primary health systems and community health workers for hypertension screening and treatment in Africa: an individual-based modelling study"

### HIV Synthesis Hypertension Supplemental Appendix

### Section 1. HIV Synthesis Hypertension Model Extended Methods

The HIV Synthesis model is an individual-based simulation model of HIV transmission, treatment, morbidity, and mortality in east, southern, central, and west Africa.[1–3] For the present analysis, we added modeling parameters for systolic blood pressure (SBP), hypertension diagnosis and treatment, and cardiovascular disease (CVD) morbidity and mortality to the HIV Synthesis model. We limit our model to SBP, rather than separately modeling diastolic blood pressure (DBP), due to the greater effect of SBP on CVD outcomes,[4] particularly with increased age.[5] In the model, hypertension is therefore diagnosed when measured SBP (incorporating measurement error from true SBP) is ≥140 mmHg on two consecutive clinic visits or ≥160 mmHg on a single visit. We model changes in SBP over the life course, and the relationship between age, sex, SBP, and HIV on incident CVD events (ischemic heart disease and cerebrovascular accident) and subsequent morbidity and mortality. Each model run generates a simulated population of adults from 1989 to 2023 and beyond, with attributes updated every 3-months. Hypertension-specific attributes updated each 3-month period include age, true underlying SBP, blood pressure measurement in the community, occurrence of a hypertension clinic visit, measured SBP, initiation of antihypertensive therapy, initiation of a 2$^{nd}$ or 3$^{rd}$ antihypertensive drug, number of antihypertensive drugs, incident ischemic heart disease, incident cerebrovascular accident, and death (details in Table S6). Individuals in the model also have an individual starting SBP at age 15, individual relative risk of SBP rise over time, and SBP reduction associated with each of three sequential hypertension medications should they be taken (Table S5). We incorporate adherence to antihypertensive medications by modeling the probability that an individual will return to the clinic for a repeat hypertension care visit; those who do not return are assumed to discontinue medications.

We generated 3000 setting-scenarios to represent a range of settings across east, southern, central, and west Africa and to incorporate uncertainty in model assumptions (Table S4). At the setting level, parameters included base probability of SBP rise per 3-month period, rates of blood pressure measurement at a clinic visit (both for those without a prior hypertension diagnosis and with a prior hypertension diagnosis), and relative rates of care-seeking for blood pressure measurement in women compared to men and younger compared to older adults. We also sample the probability of treatment initiation based on measured blood pressure and the setting-level probability of returning for follow-up. We assume a treatment target SBP of <140 mmHg.[6] We show comparable national estimates for hypertension prevalence, diagnosis, treatment, and blood pressure control where available, though we intend for setting-scenarios to represent sub-settings within countries as much as countries as a whole. Table 1 in the main text and Section 4 of the appendix show model outputs compared to data from the literature.

We modeled CVD events (ischemic heart disease and cerebrovascular accident) using risk equations incorporating age, SBP, HIV status, history of CVD, and setting-scenario variability in baseline CVD risk. For those with HIV, we additionally incorporate CD4 and HIV viral load into risk equations.[7,8] We estimate the relationship between SBP lowering down to a threshold of 115 mmHg and CVD risk based on meta-analyses of randomized controlled trials of antihypertensive medications, incorporating interaction with age.[9–11] For incident CVD events, we model a distribution of 'mild', 'moderate', and 'severe' events with an associated gradation in both acute and long-term mortality risk. Similarly, we include disability weights associated with ischemic heart disease and cerebrovascular accident based on disease severity (Table S7). We assume that most mild events are under-ascertained in available CVD incidence data from the literature and therefore limited to incident moderate and severe ischemic heart disease and cerebrovascular disease events to calibrate model output against available CVD incidence data from the literature.

Once an ischemic heart disease or cerebrovascular disease event occurs, we assume both an acute (during that 3-month period) and long-term mortality risk according to the severity of the event. Where possible, we model mortality risk based on CVD disease severity using real-world data from east, southern, central, and west Africa. We additionally modeled utilization of emergency medical care for incident CVD events, assuming that only those with moderate or severe events would seek care. We modeled setting-scenario variability in the proportion of individuals who seek care following an event and the proportion of care episodes that lead to effective treatment. We assume that ineffective care for acute CVD events may be due to a combination of delayed care seeking or inadequate available treatment at the health facility where care is sought. We model relative risk reduction in acute mortality associated with effective care based on trial data for medical therapies that are expected to be widely available in east, southern, central, and west Africa (e.g. aspirin, supportive care).

For each setting-scenario, we consider the situation in mid-2023 and then compare outcomes from continued standard hypertension care over the subsequent 50 years to three separate policies:

1) current standard of care for hypertension treatment

2) implementation of a clinic-based patient-centered hypertension care model
3) addition of annual community-based hypertension screening by community health workers

We model each of these policies based on data from the SEARCH study.[12,13]

We take a health system perspective for modelling costs. We include costs associated with clinic visits for hypertension care and for each hypertension drug taken (up to three drugs). For intervention scenarios involving community-based screening, we include costs associated with each person screened. We also model costs associated with care for acute CVD events, if care is sought.

We simulate the absolute number of cardiovascular events, costs, and disability adjusted life years (DALYs) for a base population of 10 million adults in 2023 over a 50-year period, with a 3% discount rate for future costs and DALYs averted. We analyzed resource use and cost from a healthcare system perspective. We also calculate net DALYs averted, which is the difference between DALYs averted by an intervention and DALYs that could have been averted with any additional health care system resources required to implement it, or, if the intervention saves health care system costs, it is the DALYs averted by the intervention plus the DALYs that can also be averted with the cost savings offered.[14] In other words, net DALYs is a measure of the health effects of an intervention that encompasses the full implications of the intervention being delivered by the health care system. To calculate net DALYs, we assume a cost-effectiveness threshold of $500 USD in our primary analysis. A cost-effectiveness threshold of $500 USD per DALY averted approximates the cost-effectiveness threshold used for HIV treatment decisions and presumes that implementation is likely at least partially donor-supported.[15] We also conduct sensitivity analysis, varying the cost-effectiveness threshold to values below $500 USD to investigate scenarios where funding is provided by domestic sources.

### Section 2. Demographic model

The model runs from 1989 (assumed to be the start of the HIV epidemic) with variables updated in 3-month periods. Each run of the simulation program creates 100,000 simulated people who will be age 15 or above at some point between 1989 and 2071. The model is programmed in SAS. For each model run we scale up the outputs to a total adult population size in the current base year (i.e. usually the year we are living in at the time or the following year) of 10,000,000.

#### 2.1 General population death rates and determination of age in 1989

The initial age distribution for both males and females is sampled for each population simulation from three possible distributions representing three different population demographic structures (Table S1). These are chosen such that in the absence of HIV and cardiovascular disease, and given the death rates assumed (Table S2, Table S3), the resulting population pyramids and growth rates represent the range of those seen across the setting-scenarios.[16] Thus a proportion of simulated people have an age below 15 in 1989 (and most are yet to be born). The only variables that are modelled and updated up to reaching the age of 15 are age and sex. The “youngest” person in 1989 is age -67 (i.e. will be born in 2056 and reach age 15 in 2071, just before the modelled period ends. The model assumes that 50% of the population is male and 50% female.

##### Table S1. Distribution of ages of simulated individuals in 1989

| | Probability of being in age group in 1989 | | |
|---|---|---|---|
| **Age group** | **Population demographic structure 1** | **Population demographic structure 2** | **Population demographic structure 3** |
| -68 to -56 | 0.180 | 0.150 | 0.128 |
| -55 to -46 | 0.165 | 0.130 | 0.119 |
| -45 to -36 | 0.144 | 0.120 | 0.113 |
| -35 to -26 | 0.114 | 0.110 | 0.104 |
| -25 to -16 | 0.090 | 0.100 | 0.097 |
| -15 to -6 | 0.080 | 0.090 | 0.090 |
| -5 to 4 | 0.068 | 0.080 | 0.081 |
| 5 to 14 | 0.047 | 0.065 | 0.074 |
| 15 to 24 | 0.036 | 0.048 | 0.060 |

| | | | |
|---|---|---|---|
| 25 to 34 | 0.027 | 0.040 | 0.050 |
| 35 to 44 | 0.021 | 0.030 | 0.038 |
| 45 to 54 | 0.016 | 0.021 | 0.026 |
| 55 to 64 | 0.012 | 0.016 | 0.020 |

Age specific death rates for uninfected people are based on death rates in South Africa in 1997 (before the significant impact of HIV-related deaths) and are shown in Table S2. We use mortality data from South Africa in 1997 because South Africa has national death registry data and in 1997, mortality estimates were not yet heavily influenced by the HIV pandemic. We removed CVD deaths from these age-specific death rate estimates using a multiplier of 0.97 for ages 40-49 and 0.9 for age ≥50, based on approximate estimates of the proportion of all-cause mortality contributed by CVD (ischemic heat disease and stroke).[17] Non-HIV tuberculosis deaths are also removed for the HIV-negative population using a multiplier of 0.93 as these deaths are explicitly modeled in the Synthesis model.

##### Table S2. Age specific death rates (per year) in 1989

| Age group | Annual death rate | Annual non-CVD death rate | Age group | Annual death rate | Annual non-CVD death rate |
|---|---|---|---|---|---|
| Males | | | Females | | |
| 15 – 19 | 0.0020 | 0.0020 | 15 – 19 | 0.0015 | 0.0015 |
| 20 – 24 | 0.0032 | 0.0032 | 20 – 24 | 0.0028 | 0.0028 |
| 25 – 29 | 0.0058 | 0.0058 | 25 – 29 | 0.0040 | 0.0040 |
| 30 – 34 | 0.0075 | 0.0075 | 30 – 34 | 0.0040 | 0.0040 |
| 35 – 39 | 0.0080 | 0.0080 | 35 – 39 | 0.0042 | 0.0042 |
| 40 – 44 | 0.0100 | 0.0097 | 40 – 44 | 0.0055 | 0.0053 |
| 45 – 49 | 0.0120 | 0.0116 | 45 – 49 | 0.0075 | 0.0073 |
| 50 – 54 | 0.0190 | 0.0171 | 50 – 54 | 0.0110 | 0.0099 |
| 55 – 59 | 0.0250 | 0.0225 | 55 – 59 | 0.0150 | 0.0135 |
| 60 – 64 | 0.0350 | 0.0315 | 60 – 64 | 0.0210 | 0.0189 |
| 65 – 69 | 0.0450 | 0.0405 | 65 – 69 | 0.0300 | 0.0270 |
| 70 – 74 | 0.0550 | 0.0495 | 70 – 74 | 0.0380 | 0.0342 |
| 75 – 79 | 0.0650 | 0.0585 | 75 – 79 | 0.0500 | 0.0450 |
| 80 – 84 | 0.1000 | 0.0900 | 80 – 84 | 0.0700 | 0.0630 |
| ≥85 | 0.4000 | 0.3600 | ≥85 | 0.1500 | 0.1350 |

We updated age-specific death rates using 2019 Global Burden of Disease data for all-cause mortality, subtracting mortality rates for ischemic heart disease, stroke, and HIV (Table S3).[17] Similar to the approach in 1989, we removed non-HIV tuberculosis deaths using a multiplier of 0.93 in the population of persons without HIV. We assumed a linear change in non-HIV non-CVD death rates between 1989 and 2019 and calculated time-updated age-specific death rates for each period. We assume that age-specific death rates remain constant after 2019.

##### Table S3. Age specific death rates (per year) in 2019

| Age group | Annual death rate | Age group | Annual death rate |
|---|---|---|---|
| Males | | Females | |
| 15 – 19 | 0.00128 | 15 – 19 | 0.00081 |
| 20 – 24 | 0.00189 | 20 – 24 | 0.00110 |
| 25 – 29 | 0.00226 | 25 – 29 | 0.00139 |
| 30 – 34 | 0.00274 | 30 – 34 | 0.00175 |
| 35 – 39 | 0.00342 | 35 – 39 | 0.00229 |
| 40 – 44 | 0.00472 | 40 – 44 | 0.00310 |
| 45 – 49 | 0.00643 | 45 – 49 | 0.00419 |
| 50 – 54 | 0.00954 | 50 – 54 | 0.00608 |
| 55 – 59 | 0.01388 | 55 – 59 | 0.00891 |
| 60 – 64 | 0.02045 | 60 – 64 | 0.01353 |
| 65 – 69 | 0.02913 | 65 – 69 | 0.01953 |
| 70 – 74 | 0.04279 | 70 – 74 | 0.03046 |
| 75 – 79 | 0.06150 | 75 – 79 | 0.04494 |
| 80 – 84 | 0.09105 | 80 – 84 | 0.07249 |
| ≥85 | 0.14833 | ≥85 | 0.12256 |

### Section 3. Detailed hypertension modeling parameters

#### Table S4. Parameter distributions sampled for each model run together with the distribution for the 3000 setting-scenarios.

Table S4 provides details on parameters that are sampled at the beginning of each model run and remain constant over time for that model run. These parameters are intended to reflect conditions across a given setting-scenario (e.g. a given district, region, country for which estimates are desired).

| Setting-level parameters | Description | Distribution sampled (value; % with value) | Motivation for distribution |
|---|---|---|---|
| ***Population demographics*** | | | |
| *inc_cat* | Three future demographic structures with differing levels of population growth. | 1: 33%, 2: 33%, 3: 33% | Different countries in Africa have different population growth rates so we consider a range from around 1% to 3% per year[18] |
| ***Parameters relating to hypertension*** | | | |
| *prob_sbp_increase* | Population level risk of a 1 mmHg increase in true systolic blood pressure (SBP) per 3-month period. | 0.1: 33% 0.125: 33% 0.15 33% | Informed by average SBP increase over time across studies in Africa[19,20] |
| *rr_sbp_inc_on_antihyp* | Relative risk of a 1 mmHg increase in true SBP per 3-month period if on antihypertensive medication | 0.5 | Assumption that underlying SBP may still rise while on antihypertensive treatment, but at an attenuated rate. |
| *prob_symp_hypertension* | Probability of developing symptoms of hypertension during each time period that SBP ≥180 mmHg | 0.025 | Up to 54% of females and 48% of males with severe hypertension (SBP ≥180) may have symptoms attributable to hypertension, among a cohort of patients accessing regular medical care.[21] Prevalence of symptoms/symptom awareness may be lower among patients not routinely engaged in care.<br><br>Probability of 0.025 per period yields a cumulative incidence of hypertensive symptoms of 26% at 3 years and 40% at 5 years. |
| *measurement_error_var_sbp* | Random measurement error with measured vs true SBP | Normal distribution with standard deviation of 10 mmHg | Selected based on data from studies comparing differences in SBP between routine clinical measures and research measurements.[22–26] We model blood pressure measured using an average of 2-3 measurements. |
| *p_hard_reach_w*<br><br>*hard_reach_higher_in_men_p*<br><br>*hard_reach_lower_htn* | Proportion who have a propensity to not engage in health services. Individuals who are "hard to reach" will not participate in clinic- or community-based hypertension screening unless they are experiencing symptoms of severe hypertension (see *symp_hypertension* below). Similarly, such individuals will also not engage in HIV testing. | Proportion hard to reach in women: uniform(0.05, 0.2)<br>Absolute increased proportion hard to reach among men: uniform(0, 0.1)<br>Absolute decreased proportion hard to reach in hypertension: uniform(0, 0.05) | We assume that some individuals will not participate in HIV or hypertension screening due to individual factors that are independent of policy interventions (e.g. attitudes and beliefs, stigma, other individual determinants of health seeking behaviour). The mean proportion of 'hard to reach' persons is 12.5% (range 5-20%) among women and 17.5% (range 5-30%) among men. We assume that there are a common set of factors associated with uptake of both HIV and hypertension services, though that in some settings lower hypertension-associated stigma may result in fewer individuals being hard to reach for hypertension care (0-5%; thus combined hard-to-reach proportion for hypertension is 10% for women (range 0-20%) and 15% for men (range 0-30%). |

| Setting-level parameters | Description | Distribution sampled (value; % with value) | Motivation for distribution |
|---|---|---|---|
| | | | Participation in population-level hypertension screening during SEARCH study:[12]<br>- Attended 1st community-wide screening (year 0): 73% participation (77% among age 40-79)<br>- Attended ≥1 out of 3 annual community-wide screenings (year 0, 1, and/or 2): 86% (89% among age 40-79)<br>- Thus 14% of adults did not participate in any of three annual community-based hypertension screening events |
| *prob_test_sbp_undiagnosed* | Probability of blood pressure (BP) measurement per period in a person with no diagnosed hypertension and in the context of no additional BP screening | 0.01 | Informed by the proportion of persons with hypertension diagnosed in Africa.[27] |
| *prob_test_sbp_diagnosed* | Probability of BP measurement per period in a person with diagnosed hypertension but currently not in care for hypertension | 0.05 | There is limited data on re-engagement in blood pressure measurement/hypertension care among those with a known hypertension diagnosis. We selected this parameter based on model output that approximated real-world data on retention in care and hypertension treatment.[13,27–29] |
| *rr_htn_diagnosis* | Relative risk of the frequency of hypertension screening in a given setting. Multiplier of probability of BP measurement among both undiagnosed and diagnosed | 0.67: 33%, 1: 33%, 1.5: 33% | Incorporates range of uncertainty around probability of BP measurement with the assumption that the probability of presenting to care for measurement/remeasurement is similarly low, average, or high among those with and without a prior hypertension diagnosis. |
| *rr_test_sbp_women* | Multiplier of probability of BP measurement among women | 1.0: 33%, 1.1: 33%, 1.2: 33% | Relative risk of blood pressure measurement among women compared to men in Africa was 1.10 (95% CI 1.08-1.12) in multivariate regression with all countries having the same weight.[27] |
| *rr_test_sbp_young* | Relative risk of BP measurement among people <40 years of age (multiplier of base probability for younger individuals) | 0.5 | Empirically chosen to match real-world data. |
| *rr_test_symp* | Relative risk of BP measurement among those with symptoms of hypertension | 2 | Empirically chosen to reflect greater health-seeking behaviour in the presence of symptoms |
| *prob_start_htn_tx_s1* | Probability of initiating anti-hypertensive at clinic visit in an individual diagnosed with hypertension who has never been on anti-hypertensive medication and BP is 140-159/90-99 mmHg | 0.3: 33%, 0.4: 33%, 0.5: 33% | There is limited data on medication initiation at hypertension care visits. We selected this parameter based on control-group data from the SEARCH study and model output that approximated real-world data on hypertension treatment.[13,27–29] |
| *prob_start_htn_tx_s2* | Probability of initiating anti-hypertensive at clinic visit in an individual diagnosed with hypertension who has never been on anti-hypertensive medication and BP is ≥160/100 mmHg | 0.6: 33%, 0.7: 33%, 0.8: 33% | |

| **Setting-level parameters** | **Description** | **Distribution sampled (value; % with value)** | **Motivation for distribution** |
|---|---|---|---|
| *prob_restart_htn_tx_s1* | Probability of re-initiating prior anti-hypertensive medications at clinic visit when BP is 140-159/90-99 mmHg | 0.9: 33%, 0.95: 33%, 1: 33% | Assumption that hypertensive patients presenting to clinic off of their medications will be restarted on their previous medication regimen on the majority of visits. |
| *prob_restart_htn_tx_s2* | Probability of re-initiating prior anti-hypertensive medications at clinic visit when BP is ≥160/100 mmHg | 0.9: 33%, 0.95: 33%, 1: 33% | |
| *htn_retention_patt* | Pattern of visit retention<br><br>If on antihypertensives and due for a visit (or if new diagnosis after initial visit with SBP 140-159 and given lifestyle counselling), the probability of return for subsequent visit.<br><br>Probability of return increases with each consecutively attended visit.<br><br>Persons who do not have a visit fall out of care. If they subsequently re-enter care, the visit number restarts at 1. | 33% pattern 1:<br>*prob_visit_htn_v1*: 0.57<br>*prob_visit_htn_v2*: 0.67<br>*prob_visit_htn_v3*: 0.76<br>*prob_visit_htn_v4*: 0.81<br>*prob_visit_htn_v5*: 0.86<br><br>33% pattern 2:<br>*prob_visit_htn_v1*: 0.60<br>*prob_visit_htn_v2*: 0.70<br>*prob_visit_htn_v3*: 0.80<br>*prob_visit_htn_v4*: 0.85<br>*prob_visit_htn_v5*: 0.90<br><br>33% pattern 3:<br>*prob_visit_htn_v1*: 0.63<br>prob_visit_htn_v2: 0.74<br>prob_visit_htn_v3: 0.84<br>prob_visit_htn_v4: 0.89<br>prob_visit_htn_v5: 0.90 | Probability of retention in hypertension care based on control arm of SEARCH study demonstrating 17% with consistent care engagement (≥2 visits per year) over 3 years.[27] Hypertension medications were available free of charge in the SEARCH control arm, though visit was provided through standard outpatient department care. Thus, in real-world implementation retention in care may be even lower than what was observed in SEARCH.<br><br>Assuming a 5% rate of re-engagement in care in each quarter and quarterly visits, modelled visit probabilities will yield the following proportion retained in care after 3 years: 26% retained (range 20-29%). We assumed slightly higher retention in care than that observed in SEARCH to account for observed data of persons on hypertension treatment. |
| *interval_visit_hypertension* | Interval between visits for a person on antihypertensives and with most recent measured SBP <140 | 0.25 (once every 3 months) | WHO guidelines suggest follow-up every 3-6 months for patients with controlled hypertension and monthly follow-up after initiation or a change in antihypertensive medications until BP targets are reached.[6] Under standard of care conditions, we assume that return visits are scheduled every 3 months. |
| *prob_intensify_1_2* | For a person on 1 antihypertensive medication with current SBP >=140, probability of adding a second antihypertensive | 0.10: 33%, 0.15: 33%, 0.20: 33%<br><br>Probability 1.5-fold higher if SBP 160-179 mmHg and 2-fold higher if SBP ≥180 mmHg | In the SEARCH median control arm community (where medications were available), medications were intensified from 1 to 2 drugs 15% (IQR 5-36%) of the time when SBP was 140-159 mmHg, 23% (IQR 16-49%) when SBP was 160-179 mmHg, and 61% (42-92%) when SBP was ≥180 mmHg.[13] We approximated a lower probability of intensification to account for settings where medications are not routinely available or are costly.[30] |
| *prob_intensify_2_3* | For a person on 2 antihypertensive medications with current SBP >=140, probability of adding a third antihypertensive | 0: 33%, 0.02: 33%, 0.04: 33% | In the SEARCH median control arm community (where medications were available), medications were intensified from 2 to 3 drugs 0% (IQR 0-3%) of the time when SBP was 140-159 mmHg, 1% (IQR 0-3%) when SBP was 160-179 mmHg, and 8% (3-16%) when SBP |

| Setting-level parameters | Description | Distribution sampled (value; % with value) | Motivation for distribution |
|---|---|---|---|
| | | Probability 1.5-fold higher if SBP 160-179 mmHg and 2-fold higher if SBP ≥180 mmHg | was ≥180 mmHg.[13] We approximated a lower probability of intensification to account for settings where medications are not routinely available or are costly.[30] |
| *baseline_ihd_risk* | Setting-level baseline risk for myocardial infarction to model risk in both high and average risk settings | 50% 1.8 $x10^{-5}$,<br>50% 2.7 $x10^{-5}$ | We estimate incidence of ischemic heart disease based on Global Burden of Disease data for incident myocardial infarction.[17] |
| *baseline_cva_risk* | Setting level baseline risk for stroke to model risk for both high and average risk settings. | 40% 4.0 $x10^{-6}$,<br>40% 6.0 $x10^{-6}$,<br>20% 7.5 $x10^{-6}$ | We estimate stroke incidence using data from Global Burden of Disease data for stroke and from a community-based study assessing stroke incidence in both rural and urban Tanzania, sampling from two baseline risk parameters to estimate stroke incidence in both high and low incidence settings (e.g. urban vs rural).[31] Stroke incidence in the Tanzania study was substantially higher than estimates from the Global Burden of Disease; to account for this, we a range of baseline risk estimates to account for a range of stroke that included both data sources, with a lower proportion of setting-scenarios drawing from the higher risk found in Walker et al.. |
| *prob_ihd_tx* | Base probability of receiving acute treatment for myocardial infarction | Mild / silent MI / angina: 0<br>Moderate/severe myocardial infarction: 0.4 | We assume a base case where 40% of persons with moderate or severe acute MI or stroke receive emergency care, and that 75% of those receive care that is not effective for reducing acute mortality (thus 10% receive effective emergency care in the base case).<br><br>These assumptions are based on data that that between 1 to 17% of patients in Africa with stroke were admitted to a stroke unit within 3 hours of symptom onset.[32] In the absence of similar data for acute myocardial infarction, we assume a similar proportion of patients with myocardial infarction receive timely and effective emergency care. |
| *prob_cva_tx* | Base probability of receiving acute treatment for stroke | Mild stroke: 0<br>Moderate/severe stroke: 0.4 | |
| *prob_ihd_tx_effective* | Probability that emergency care for myocardial infarction is effective (associated with mortality risk reduction) | 0.25 | Globally, more than 8 million people in low- and middle-income countries die due to poor-quality healthcare. More than 80% of avoidable deaths due to cardiovascular deaths are due to poor quality care, with the remaining due to non-utilization of health systems.[33] Though data on accessing emergency care for acute myocardial infarction and stroke is limited, we assume a base scenario where 40% of individuals with clearly symptomatic events (moderate/severe MI or CVA) seek care in the hospital (prob_ihd_tx and prob_cva_tx). We further assume a base scenario where 25% of care is effective and the remaining 75% of such care is ineffective, either due to late presentation or poor quality/inadequate health services. Because such ineffective care may partially involve lower level health facilities or less utilization of diagnostic or therapeutic interventions, we also assume a lower cost associated with accessing ineffective care (see **Table S9**). |
| *prob_cva_tx_effective* | Probability that emergency stroke care is effective (associated with mortality risk reduction) | 0.25 | |

| Setting-level parameters | Description | Distribution sampled (value; % with value) | Motivation for distribution |
|---|---|---|---|
| *rr_cvd_tx* | Relative risk of receiving acute treatment for myocardial infarction or stroke | 33%: 0.5, 33%: 1.0, 33%: 2.0 | Multiplier of prob_ihd_tx and prob_cva_tx to account setting level variation in availability and uptake of acute treatment for acute CVD events, assuming collinearity with low/higher utilization of acute care for both MI and CVA. This results in a range of acute CVD treatment seeking from 20% to 80% across setting-scenarios. |
| *rr_cvd_tx_effective* | Relative risk of receiving effective emergency treatment for myocardial infarction or stroke | 33%: 0.5, 33%: 1.0, 33%: 2.0 | Multiplier of prob_ihd_tx_effective and prob_cva_tx_effective to account for setting level variation in availability and uptake of effective acute treatment for acute CVD events, assuming collinearity of care effectiveness for both MI and CVA. When combined with variability in the probability of treatment seeking, this results in a range of the proportion of acute MI/stroke events that receive effective acute care from 2.5% to 40% across setting-scenarios. |
| *rr_mort_ihd_tx* | Relative risk of mortality with effective treatment for acute myocardial infarction | 0.8 | Given lack of access to fibrinolysis or cardiac catheterization (see risk_cvd_death in **Table S6** for discussion of advanced therapies), we assume that individuals receiving effective emergency treatment for acute myocardial infarction receive aspirin and supportive care. Treatment with antiplatelet therapy for one month in the setting of acute MI is associated with a relative risk for mortality of approximately 0.80 (i.e. 20% risk reduction).[34] |
| *rr_mort_cva_tx* | Relative risk of mortality with effective treatment for acute stroke | 0.6 | Treatment is assumed to be supportive care and aspirin (see risk_cvd_death in **Table S6** for discussion of advanced therapies). Treatment with antiplatelet therapy for one month in the setting of acute stroke is associated with a relative risk reduction for mortality of approximately 0.94.[34] A pilot study of a stroke unit focused on supportive care demonstrated a very high relative risk reduction for mortality of 0.33 (67% reduced mortality), largely by managing blood pressure, managing glucose, treating infections, and preventing aspiration pneumonia.[35] We assume a more modest risk reduction associated with acute stroke care due to incomplete treatment implementation or adherence of this package of interventions. |

#### Table S5. Individual variables defined at the beginning of each model run

Table S5 describes variables that are sampled for each individual person at the beginning of each model run. These variables are intended to reflect variability between individuals within the population.

| Individual-level variable (fixed) | Description | Distribution sampled (value; % with value) | Justification for variable distribution |
|---|---|---|---|
| *sbp_age15* | True SBP modelled for each individual at age 15. Individual | At age 15, SBP sampled from the following values derived from a normal distribution with mean 112, standard deviation 20, and truncation at 95 and 165. SBP at age 15 is then increased by 2 for males and decreased by 2 for females.<br><br>95: 27%, 105: 20%, 115: 19%, 125: 16%, 135: 10%, 145: 5%, 155: 2%, 165: 1%<br><br>Males: +2 mmHg<br>Females: -2 mmHg | Available population-based surveys do not measure SBP in 15-year-olds. This distribution is based on data from SEARCH with population-level blood pressure measurement in 32 rural communities in Kenya and Uganda for adults aged 18 and older (n = 6,534 for age 18-19)[12] and population-level surveys with age ≥20.[20] |
| *sbp_rr* | Individual modifier (i.e. sampled once for each individual in each setting scenario) for risk of 1 mmHg increase in SBP per 3-month period (multiplicative with setting-specific prob_sbp_increase) | 0.67: 33%, 1.0: 33%, 1.5: 33% | We use a range of values centred around 1 to model the within-person variability in SBP rise over the life course. |
| *sbp_rr_age* | Individual modifier for risk of 1 mmHg increase in SBP from age 45 to <65 (multiplicative with prob_sbp_increase and sbp_rr). | 1: 33%, 2: 33%, 3: 33% | Informed by higher average SBP increase during middle-age across studies in Africa.[20] We use a range of values for this multiplier to reflect within-person variability in higher SBP rise during middle-age. |
| *effect_anti_hyp_1* | Person-specific SBP reduction (mmHg) of the first antihypertensive drug added, should the participant take them | 10: 70% 20: 20% 30: 10% | Informed by an instrumental variable analysis of SPRINT data where each additional drug reduced SBP by 14.4 mmHg.[36]<br><br>We estimated a person-specific effect for each drug of 10, 20, or 30 mmHg, weighted to yield a mean effect for each drug of 14 mmHg. |
| *effect_anti_hyp_2* | Person-specific SBP reduction (mmHg) of the second antihypertensive drug added, should the participant take them | 10: 70% 20: 20% 30: 10% | |
| *effect_anti_hyp_3* | Person-specific SBP reduction (mmHg) of the third antihypertensive drug added, should the participant take them | 10: 70% 20: 20% 30: 10% | |
| *hard_reach_htn* | Person-specific indicator that an individual will not engage in screening for hypertension services. Individuals who are "hard to reach" will not participate in clinic- or community-based hypertension screening unless they are experiencing symptoms of severe hypertension (see *symp_hypertension* below). | The mean proportion of 'hard to reach' persons for hypertension screening or treatment is 10% (range 2.5-20%) among women and 15% (range 2.5-30%) among men. | Probability of an individual person being 'hard to reach' for hypertension services is defined by setting-level variables hard_reach_w, *hard_reach_higher_in_men_p*, and *hard_reach_lower_htn* (see **Table S4**). |

#### Table S6. Individual variables calculated/updated each time step

Table S6 describes individual-level variables that are updated during each time step of the model. For example, true underlying systolic blood pressure is modeled for each individual and updated each time step of the model.

| Individual-level Variable (time-updated) | Description | Calculation | Justification |
|---|---|---|---|
| *sbp* | True underlying systolic blood pressure (SBP), regardless of whether measured | Each time step, the probability of having a 1 mmHg increase is defined by *prob_sbp_inc * sbp_rr * sbp_rr_age (if age >=40 & <60)* | SBP tends to increase over time in a given individual. Model calibrated with data from the Global Burden of Disease Survey and the NCD Risk Factor Collaboration.[19,20] |
| *symp_hypertension* | Presence of symptomatic hypertension | Each time step where *sbp* >180, probability of symptom onset is defined by prob_symp_hypertension. Once symptoms begin, they persist until *sbp* <180 | See *prob_symp_hypertension* in **Table S4**. |
| *test_sbp_comm* | Indicator that an individual was tested for hypertension during community-based screening in the current 3-month period (if such screening is included in the modelled intervention scenario) | If a community screening event is conducted during the time step (defined by *comm_test_interval*), probability of screening is defined by *prob_test_sbp_comm*. If an individual is hard to reach (*hard_reach* =1), the probability of screening is 0 unless they have symptomatic hypertension (*symp_hypertension* = 1). | See *prob_test_sbp_comm* in **Table S8**. |
| *sbp_m_comm* | Value of measured systolic blood pressure at community screening, if blood pressure is measured (i.e. *test_bp_comm* = 1) | Defined by *sbp* + random error with standard deviation *measurement_error_var_sbp* | See *measurement_error_var_sbp* in **Table S4**. |
| *tested_bp* | Indicator that an individual had their blood pressure measured at a clinic in the current 3-month period. Individuals can present spontaneously, or after community-based screening if sbp was >=140 and they linked to care. | Probability of blood pressure measurement determined by:<br>• If never diagnosed with hypertension: *prob_test_sbp_undiagnosed*<br>• If previously diagnosed with hypertension but out of care: *prob_test_sbp_diagnosed*<br>• If tested at community-based screening and SBP >=140: *prob_htn_link*<br><br>If *tested_bp* = 1 in a given period, the value of the measured systolic blood pressure is defined by *sbp_m* | See *prob_test_sbp_undiagnosed and prob_test_sbp_diagnosed* in **Table S4** and **Table S8**. |
| *sbp_m* | Value of measured systolic blood pressure at a clinic, if blood pressure is measured (i.e. *tested_bp =1)* | Defined by *sbp* + random error with standard deviation *measurement_error_var_sbp* | See *measurement_error_var_sbp* in **Table S4**. |

| Individual-level Variable (time-updated) | Description | Calculation | Justification |
|---|---|---|---|
| *htn_true* | Indicator that an individual has true hypertension (true underlying sbp >=140 mmHg), regardless of whether blood pressure has been measured or they have been diagnosed. | *htn_true* = 1 if maximum *sbp* >=140 (i.e. pre-treatment sbp if on antihypertensives)<br><br>Once htn_true = 1, it remains 1 throughout the person's lifetime. | |
| *diagnosed_hypertension* | Indicator that an individual has received a hypertension diagnosis. | *diagnosed_hypertension* = 1 if *sbp_m* >=140 on two consecutive clinic measurements OR *sbp_m* >=160 on one occasion OR antihypertensive medications are initiated | Guidelines indicate that a diagnosis of hypertension should be made over 2 or more visits with consistent elevated BP. If BP is more severely elevated, a diagnosis can be made on the first visit with medications initiated on the same day.[6,37] |
| *visit_hypertension* | Indicator that an individual had a visit for hypertension screening, diagnosis, or treatment in the current time step | Probability of having a visit for hypertension:<br>• If *visit_hypertension* =0 at time t-1 (i.e. the previous 3-month period), a hypertension visit occurs if *tested_bp* =1 in the current 3-month period<br>• If *visit_hypertension* =1 at time t-1 and due for a visit this period, the probability of attending a follow-up visit is defined by *prob_visit_htn_v#* where # is defined by *htn_visit_count* (the number of consecutive previously attended hypertension visits) | |
| *htn_visit_count* | Count variable of consecutive attended *visit_hypertension* when due for a visit. This variable resets to 0 if that individual falls out of care. | Increases by 1 for each consecutive attended visit for hypertension (*visit_hypertension* ==1) | |
| *on_anti_hypertensive* | Number of antihypertensive medications the patient is taking this period (range 0-3) | If *sbp_m* >=140 and not yet on antihypertensives, probability of initiating 1 drug this period (assume start with 1 drug) defined by:<br>• If *sbp_m* 140-159: *prob_start_htn_tx_s1*<br>• If *sbp_m* >=160: *prob_start_htn_tx_s2*<br><br>If already on antihypertensives and sbp_m >=140, probability of adding an additional drug defined by:<br>• 1 -> 2 drugs: *prob_intensify_1_2*<br>• 2 -> 3 drugs: *prob_intensify_2_3*<br>• For drug intensification, probability increased by 1.5-fold if *sbp_m* 160-179 and 2-fold if *sbp_m* >=180 | |

| Individual-level Variable (time-updated) | Description | Calculation | Justification |
|---|---|---|---|
| | | The model assumes that only one drug is started/added per 3-month time step. | |
| *risk_ihd* | Risk of having a myocardial infarction per period | *Baseline_ihd_risk* * exp(((*age* – 15) x 0.07) + (0.4 if male) + ((*sbp*-115) x *ihd_sbp_age_effect*) + *prior_cvd* * 1.8)<br><br>*ihd_sbp_age_effect*:<br>• Age <60: 0.035<br>• Age 60-79: 0.03<br>• Age ≥80: 0.02 | Overall incidence: There are no population-level studies to inform true incidence of acute myocardial infarction (AMI) in Africa. The Global Burden of Disease consortium estimates incidence using available data from hospital-based cohorts and extrapolations using covariates,[17] though this likely underestimates incidence by excluding those who do not seek medical care or who die prior to hospitalization.[38] We use data from the 2019 Global Burden of Disease Study data to calibrate estimates, assuming that many mild cases are not captured in surveillance studies and not reflected in available incidence data. We therefore calibrate the incidence of moderate or severe AMI to available data. Additionally, we calibrate incidence to a study Hertz et al that combined community surveillance with modelling to estimate the total number of AMI including those missed by hospital surveillance.[38] This study estimated an overall age-standardized incidence of AMI of 211 per 100,000 (263 per 100,000 among men and 170 per 100,000 among women), which mirrors Global Burden of Disease estimates. Overall incidence estimated by Hertz et al ranged from 155 to 341 per 100,000 in rural and urban communities, respectively.<br><br>Effect of SBP and age on AMI incidence: Meta-analysis of randomized trials estimates a 0.74 relative risk reduction in major CVD events per 10 mmHg reduction in SBP among individuals without existing CVD.[9] Another meta-analysis found a similar relative risk of 0.79 per 10 mmHg reduction in SBP for AMI events over the first year of follow-up among individuals without a known history of CVD, suggesting that benefits are realized even over near-term follow-up.[10] In a large meta-analysis, reduction in SBP by 20 mmHg reduced mortality from ischemic heart disease by 0.49 for individuals aged 40-49 and by 0.60 for individuals aged 70-79.[11] We modelled age-specific effects of SBP on incidence of AMI based on Lewington et al,[11] such that a 20 mmHg reduction in SBP corresponded to a RR of 0.5 (exp(-20*0.035)) for age <60, RR of 0.55 for age 60-79, and RR of 0.67 for age ≥80 years. This corresponds to relative risks of 0.70, 0.74, and 0.82 for a 10 mmHg SBP reduction, respectively. We model risk associated with SBP down to a SBP of 115, consistent with multiple studies showing benefit in reduction of CVD events down to a SBP of 115 with no substantial heterogeneity of relative treatment effect by starting SBP.[9,11] We assume that there is no interaction in the relative risk reduction of AMI risk by absolute baseline SBP or prior CVD history, consistent with above cited studies.<br><br>Effect of sex on AMI incidence: Numerous studies have reported a higher incidence of AMI in men than women, with studies from high income settings reporting as high as 2-fold increased risk in men that persists throughout the life course.[39] In the large INTEHEART case control study, women had their first AMI approximately 9 years later |

| Individual-level Variable (time-updated) | Description | Calculation | Justification |
|---|---|---|---|
| | | | than men.[40] Crude incidence rates in Africa are approximately 1.5-fold higher in men than women.[17,38] We modelled a relative risk for incident AMI of 1.5 for male sex (exp(0.4)).<br><br>Effect of CVD on AMI incidence: Prior studies in high-income settings before AMI therapies were available suggest an approximate 10-fold greater risk of recurrent AMI than risk of first AMI among individuals with unfavourable risk factors.[41] These studies were likely biased toward more severe presentations given limited availability of therapy, thus we conservatively estimated a 6-fold increase (exp(1.8)) in risk of recurrent myocardial infarction following an initial CVD event (stroke or MI). |
| *risk_cva* | Risk of cerebrovascular accident (stroke) per period | *baseline_cva_risk* * exp(((*age* – 15) x 0.09) + ((*sbp*-115) x *cva_sbp_age_effect*) + *prior_cvd* * 1.8)<br><br>*cva_sbp_age_effect*:<br>• Age <60: 0.05<br>• Age 60-79: 0.04<br>• Age ≥80: 0.02 | Overall stroke incidence: There are few population-level studies of stroke (aka cerebrovascular accident, CVA) incidence in Africa. Hospital-based cohorts likely underestimate stroke incidence by excluding those who die prior to hospital admission or who do not seek medical care. We estimate stroke incidence using data from a community-based study assessing stroke incidence in both rural and urban Tanzania, sampling from two baseline risk parameters to estimate stroke incidence in both high and low incidence settings.[31] We also use data from the 2019 Global Burden of Disease Study data to calibrate estimates.[17] We assume that many mild strokes are not captured in surveillance studies and not reflected in available incidence data. We therefore use the incidence of moderate or severe strokes to calibrate stroke incidence to available data.<br><br>Effect of SBP and age on stroke incidene: We derived estimates for the association with SBP and stroke risk from meta-analyses of randomized trials of antihypertensive treatment on cardiovascular disease (CVD) events. Meta analyses have reported a range of estimates for relative risk reduction of acute stroke associated with SBP reduction:<br>• Ettehad et al:[9] RR 0.74 per 10 mmHg SBP reduction (similar for those with and without history of CVD)<br>• Law et al:[10] RR 0.54 per 10 mmHg SBP reduction if no history of CVD, RR 0.65 (95% CI 0.53-0.80) if history of coronary heart disease and RR 0.66 (95% CI 0.56-0.79) if history of stroke, with no significant difference in treatment effect comparing trials. Overall RR 0.59, 95% 0.52-0.67.<br>• Lewington et al:[11] RR 0.36 for 20 mmHg SBP reduction among age 40-49 and RR 0.50 for individuals aged 70-79.<br><br>We modelled age-specific effects of SBP on CVA incidence based on Lewington et al, such that a 20 mmHg reduction in SBP corresponded to a RR of 0.37 (exp(-20*0.05)) for age <60, RR of 0.45 for age 60-79, and RR of 0.67 for age ≥80 years. This corresponds to relative risks of 0.61, 0.67, and 0.82 for a 10 mmHg SBP reduction, respectively. We model risk associated with SBP down to a SBP of 115, consistent with |

| Individual-level Variable (time-updated) | Description | Calculation | Justification |
|---|---|---|---|
| | | | multiple studies showing benefit in reduction of CVD events down to a SBP of 115 with no substantial heterogeneity of relative treatment effect by starting SBP.[9,11] We assume that there is no interaction in the relative risk reduction of stroke risk by absolute SBP or prior CVD history.<br><br>Effect of sex on stroke incidence: Age-standardized stroke incidence rates are similar between men and women.[42] We therefore assumed similar risk of stroke among men and women.<br><br>Effect of CVD history on stroke incidence: We were unable to identify data from Africa on the relative risk of recurrent stroke associated with history of a prior stroke. We used data from the Community Stroke Study in Australia[43] to estimate a 6-fold increase in relative risk of stroke associated with prior history of cardiovascular disease (exp(1.8) if prior CVD). |
| *risk_cvd_hiv* | Relative risk of CVD event (IHD or CVA) with HIV | RR if CD4 ≥500 & viral load <500: 1.2<br>RR if CD4 <500 or viral load ≥1000: 1.44 ($1.2^2$)<br>RR if CD4 < 500 & viral load ≥1000: 2.07 ($1.2^4$) | HIV is associated with higher risk of CVD,[44] though studies comparing CVD people with and without HIV may be confounded both by differences in risk behaviours and differences in access to healthcare and/or health seeking behaviour. For example, people with HIV with a high CD4 count and suppressed viral load likely access clinical care regularly and may have differential access to preventive treatment. In the VACS study, HIV was associated with a 1.39 (95% CI 1.17-1.66) relative risk of MI compared to a risk-adjusted uninfected population if the HIV viral load was < 500 and 1.75 (95% CI 1.40-2.18) if the viral load was ≥500.[7] Similarly, in a population with access to medical care, HIV with CD4 >500 was associated with a 1.18-fold increased risk of MI (95% CI 0.96-1.45) compared to HIV uninfected population, with risk increased with decreasing CD4: RR 1.59 (95% CI 1.34-1.90) if CD4 200-500 and RR 1.76 (95% CI 1.31-2.37) if CD4 <200.[8] We assume a relative risk for CVD events of 1.2 for people with HIV with suppressed viral load and high CD4, with further increased risk associated with unsuppressed viral load and/or low CD4 count. |
| *ihd_severity* | Proportion of acute myocardial infarction/ischemic heart disease events that are mild, moderate, and severe | Mild: 60% (silent MI / Angina)<br>Moderate: 30% (MI)<br>Severe: 10% (MI) | There is limited population-level data to inform severity of acute myocardial infarction events in Africa. We chose these parameters as an estimate of severity based on calibration of incidence, prevalene and to model heterogeneity, calibrating model output to available data on incidence, prevalence, and mortality due to ischemic heart disease.<br><br>We assume that subsequent events have the same distribution of severity, though chronic mortality risk in subsequent periods cannot be downgraded by a later event – for example, if someone had a myocardial infarction of moderate severity, and then subsequently had a mild event, their future mortality risk and disability weights remain at the moderate level following the period of their repeat acute event. |
| *cva_severity* | Proportion of acute CVA events that are | Mild: 60%<br>Moderate: 30% | Available data on stroke severity in Africa is limited to hospital-based cohorts which likely miss many mild cases and may also miss strokes |

| Individual-level Variable (time-updated) | Description | Calculation | Justification |
|---|---|---|---|
| | mild, moderate, and severe | Severe: 10% | that result in pre-hospital deaths. In a hospital-based cohort in The Gambia (n=148), presenting stroke severity was mild in 19%, moderate in 42% and severe in 41%.[45] Overall mortality was 52% at 3 months and 62% at 1 year. Stratifying by presenting severity, one year mortality was 20% in mild, 52% in moderate, and 89% in severe strokes respectively.<br><br>Based on trial data with presumed comprehensive ascertainment of all strokes regardless of severity,[46] we assume that 60% of strokes are mild, 30% moderate, and 10% severe. Stroke severity corresponds to both mortality (see *risk_cvd_death* below) and disability weights (see **Table S7**).<br><br>We assume that subsequent events have the same distribution of severity, though severity cannot be downgraded by a subsequent event – for example, if someone had a stroke of moderate severity, and then subsequently had a mild stroke, their future mortality risk and disability weights remain at the moderate level following the period of their repeat acute stroke. |
| *risk_cvd_death* | Acute myocardial infarction mortality | Acute mortality:<br>- Mild: 0.05<br>- Moderate: 0.2<br>- Severe: 0.4<br>Mortality in subsequent periods:<br>- Mild: 0.01<br>- Moderate: 0.02<br>- Severe: 0.04 | Hospital-based cohort studies in Africa have shown a wide range of outcomes following presentation for acute myocardial infarction, with 30-day mortality ranging from 7.8 to 43.3%.[47] Two thirds of the referral hospitals included in this study offered fibrinolysis and 56% offered percutaneous coronary interventions. While availability of such advanced therapies is increasing, implementation is limited by long pre-hospital delays (median 31 hours), limited number of facilities with needed expertise, and limited availability of advanced therapies for acute MI for individuals of low socioeconomic status or who lack insurance.[48] Another more recent study among patients diagnosed with acute myocardial infarction at a tertiary care centre in Tanzania found 3-month and 1-year mortality of 49% and 60%,[49] with no patients receiving fibrinolysis and only 5% consistently taking aspirin post-discharge.[50] As such, we assume that individuals with myocardial infarction do not receive advanced therapies (i.e. fibrinolysis or cardiac catheterization), thus not incurring their benefits (or associated costs).<br><br>We chose empiric estimates for acute mortality stratified by severity, yielding weighted 3-month and 1-year mortality estimates of 13% and 17%, respectively. These estimates are likely conservative, though they account for our modelled higher incidence of mild acute myocardial infarction/angina events that are likely undetected by hospital-based cohort studies. Stratified by severity, 1-year mortality estimates are 8% for mild, 25% for moderate, and 46% for severe acute ischemic heart disease events. |
| | Stroke mortality | Mortality, acute:<br>- Mild CVA: 0.1<br>- Moderate CVA: 0.3<br>- Severe CVA: 0.6 | Stroke mortality estimates from Africa are generally based on hospital-based cohort studies.[51] Such studies may be biased toward more severe cases, though may also exclude cases resulting in pre-hospital death. A recent systematic review estimated an in-hospital mortality of 22% from |

| Individual-level Variable (time-updated) | Description | Calculation | Justification |
|---|---|---|---|
| | | Mortality, subsequent periods<br>- Mild CVA: 0.01<br>- Moderate CVA: 0.03<br>- Severe CVA: 0.06 | acute stroke.[51] Studies with longer follow-up include estimates of 3- and 12-month mortality of 52% and 62%, respectively, in The Gambia[45] and 25% and 38% in South Africa.[52] In the study in The Gambia, 1-year mortality varied by presenting severity: 20% in mild, 52% in moderate, and 89% in severe strokes, respectively. In a community-based study in Tanzania that aimed to capture all incident strokes in the community (and thus was not biased by hospital presentation), 28-day mortality was 28.7% and 3-year mortality was 84.3%.[53] Though many studies of stroke mortality in Africa are dated, access to acute stroke care remains very limited in most settings and it is unlikely that contemporary acute mortality is substantially lower than prior estimates.[54]<br><br>Assuming that stroke incidence studies may underestimate the incidence of mild strokes, and therefore over-estimate average mortality rates, we used mortality estimates that led to lower mortality than available data. We estimated acute mortality of 10% for mild, 30% for moderate, and 60% for severe stroke, leading to a weighted acute mortality of 21% in the first 3-month period following the stroke. We estimated mortality in subsequent quarters as 1% in mild, 3% in moderate, and 6% in severe strokes. Ignoring competing risks and subsequent stroke events (thus under-estimating mortality), this yields an approximate overall 1-year mortality of 25% and 3-year mortality of 34%, corresponding to a 1-year mortality of 13% for mild, 36% for moderate, and 66% for severe strokes. |

#### Table S7. Disability Weights

| Condition | Disability weight | IHME GBD Disability Weight category[17] |
|---|---|---|
| *Stroke, mild (acute and chronic)* | 0.019 | Acute/Chronic ischemic stroke severity level 1 |
| *Stroke, moderate (acute and chronic)* | 0.316 | Acute/Chronic ischemic stroke severity level 3 |
| *Stroke, severe (acute and chronic)* | 0.588 | Acute/Chronic ischemic stroke severity level 5 |
| *Ischemic heart disease, mild (acute and chronic)* | 0.037 | Mild angina/heart failure due to ischemic heart disease (assume that initial myocardial infarction was minimally symptomatic) |
| *Ischemic heart disease, moderate (acute and chronic)* | Acute: 0.084<br>Chronic: 0.076 | Weighted average over 3 months of:<br>Acute myocardial infarction first 2 days and mean value of disability weight for moderate angina and moderate heart failure due to ischemic heart disease for remainder of 3-month period |
| *Ischemic heart disease, severe (acute and chronic)* | Acute: 0.179<br>Chronic: 0.173 | Weighted average over 3 months of:<br>Acute myocardial infarction first 2 days and mean value of disability weight for severe angina and severe heart failure due to ischemic heart disease for remainder of 3-month period |

#### Table S8. Intervention Parameters

Table S8 describes parameters for the intervention policies considered (chronic care clinic and community health worker + chronic care clinic). Parameters not included in Table S8 are assumed to be unaffected by the policy interventions considered and parameters are sampled from the preceding tables.

| Parameter (under intervention conditions) | Description | Sampled value | Justification |
|---|---|---|---|
| *prob_test_sbp_comm* | Setting-level probability of undergoing community-based blood pressure screening if eligible and screening occurs that period. **CHW policy only** | 0.75: 33%, 0.80: 33%, 0.85: 33% | In the community health worker (CHW) screening intervention, CHWs will attempt to screen all eligible individuals for hypertension on an annual basis, though we assume that some individuals will not be reached for screening in any given period when screening occurs. Some individuals will not be reached due to random chance, while others will not be reached due to individual-level characteristics that make them less likely to uptake health interventions (i.e. they are 'hard to reach', (see hard_reach_htn parameter in Table S5). In the model, 'hard to reach' individuals will not participate in screening unless they have symptomatic hypertension (SBP ≥180 + symptoms) or a history of moderate or severe CVD. The mean proportion of persons who are 'hard to reach' for hypertension screening is 10% (range 0-20%) among women and 15% (range 0-30%) among men.<br><br>Integrating prob_test_sbp_comm with hard_reach, an average of 70% of women and 65% of men will be reached for community-based CHW blood pressure screening (range 55-85% of women and 45-85% of men) during each period when screening occurs.<br><br>Preliminary data from a cluster randomized trial of home-based CHW screening for hypertension and HIV testing/risk assessment as part of the SEARCH collaboration, CHWs screened 89% (n= 13,697 of 15,450) of adults aged ≥40 years across eight communities in Kenya and Uganda.[55] Our model assumptions are conservative compared to high degrees of screening reach in this study. |
| *prob_htn_link* | Probability of linking to the clinic for hypertension care following community-based screening. **CHW policy only** | 33%: 0.5, 33% 0.6, 33%: 0.7 | Setting-level probability of linking to hypertension care if community-measured SBP ≥140 mmHg. In a recent study linkage to care was 66% following community-based screening by a CHW.[56] We conservatively use a range of linkage to care from 50-70%. |
| *prob_start_htn_tx_s1* | Initiation of antihypertensive medication if known diagnosis but not previously on medications when SBP is 140-159 mmHg. **CCC and CHW policies.** | 33%: 0.4, 33% 0.5, 33% 0.6 | In SEARCH intervention communities in Uganda (n=10), antihypertensive medication was initiated in persons diagnosed with hypertension who have not previously been on medications with the following frequency (community-level):<br>• SBP 140-159: 51% (IQR 42-62%)[13] |
| *prob_start_htn_tx_s2* | Initiation of antihypertensive medication if known diagnosis but not previously on medications when SBP is ≥160 mmHg. **CCC and CHW policies.** | 33%: 0.95, 33% 0.975, 33% 1.0 | In SEARCH intervention communities in Uganda (n=10), antihypertensive medication was initiated in persons diagnosed with hypertension who have not previously been on medications with the following frequency (community-level):<br>• SBP ≥160: 98% (97-100%)[13] |
| *prob_intensify_1_2* | For a person on one antihypertensive medication with current SBP ≥140, probability of adding a second antihypertensive | 0.25: 33%, 0.30: 33%, 0.35: 33%<br><br>Probability 1.5-fold higher if SBP 160-179 mmHg and 2-fold higher if SBP ≥180 mmHg | In the SEARCH median intervention arm community (where medications were available), medications were intensified from 1 to 2 drugs 32% (IQR 26-37%) of the time when SBP was 140-159 mmHg, 48% (IQR 30-57%) when SBP was 160-179 mmHg, and 70% (63-79%) when SBP was ≥180 mmHg.[13] |

| Parameter (under intervention conditions) | Description | Sampled value | Justification |
|---|---|---|---|
| | medication. **CCC and CHW policies.** | | |
| *prob_intensify_2_3* | For a person on two antihypertensive medications with current SBP ≥140, probability of adding a third antihypertensive medication. **CCC and CHW policies.** | 0.02: 33%, 0.03: 33%, 0.04: 33%<br><br>Probability 1.5-fold higher if SBP 160-179 mmHg and 2-fold higher if SBP ≥180 mmHg | In the SEARCH median intervention arm community (where medications were available), medications were intensified from 2 to 3 drugs 3% (IQR 3-7%) of the time when SBP was ≥140 mmHg.[30] |
| *htn_retention_patt* | Pattern of visit retention under intervention conditions<br><br>If on antihypertensives and due for a visit (or if new diagnosis after initial visit with SBP 140-159 and given lifestyle counselling), the probability of return for subsequent visit.<br><br>Probability of return increases with each consecutively attended visit.<br><br>Persons who do not have a visit fall out of care. If they subsequently re-enter care, the visit number restarts at 1. **CCC and CHW policies.** | 33% pattern 1:<br>*prob_visit_htn_v1*: 0.65<br>*prob_visit_htn_v2*: 0.75<br>*prob_visit_htn_v3*: 0.85<br>*prob_visit_htn_v4*: 0.90<br>*prob_visit_htn_v5*: 0.95<br><br>33% pattern 2:<br>*prob_visit_htn_v1*: 0.70<br>*prob_visit_htn_v2*: 0.80<br>*prob_visit_htn_v3*: 0.90<br>*prob_visit_htn_v4*: 0.95<br>*prob_visit_htn_v5*: 0.95<br><br>33% pattern 3:<br>*prob_visit_htn_v1*: 0.75<br>prob_visit_htn_v2: 0.85<br>prob_visit_htn_v3: 0.95<br>prob_visit_htn_v4: 0.95<br>prob_visit_htn_v5: 0.95 | Probability of retention in hypertension care based on intervention arm of SEARCH study (patient-centred care intervention) demonstrating 48% with consistent care engagement (≥2 visits per year) over 3 years.[13]<br><br>Assuming a 5% rate of re-engagement in care in each quarter, modelled visit probabilities will yield the following proportion retained in care after 3 years (assuming 2 quarterly visits, then twice-yearly visits): 49% retained (range 39-55%) |

#### Table S9. Cost Assumptions

Table S9 provides cost assumptions for listed intervention components evaluated in the model. We explore variability in cost estimates using 50% and 200% costing scenarios. All costs listed are modelled in 2024 US Dollars.

| Parameter | Description | Sampled value (in US dollars) | Justification |
|---|---|---|---|
| cost_screen_comm | Per-patient cost of community-based screening by a community health worker (CHW), conducted annually | $3 | Based on a pilot study in SEARCH of community-based hypertension screening in Kenya and Uganda (n=200), community health workers received a stipend of $60 USD per month (including phone airtime) and screened an average of 6 adults ≥40 years of age per day (range 4-8 between communities).<br><br>We assume a higher stipend of $80 USD per month in 2024, which is the approximate median minimum wage across 24 countries and 25 sub-settings within Africa where minimum wages have been established and were updated in the past 5 years. As described above, we also explore variability in screening costs using 50% and 200% costing scenarios to explore cost-effectiveness in settings where screening costs were different than the base-case assumptions. This variability encompasses the variability in minimum wages set across countries. See Table S10 for details on minimum wages across countries in continental east, southern, central, and west Africa (including Madagascar but excluding small island nations, e.g. Seychelles, where economic conditions may differ more substantially).<br><br>Assuming that all community health workers in a community participate, that each community health worker oversees approximately 100 households (total 500 people) and that approximately 15% are over the age of 40, CHWs would screen approximately 75 individuals annually. Screening could likely be completed in a single month, but conservatively we assume it will occur over 2 months with 1-2 people screened per day (thus allowing for other CHW activities to continue uninterrupted and to avoid over-burdening clinics). We allocate 50% of CHW costs during that 2-month period to the CHW hypertension screening intervention (1 person-month/year). This corresponds to an approximate CHW cost of $1.07 USD per person screened. Other inputs include one blood pressure cuff and battery supply for every village (2-3 CHWs/village, $120 USD every 2 years). Annual training costs are assumed to be $1000 USD per 20 CHWs for per diem costs for four nurses to provide hands-on training on blood pressure screening, facilities, and facilitation costs in each community (ratio 1 nurse per 5 CHWs for training. We assume that existing CHW supervisory structures will be leveraged and that 50% FTE of these costs are allocated to hypertension screening during the two-month screening period each year (supervisor annual salary $8000 USD for supervision of 20 CHWs, allocating 50% monthly salary for 2 months to 1500 screened per community contributes $0.44 per person to screening costs). We add 20% to account for additional overhead costs, yielding a total estimated cost of $3 USD per person screened. |
| cost_htn_visit | Cost per hypertension clinic visit | If SBP <140: $5<br>If SBP ≥140: $10 | We assume hypertension costs are $10 per period when blood pressure is uncontrolled (SBP ≥140) and $5 per period when blood pressure is controlled. This cost structure is analogous to the visit costs assumed for HIV care in the HIV Synthesis model with higher costs for more complex care. With chronic care clinic implementation, we further assume a cost of $0 for hypertension care when a patient is also seen for HIV. We also separately model 50% and 200% cost scenarios to capture variability.<br><br>Based on data from SEARCH, integrated patient-centred hypertension care cost $2.25 USD per visit for individuals without HIV, adjusted to 2024 dollars (annual non-medication cost of $8.32 USD in 2024 and average of 3.7 visits/year). Cost was $0 for those with HIV because co-management of HIV and hypertension did not add any additional staff time or other resources to the visit.[57] |

| | | | |
|---|---|---|---|
| | | | Subramanian et al found that quarterly patient costs for hypertension care at public facilities in Kenya inclusive of medications were $6.41 for patients on 1 drug, $16.81 for patients on 2 drugs, and $20.30 for patients on 3 drugs.[58] Removing generic drug costs[59] (see below) and converting to 2023 dollars, this yields an estimated per visit cost of $5 for those on 1 drug and $14 for those on 2-3 drugs.<br><br>In the INTE-AFRICA study, costs are reported in two different ways. Total cost of hypertension care exclusive of medications and attributing administrative costs of HIV to hypertension was $14.96 per month ($44.88 quarterly) for people with hypertension alone.[60] In contrast, health system costs of a hypertension visit based on time-in-motion data was $4.31 per visit, adjusted to 2024 US dollars. There was no additive cost for hypertension/HIV visits compared to HIV alone. |
| Annual training cost for chronic care clinics (fixed) | Cost associated with training primary health centres to implement integrated hypertension care | $960,000 per 10 million adult population | We assume that integrated hypertension treatment would require additional training for clinic staff, and that this training would leverage existing continuing medical education mechanisms. We assume a cost of $100 per day for per diem trainer costs and $100 cost training facilitation (rent, supplies) with two training days per year plus 20% overhead. Assumed annual training cost per clinic is therefore $480 USD. Assuming that primary health centres serve a population of approximately 5000 adults, annual implementation costs would scale to $960,000 annually across 2000 health facilities serving 10 million adults. |
| cost_htn_drug1 | Quarterly cost of 1st antihypertensive medication (in USD) | $1.5 | Based on data from the Resolve to Save Lives 2022 report on hypertension treatment costs in low and middle income countries.[59] We multiplied generic medication costs for a 90-day supply by 1.3 to account for costs associated with procurement and quality monitoring.<br><br>Medication costs from the Resolve to Save Lives report compared to model inputs (price per patient for 90-day supply, in USD):<br>• Amlodipine 10mg<br>o Generic: $1.15<br>o South Africa public sector: $1.07<br>o Nigeria public sector: $2.16<br>o Model (use as 1st or 2nd line drug): $1.50<br>• Hydrochlorothiazide 25mg<br>o Generic: $1.12<br>o South Africa public sector: $0.96<br>o Nigeria public sector: $1.53<br>o Model (use as 1st or 2nd line drug): $1.50<br>• Losartan 100mg<br>o Generic: $2.03<br>o South Africa public sector: $3.78<br>o Nigeria public sector: $14.49<br>o Model (3rd line drug): $3.00<br>• Telmisartan 80mg<br>o Generic: $2.30<br>o South Africa public sector: $8.19<br>o Nigeria public sector: n/a<br>o Model (3rd line drug): $3.00 |
| cost_htn_drug2 | Quarterly cost of 2nd antihypertensive medication (in USD) | $1.5 | |
| cost_htn_drug3 | Quarterly cost of 3rd antihypertensive medication (in USD) | $3 | |
| cost_ihd_tx | Cost of hospitalization for acute MI and associated treatment | $2,270 | Derived from public sector costs from Subramanian et al, adjusted for inflation to 2024 US dollars.[58] Cost only incurred if acute treatment is accessed (defined by prob_ihd_tx). |

| | | | |
|---|---|---|---|
| cost_cva_tx | Cost of hospitalization for acute stroke and associated treatment | $2,420 | Derived from public sector costs from Subramanian et al, adjusted for inflation to 2024 US dollars.[58] Cost only incurred if acute treatment is accessed (defined by prob_cva_tx). |
| cost_ihd_tx_lowqual | Cost of hospitalization for acute MI associated with low-quality or delayed medical care | $ 567.5 / $1,135 / $2,270 | Based on assumption that low-quality or delayed medical care may be less extensive or at a lower-level facility that lacks advanced therapy, and therefore less costly. We sampled from three values, evaluating conditions where low-quality care is 25%, 50%, and 100% of the cost of effective care. |
| cost_cva_tx_lowqual | Cost of hospitalization for acute stroke associated with low-quality or delayed medical care | $605 / $1,210 / $2,420 | Based on assumption that low-quality or delayed medical care may be less extensive or at a lower-level facility that lacks advanced therapy, and therefore less costly. We sampled from three values, evaluating conditions where low-quality care is 25%, 50%, and 100% of the cost of effective care. |

#### Table S10. Minimum Wage Policies Across Countries and Settings in Africa

Table S10 provides information on minimum wages set in countries across continental Africa (including Madagascar). We excluded countries with minimum wage policies that were last updated >5 years ago and excluded small island nations due to smaller population size and economic conditions that may differ more substantially from larger nations across the region. Source: wageindicator.org (accessed 12-Feb-2024)

**Median minimum wage:** $76 USD/month
**10$^{th}$ percentile:** $33 USD/month; **90$^{th}$ percentile:** $134 USD/month
**Range:** $5 USD/month (Sudan) to $240 USD/month (South Africa)

| Country | Currency code | Currency Name | Exchange rate to USD (1/1/2024) | Monthly minimum wage (local currency) | Monthly minimum wage (US Dollars) | Year minimum wage policy updated |
|---|---|---|---|---|---|---|
| Angola | AOA | Angolan Kwanza | 0.00120657 | 32181 | 38.83 | 2022 |
| Benin | XOF | West African CFA franc | 0.00168388 | 52000 | 87.56 | 2023 |
| Botswana | BWP | Botswanan Pula | 0.07439049 | 1084 | 80.64 | 2022 |
| Burkina Faso | XOF | West African CFA franc | 0.00168388 | 45000 | 75.77 | 2023 |
| Cameroon | XAF | Central African CFA franc | 0.00168388 | 41875 | 70.51 | 2023 |
| Central African Republic | XAF | Central African CFA franc | 0.00168388 | 35000 | 58.94 | 2018 |
| Cote d'Ivoire | XOF | West African CFA franc | 0.00168388 | 75000 | 126.29 | 2023 |
| Djibouti | DJF | Djiboutian franc | 0.00562682 | 35000 | 196.94 | 2018 |
| Democratic Republic of the Congo | CDF | Congolese franc | 0.00037369 | 153174 | 57.24 | 2019 |
| The Gambia | GMD | Gambian dalasi | 0.01485332 | 1083 | 16.09 | 2018 |
| Ghana | GHS | Ghanaian Cedi | 0.08334340 | 1572 | 131.02 | 2024 |
| Guinea | GNF | Guinean Franc | 0.00011598 | 550000 | 63.79 | 2022 |
| Kenya (Nairobi) | KES | Kenyan shilling | 0.00635163 | 15202 | 96.56 | 2022 |
| Kenya (rural) | KES | Kenyan shilling | 0.00635163 | 8110 | 51.51 | 2022 |
| Lesotho | LSL | Lesotho Loti | 0.05396725 | 2053 | 110.79 | 2022 |
| Madagascar | MGA | Malagasy ariary | 0.00021641 | 184653 | 39.96 | 2022 |
| Malawi | MWK | Malawian kwacha | 0.00059238 | 50000 | 29.62 | 2021 |
| Mozambique | MZN | Mozambican metical | 0.01581029 | 8574 | 135.56 | 2024 |
| Namibia | NAD | Namibian dollar | 0.05396621 | 1564 | 84.4 | 2018 |
| Nigeria | NGN | Nigerian naira | 0.00111222 | 65000 | 72.29 | 2023 |
| South Africa | ZAR | South African Rand | 0.05459886 | 4403 | 240.4 | 2023 |
| Sudan | SDG | Sudanese pound | 0.00166389 | 3000 | 4.99 | 2020 |
| Tanzania | TZS | Tanzanian shilling | 0.00039618 | 195000 | 77.26 | 2023 |
| Togo | XOF | West African CFA franc | 0.00168388 | 52000 | 87.56 | 2023 |
| Zambia | ZMW | Zambian kwacha | 0.03874934 | 1699 | 65.84 | 2018 |

### Section 4. Comparison of model output in 2015 to data from the literature

The purpose of this section is to compare baseline model outputs to available data from the literature.

Based on 3000 model runs

Model outputs in 2015 reflect output averaged across 3-month model steps from 2013 through <2017 (i.e. 2015 +/- 2 years). This wider time window was selected to reduce the effect of stochastic variation in individual setting-scenarios.

#### Definitions

SOC = Standard of Care (model output estimating current conditions in 2015, the mid-point of currently available data)
GBD = Global Burden of Disease data

#### Data Sources

Hypertension prevalence, diagnosis, treatment, and control using STEPS survey data from 15 countries in Africa (2005-2015; Geldsetzer Lancet 2019).[27]

Systolic blood pressure for men and women based on 2015 Global Burden of Disease survey data (Forouzanfar et al JAMA 2017).[19]

Global Burden of Disease Collaborative Network. Global Burden of Disease Study 2019 (GBD 2019) Results. Seattle, United States: Institute for Health Metrics and Evaluation (IHME), 2020. Available from https://vizhub.healthdata.org/gbd-results/.[17]

#### SBP Increase with Age

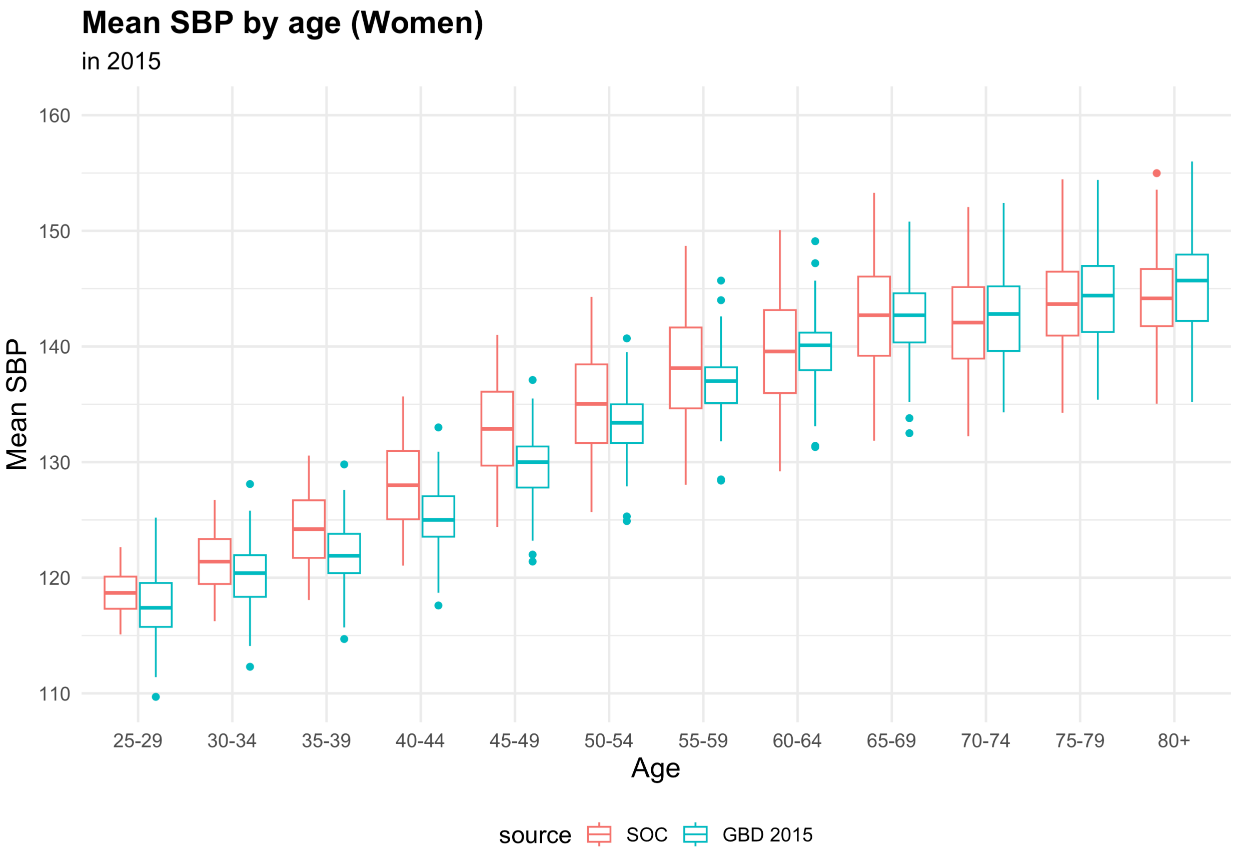

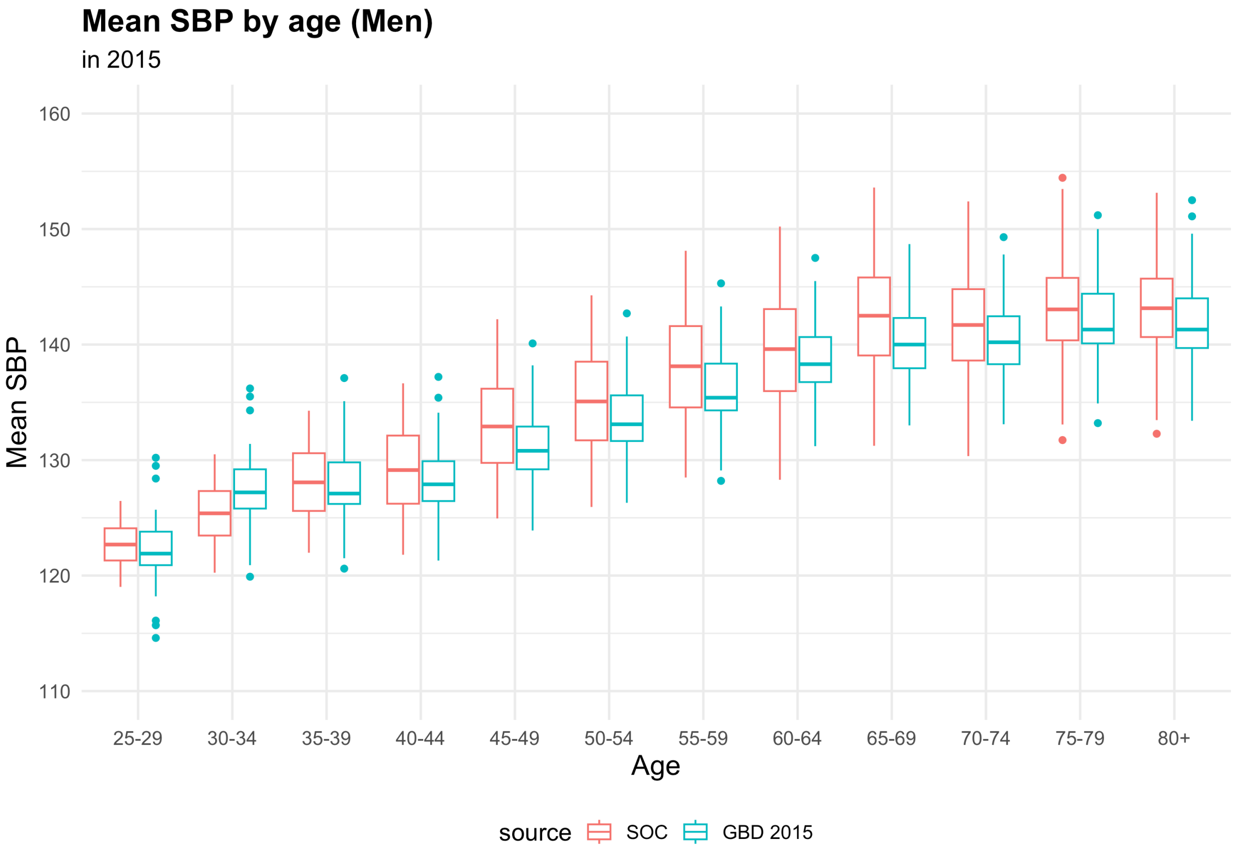

#### Hypertension Prevalence

Hypertension defined as true SBP >=140 mmHg, regardless of whether measured.

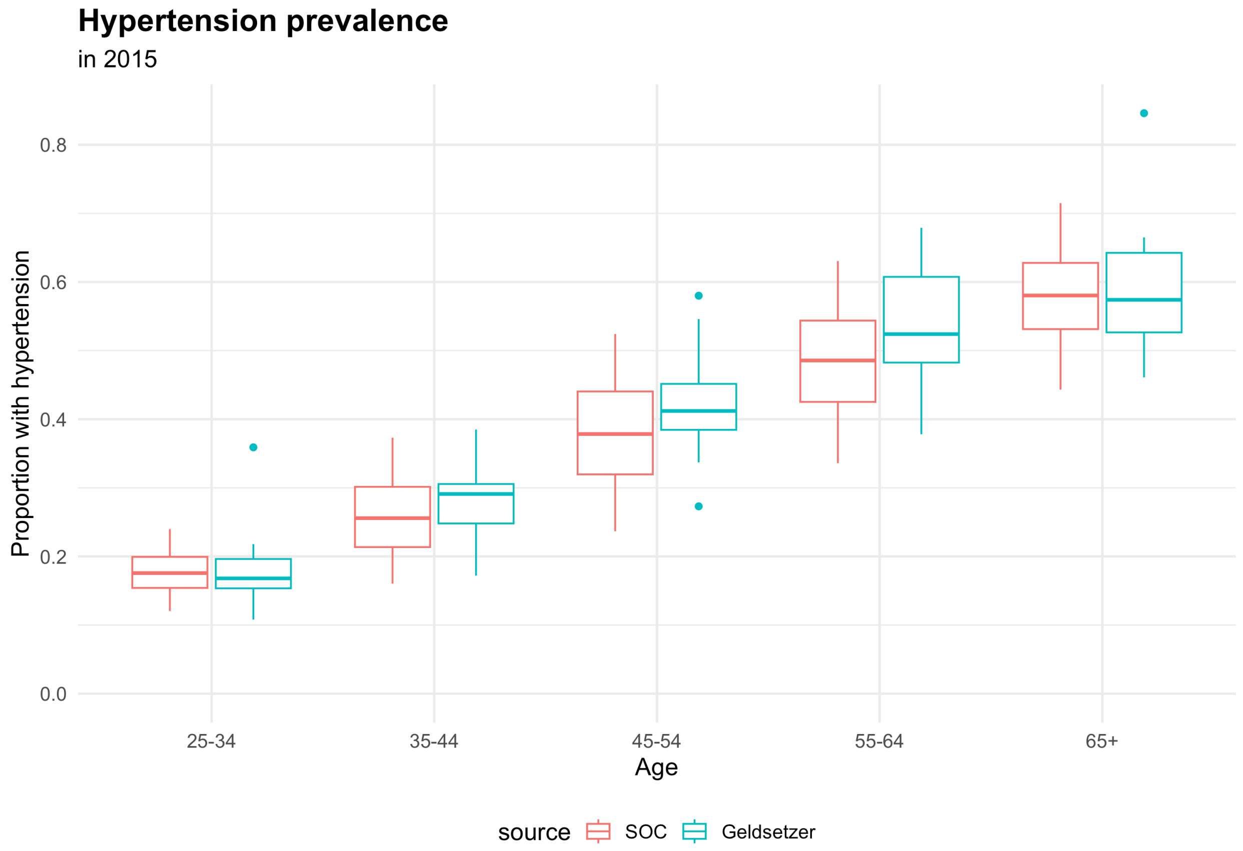

#### Hypertension Diagnosis

- Measured SBP value if measured: true SBP + random error with SD 10 mmHg
- Hypertension diagnosis made if any of the following:
    1. measured SBP >=140 mmHg on two consecutive clinic visits
    2. measured SBP >=160 mmHg once
    3. started on antihypertensive medication (see Hypertension Treatment section below)

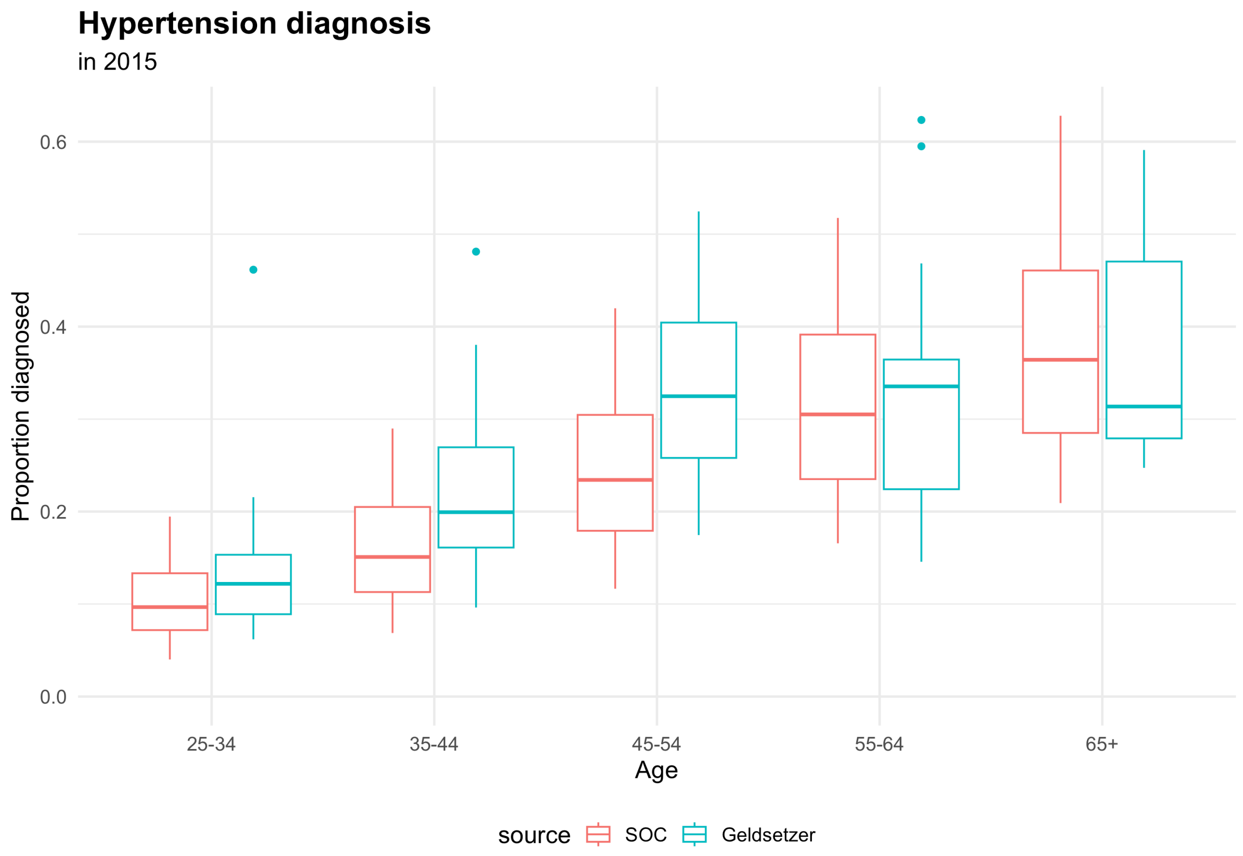

### Hypertension Treatment

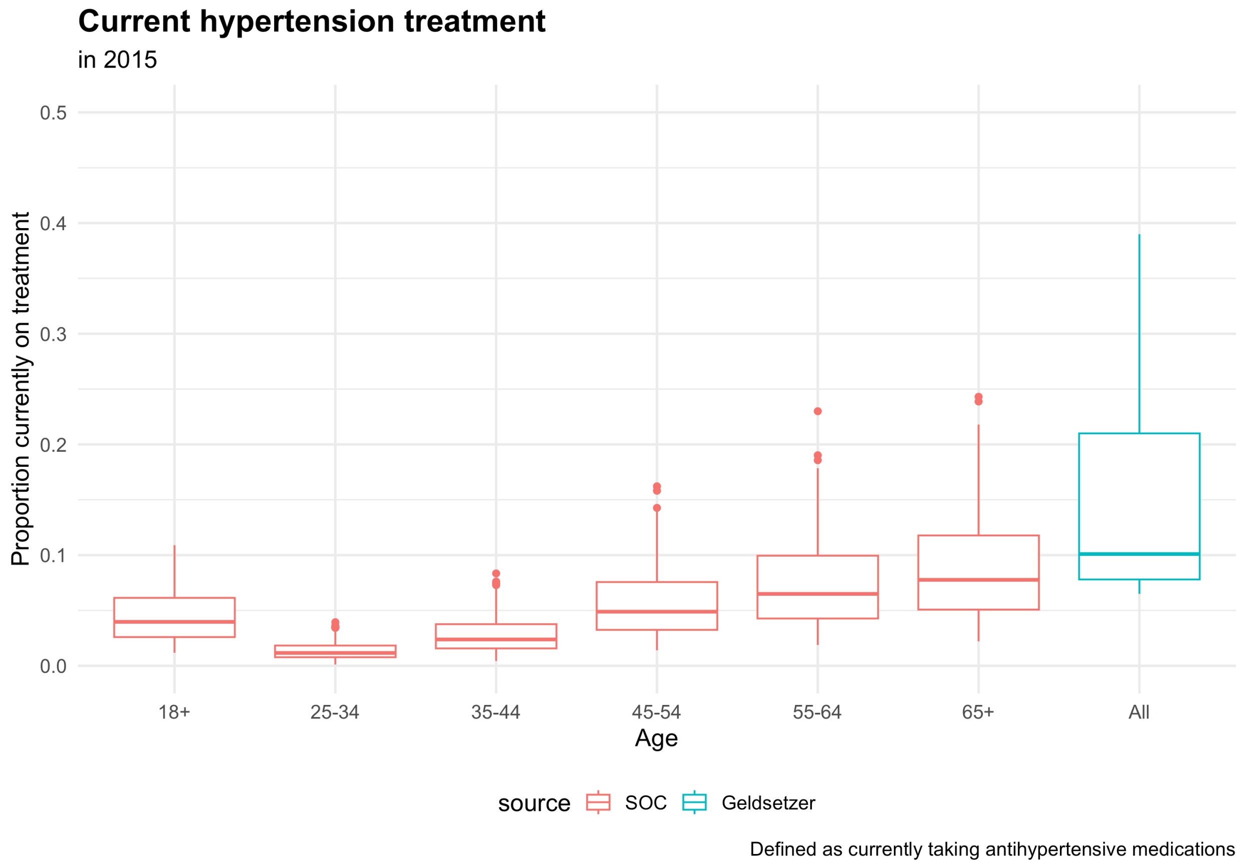

**Note:** The model considers treatment to be on medications and adherent to those medications. Real-world data is based on self-report of treatment and may include some who report taking medications but are non-adherent.

Geldsetzer Lancet 2019 includes country-specific data on 13 countries in Africa with data on medication treatment and reports an overall prevalence of pharmacologic hypertension treatment of 13.0% (95% CI 12.0-14.2%) when weighting countries by population size.

| Country | Hypertension treatment (95% CI) |
|---|---|
| Tanzania | 6% (4.9-8.4) |
| Mozambique | 7% (4.4-9.6) |
| Uganda | 8% (5.8-9.5) |
| Kenya | 8% (5.8-10.1) |
| Togo | 8% (5.7-11.1) |
| Burkina Faso | 9% (6.9-11.9) |
| Ghana | 10% (7.3-13.4) |
| Benin | 15% (12.6-17.6) |
| Liberia | 16% (12.6-19.4) |
| Swaziland | 21% (17.5-24.8) |
| South Africa | 28% (24.4-30.7) |
| Lesotho | 37% (33.6-40.9) |
| Namibia | 39% (35.9-42.1) |

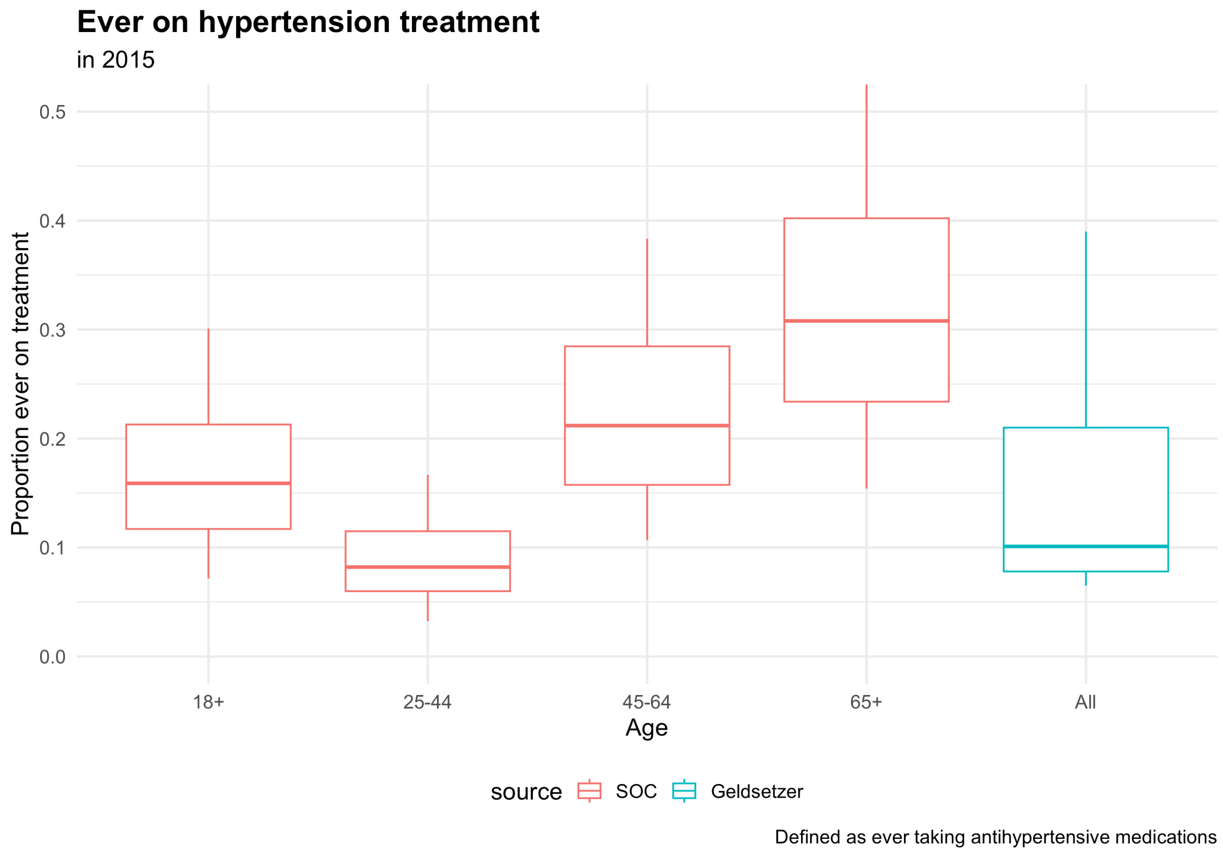

#### Hypertension Control

- Control defined as current true SBP <140 among persons with maximum SBP >=140 (pre-treatment)

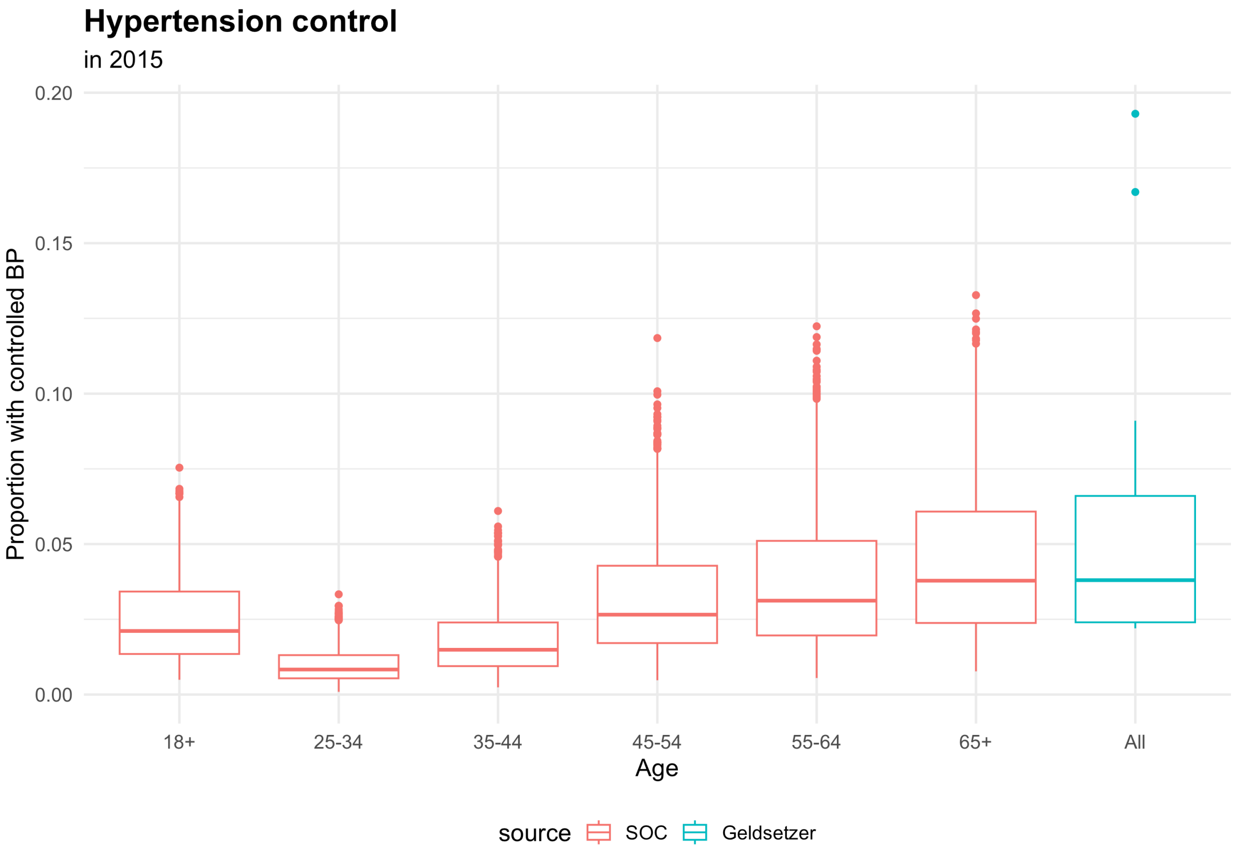

Geldsetzer Lancet 2019 includes country-specific data on 13 countries in Africa with data on hypertension control and reports an overall prevalence of hypertension control (<140/90 mmHg) of 4.3% (95% CI 3.6-4.9%).

| Country | Hypertension control (95% CI) | |
|---|---|---|
| Ghana | 2% | (1.2-3.5) |
| Tanzania | 2% | (1.4-3.1) |
| Uganda | 2% | (1.5-3.4) |
| Mozambique | 2% | (1.4-3.7) |
| Kenya | 3% | (1.3-5.0) |
| Togo | 3% | (1.9-5.4) |
| Burkina Faso | 4% | (2.2-5.8) |
| Benin | 4% | (2.7-5.9) |
| Liberia | 4% | (2.7-6.0) |
| Swaziland | 7% | (4.1-9.7) |
| South Africa | 9% | (6.9-11.5) |
| Namibia | 17% | (14.4-19.1) |
| Lesotho | 19% | (16.4-22.3) |

#### CVD Events

Model output (depicted as SOC in the figures) compared to CVD incidence and prevalence data from the Global Burden of Disease (GBD) Project grouped by regions (western, eastern, southern, and central Africa).[17]

##### Incidence of Ischemic Heart Disease

Incidence of ischemic heart disease (angina and myocardial infarction(MI)). We assume that angina/silent MI are under-ascertained, thus we compare incidence only of moderate/severe MI to available incidence data.

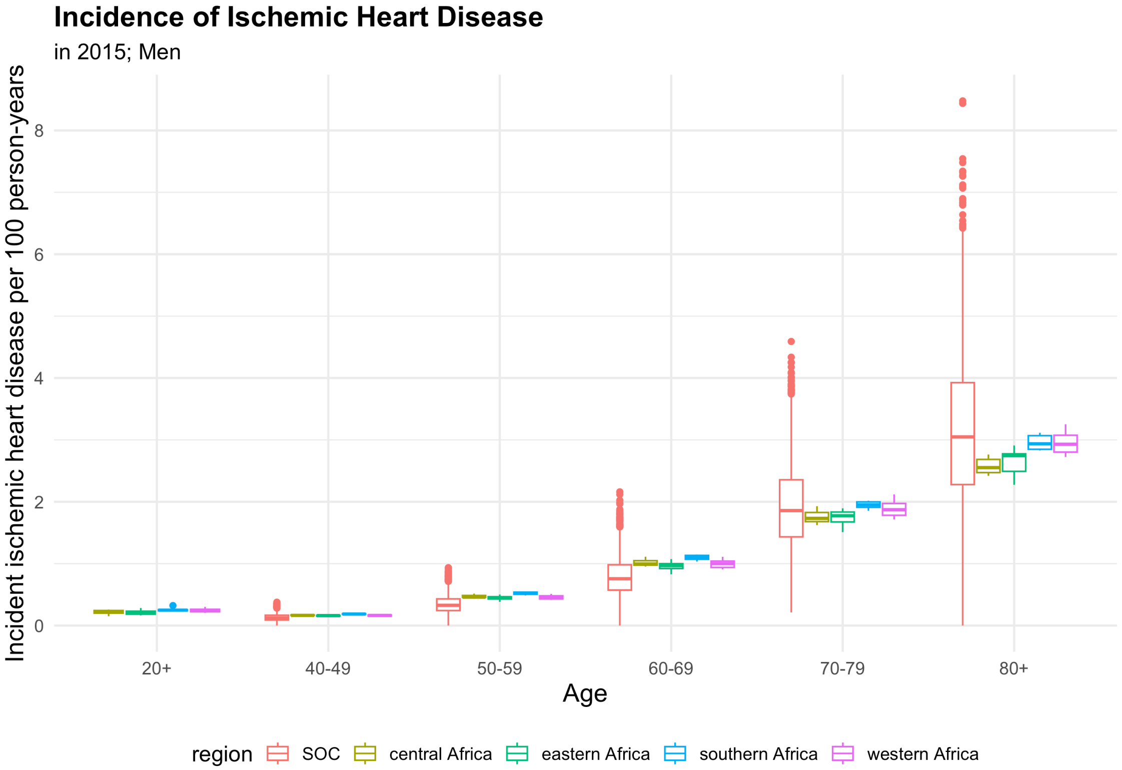

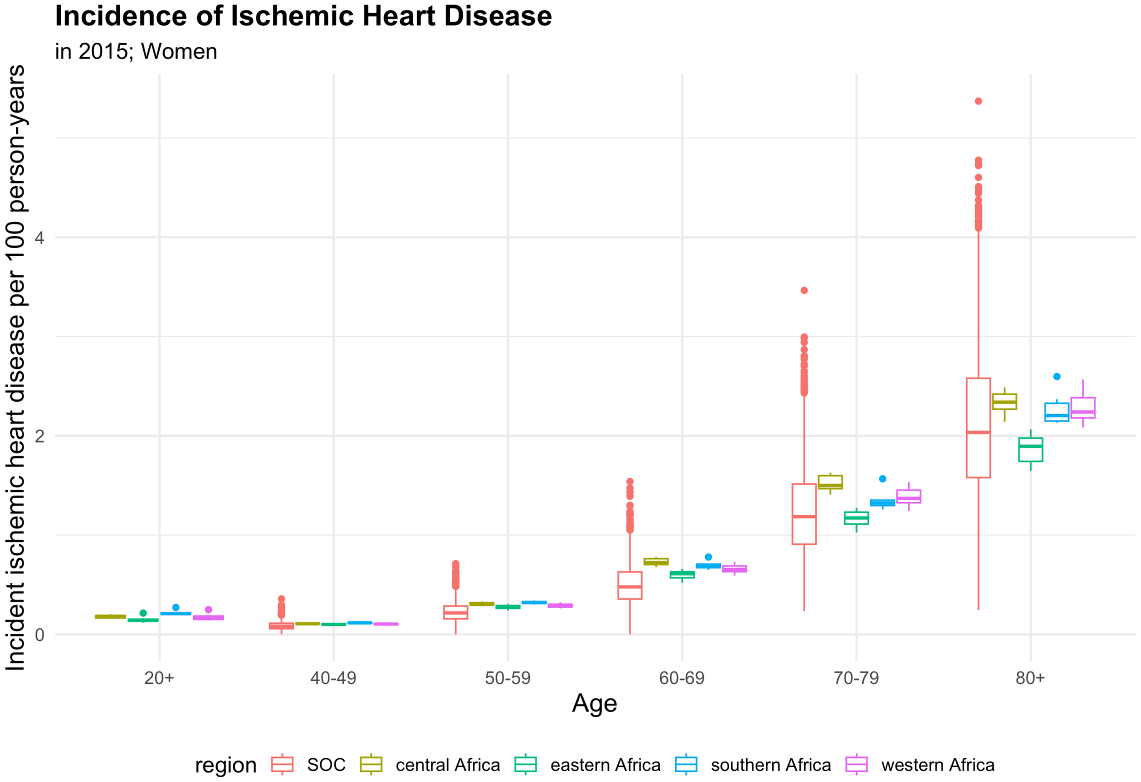

#### Prevalence of Ischemic Heart Disease

Defined as any prior history of ischemic heart disease among living adults.

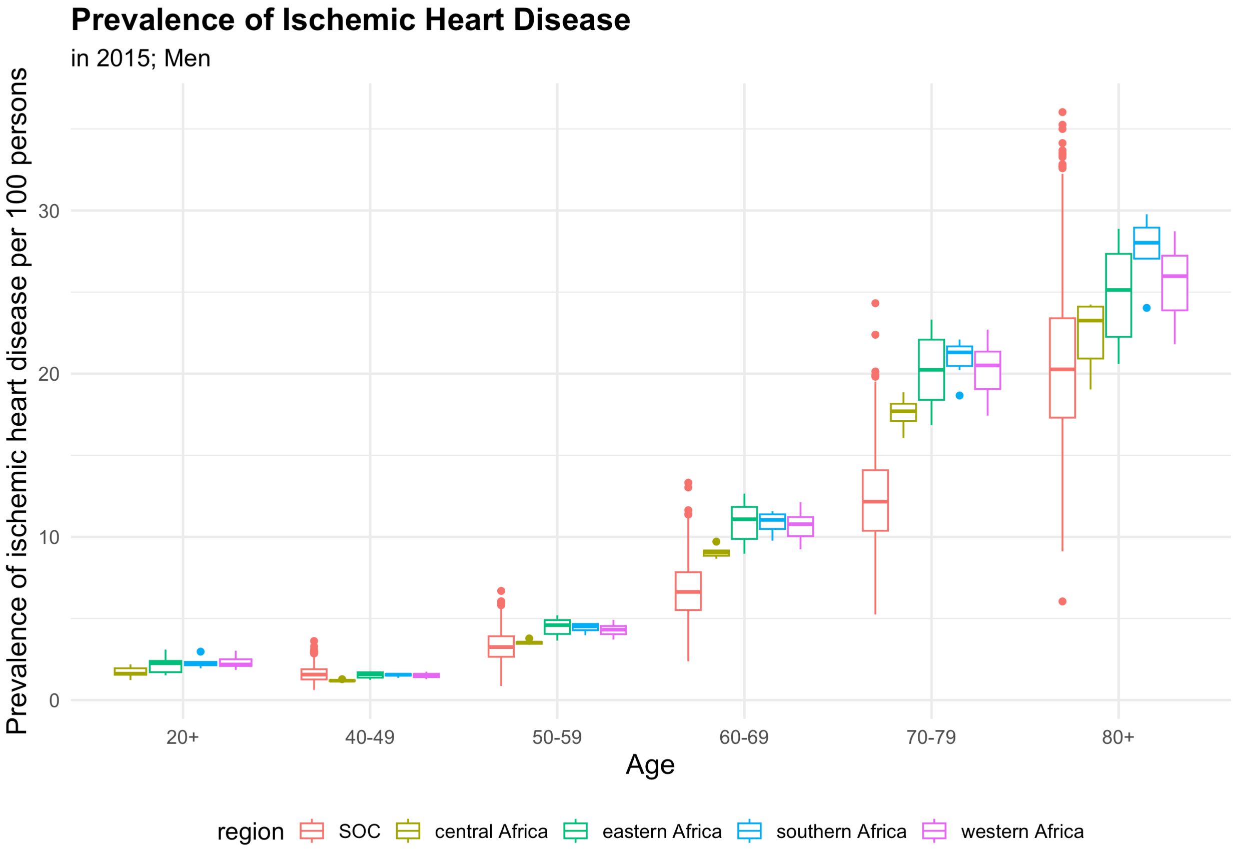

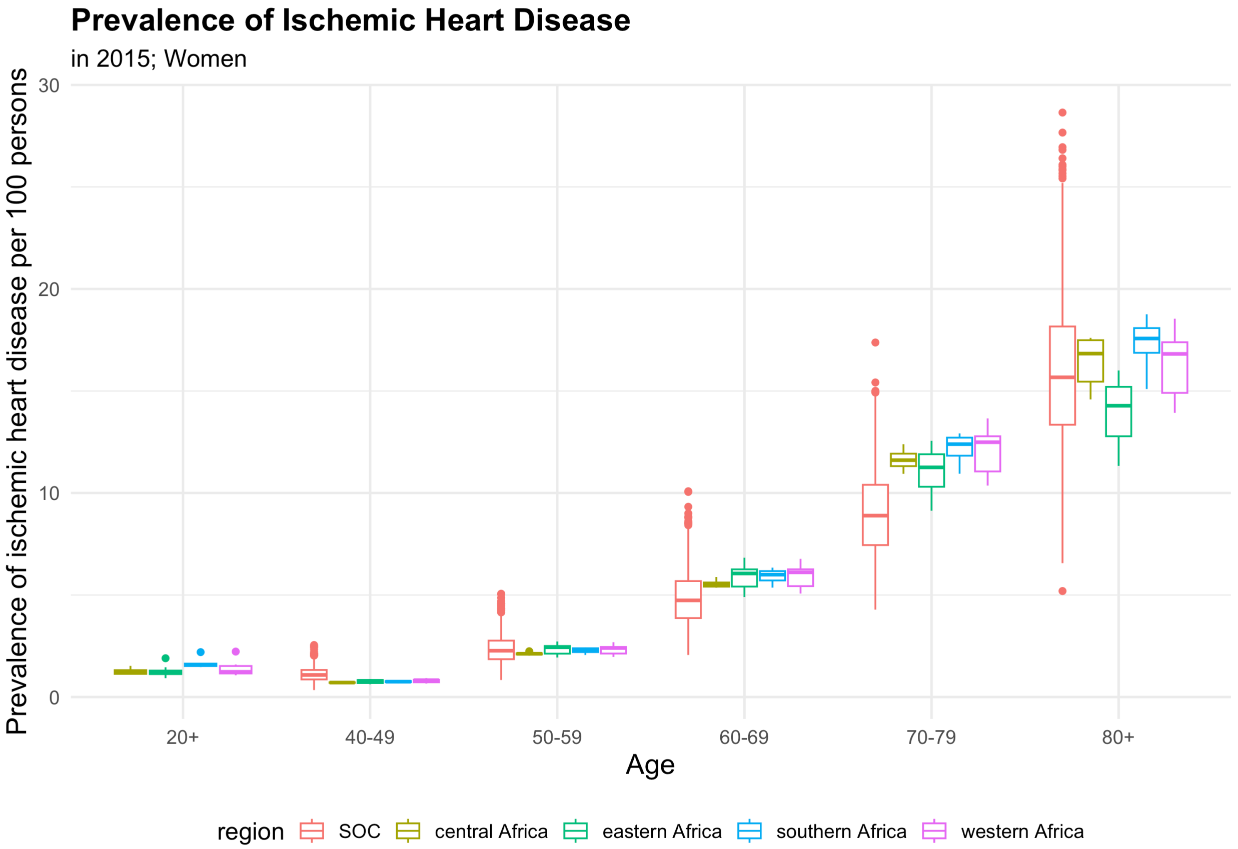

#### Incidence of Stroke

We assume that mild strokes under-ascertained and thus compare incidence only of moderate/severe stroke to available incidence data.

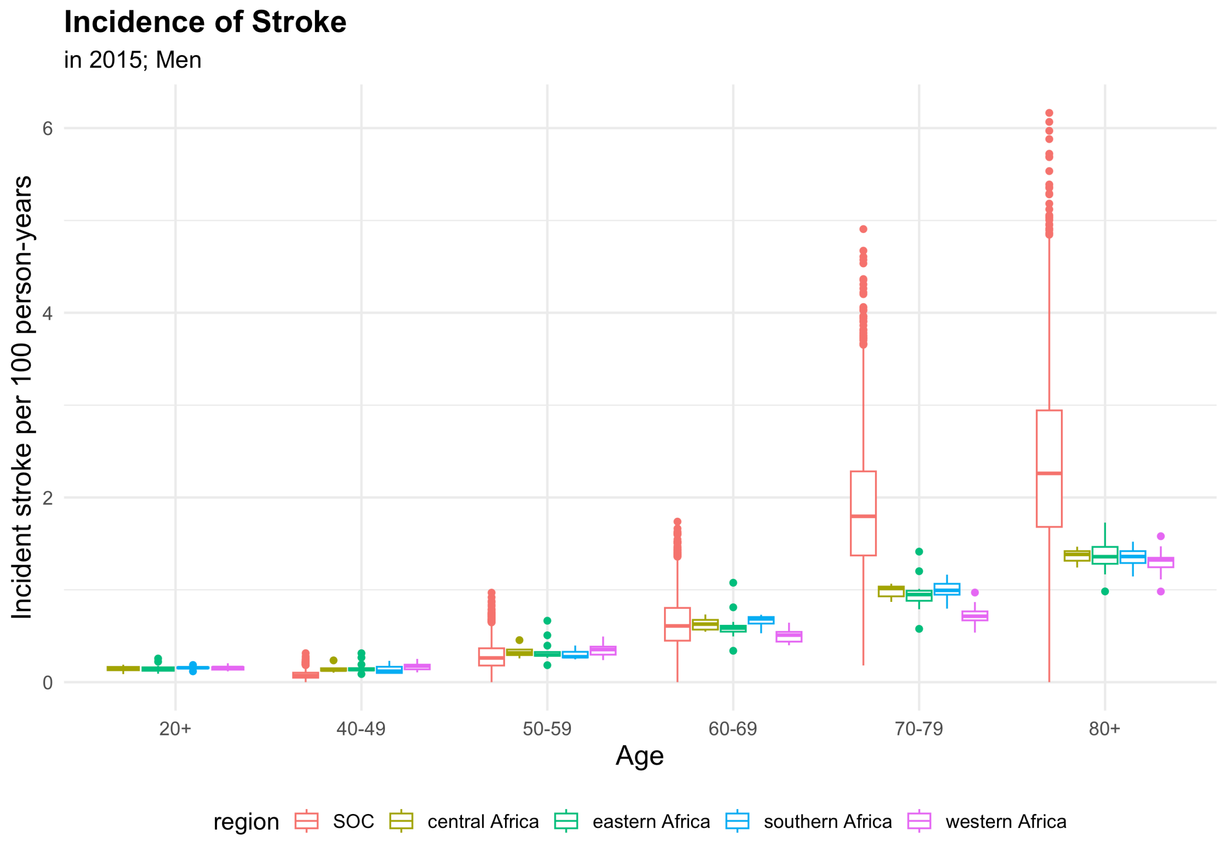

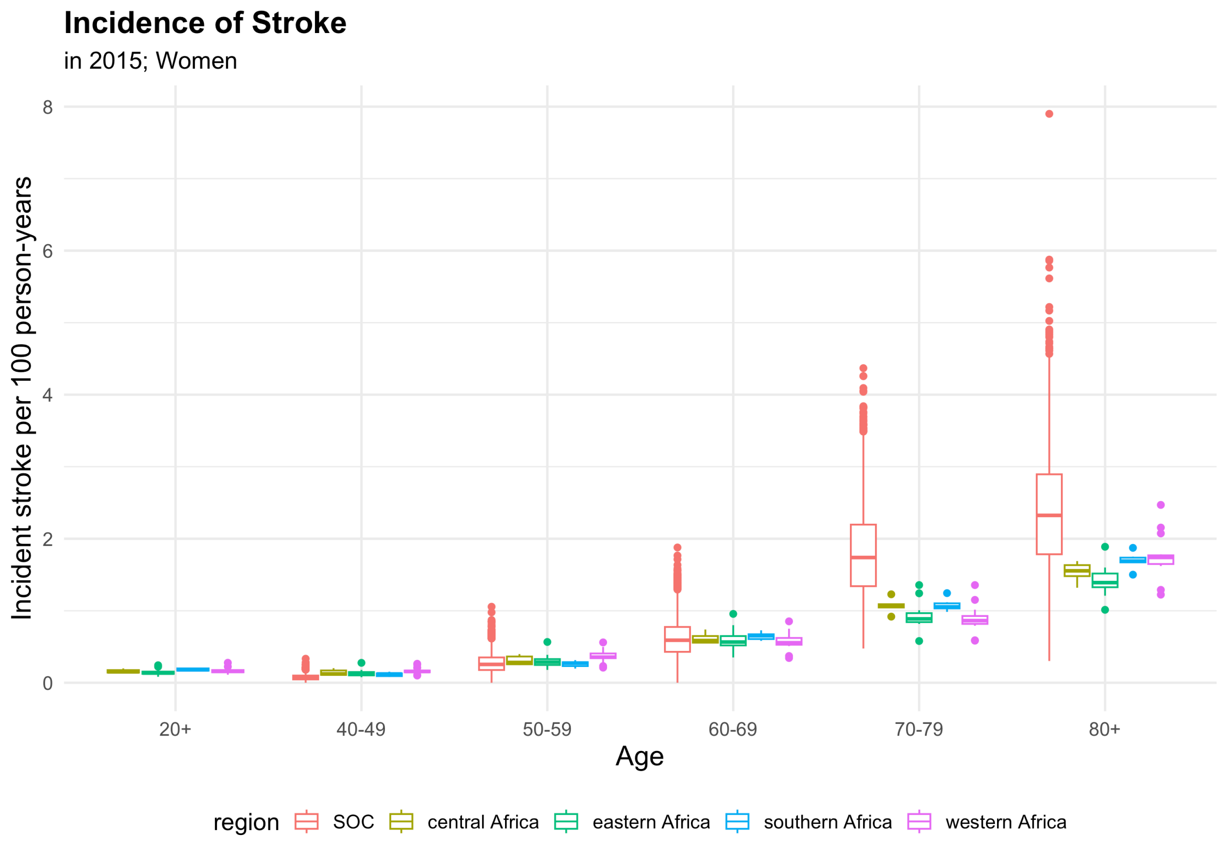

#### Prevalence of Stroke

Defined as any prior history of stroke among living adults.

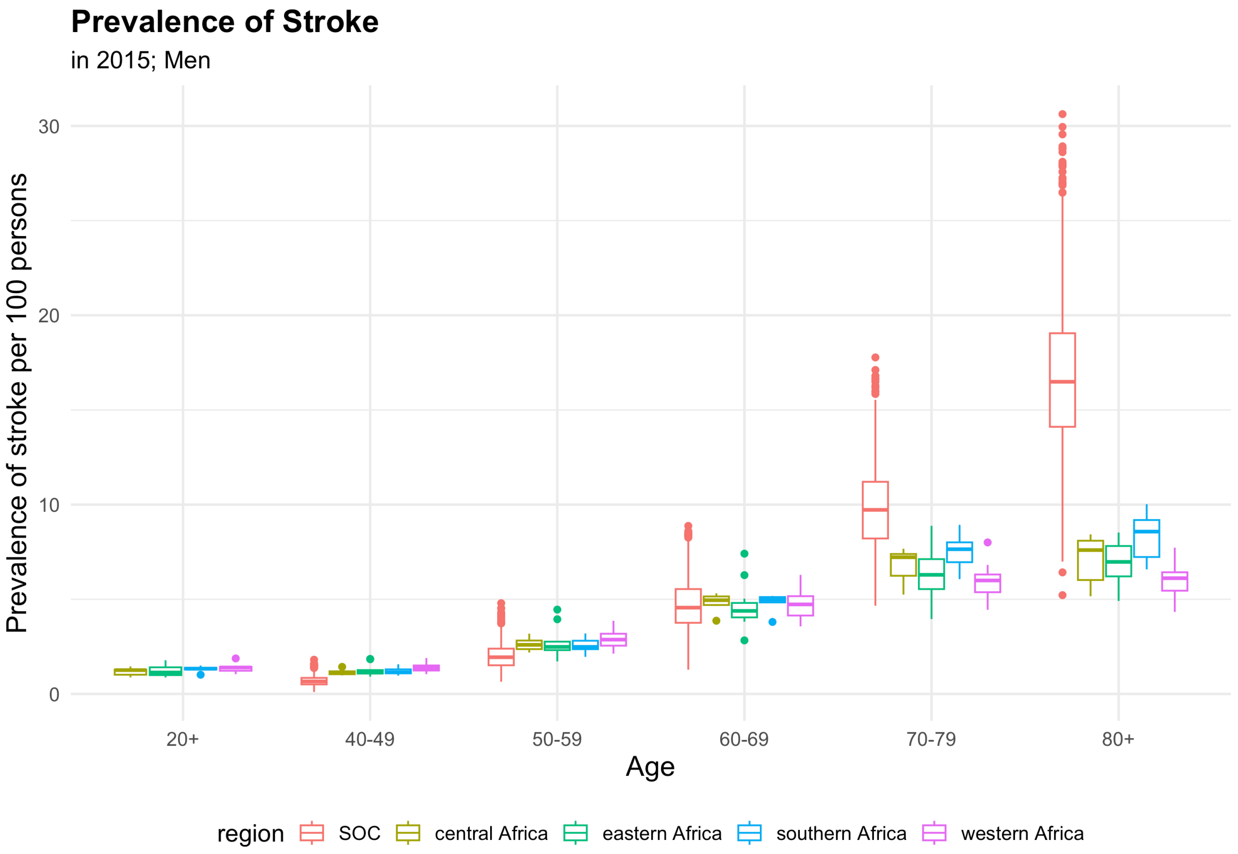

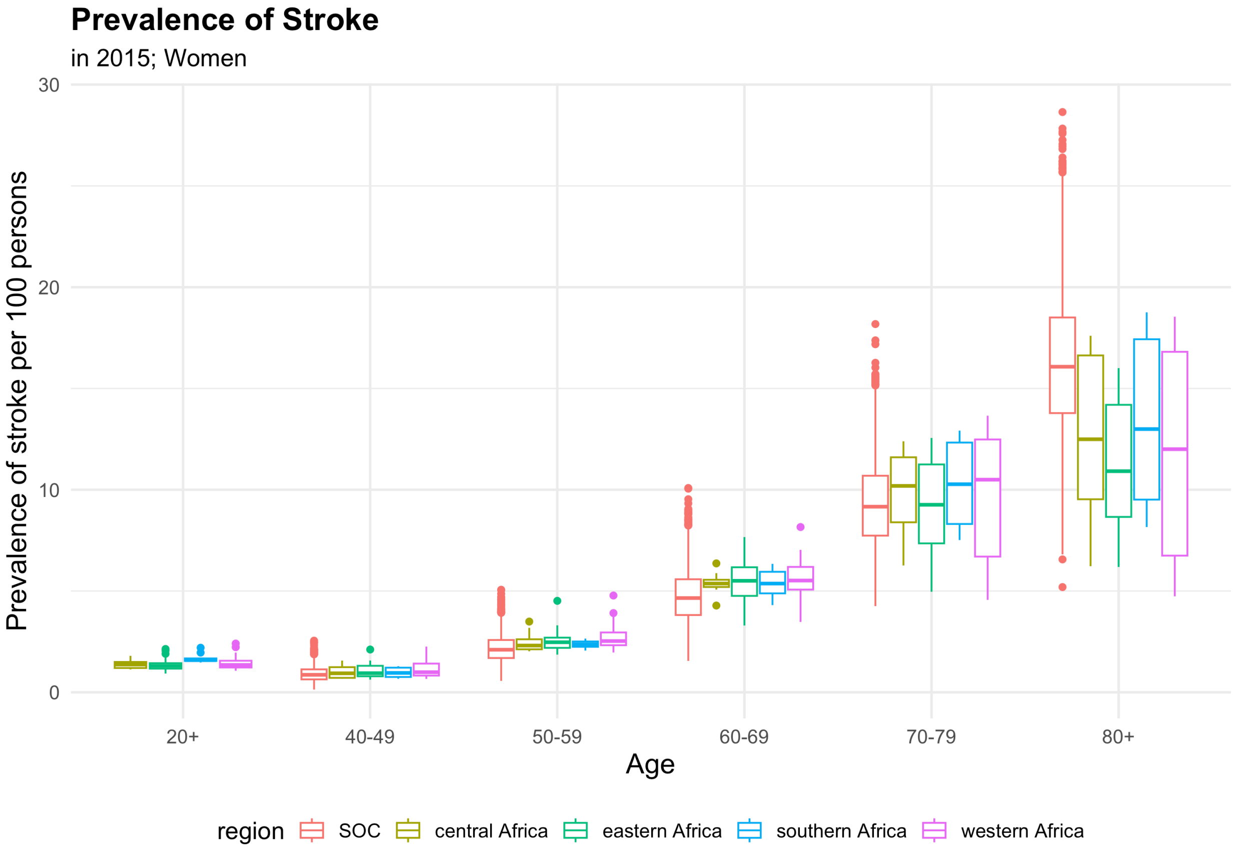

#### CVD Mortality

Mortality resulting from ischemic heart disease or stroke.

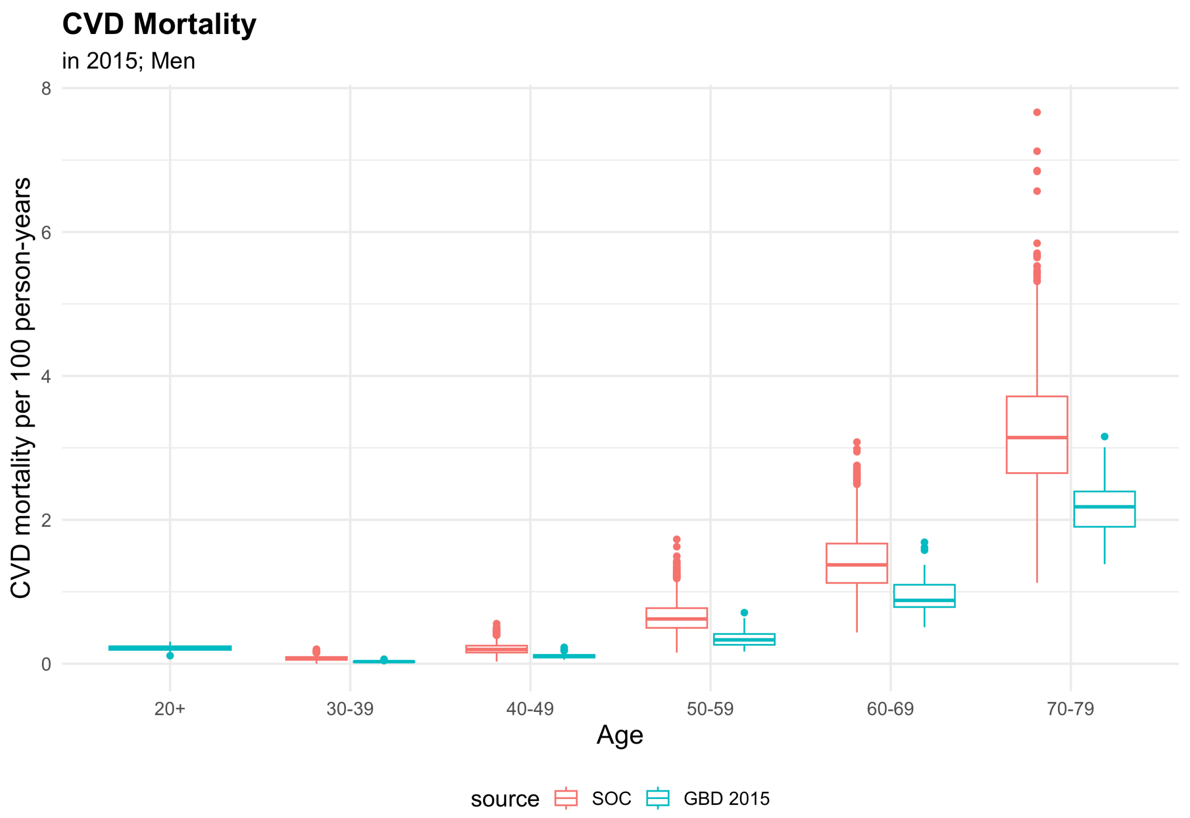

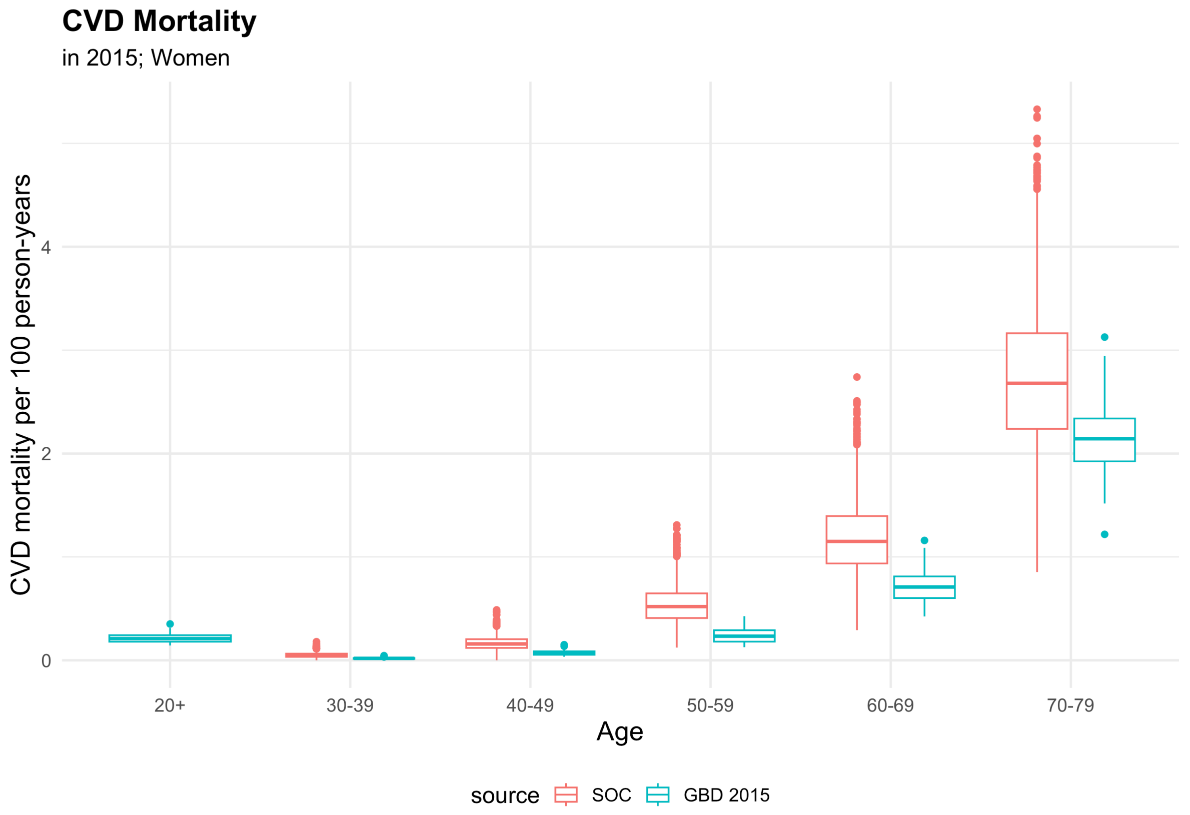

### Section 5: Supplemental Results

Hypertension care cascade for CCC and CHW policy scenarios compared to standard of care. Model estimates averaged over 2024-2074.

#### Hypertension Prevalence

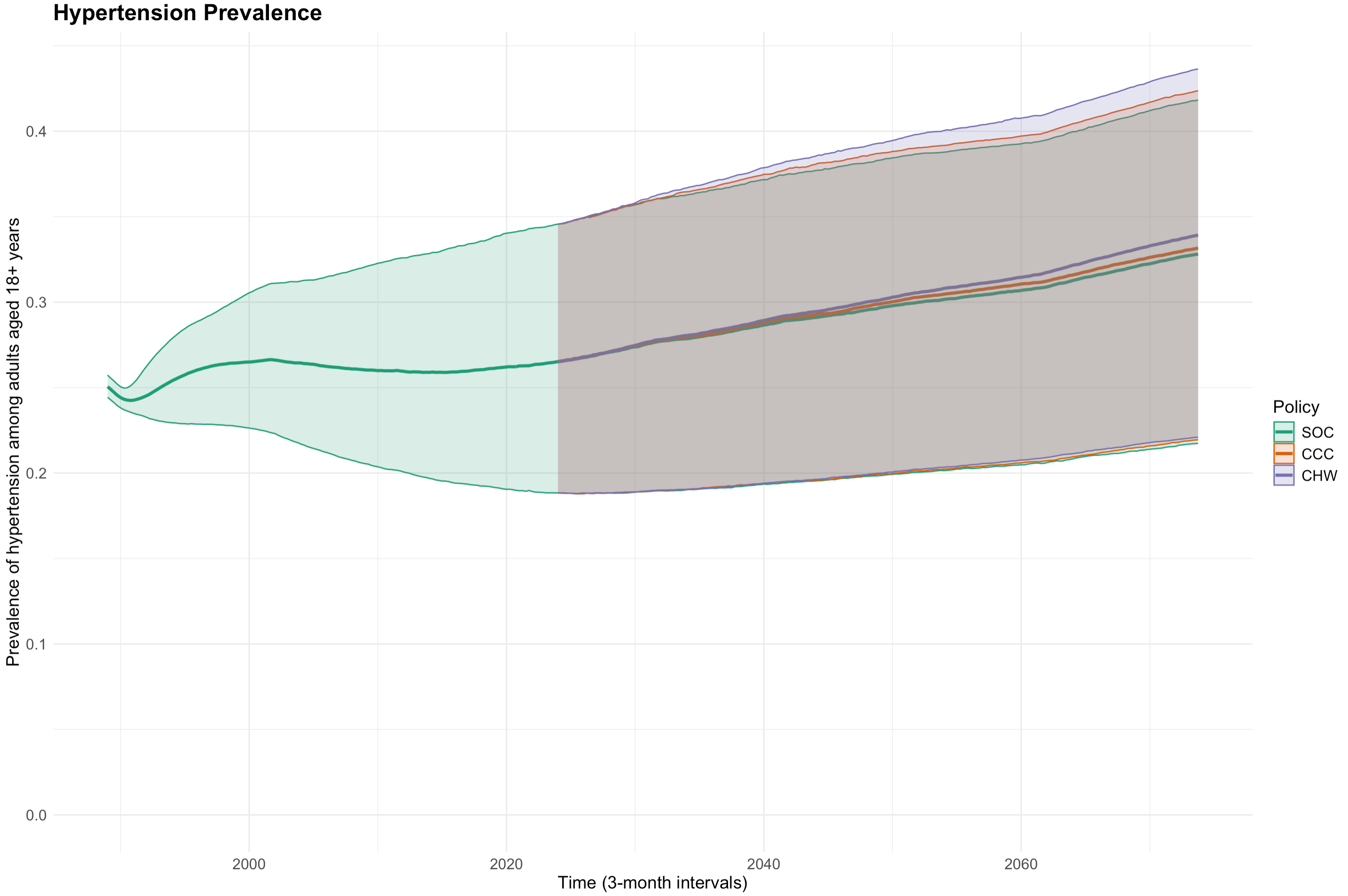

Median and 90% range of hypertension prevalence (true current or pre-treatment SBP ≥140, regardless of whether measured) across setting-scenarios (n=3000), plotted in 3-month intervals.

**Hypertension prevalence**

Average from 2024-2074

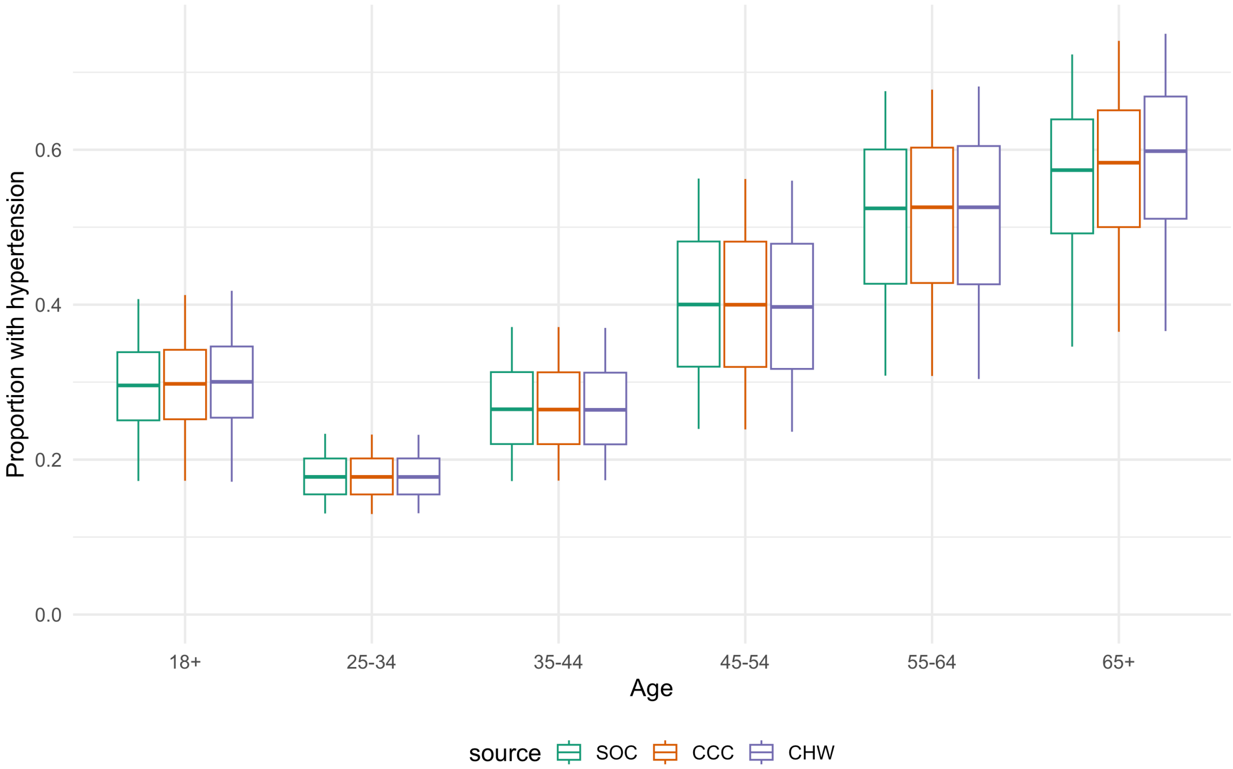

**Severe hypertension prevalence (SBP ≥160 mmHg)**

Average from 2024-2074, max SBP (regardless of current BP control)

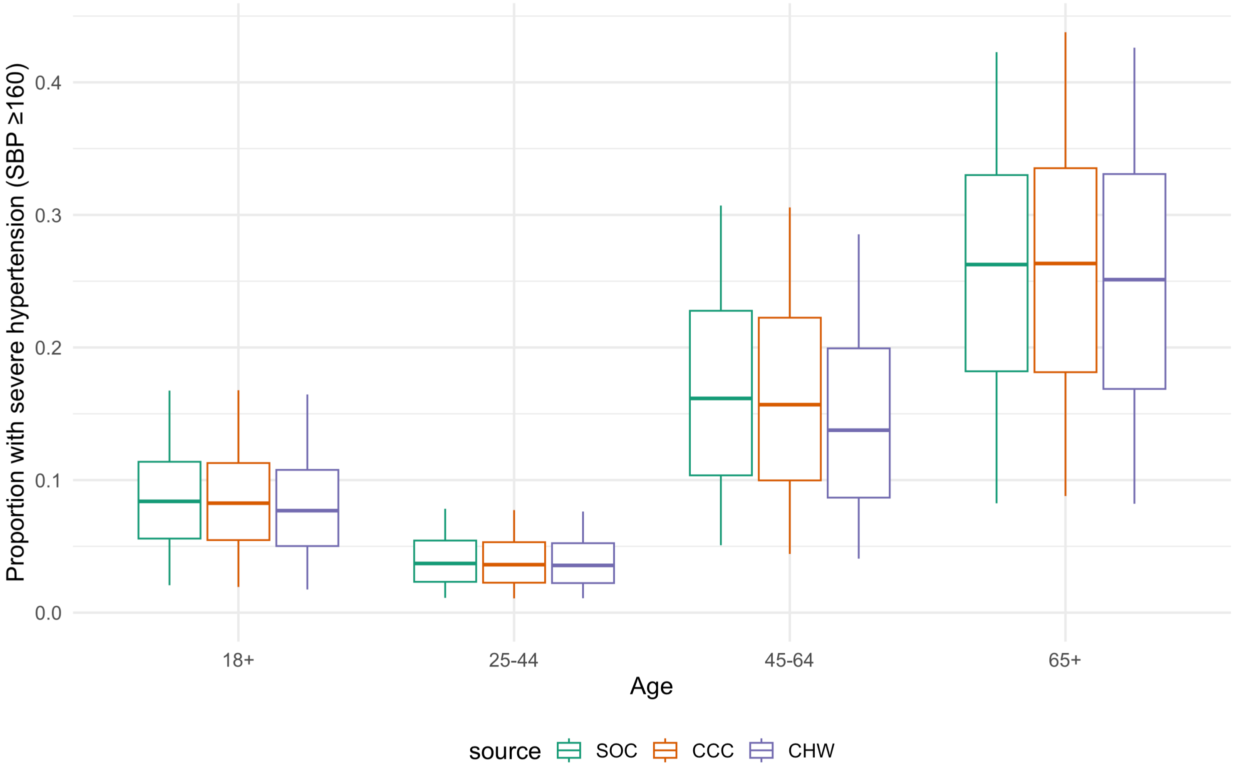

### Hypertension Diagnosis

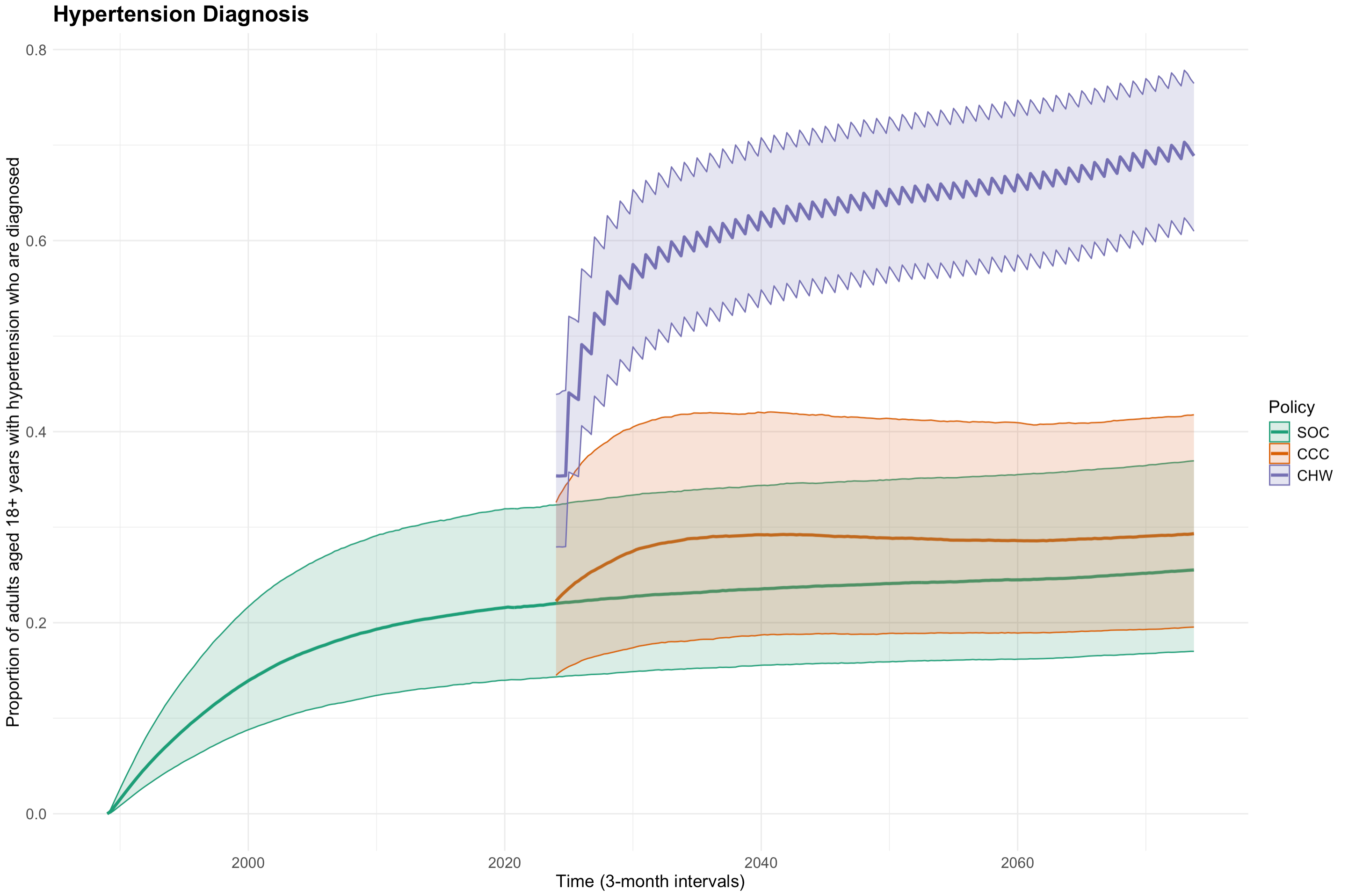

Median and 90% range of hypertension diagnosis across setting-scenarios (n=3000), plotted in 3-month intervals.

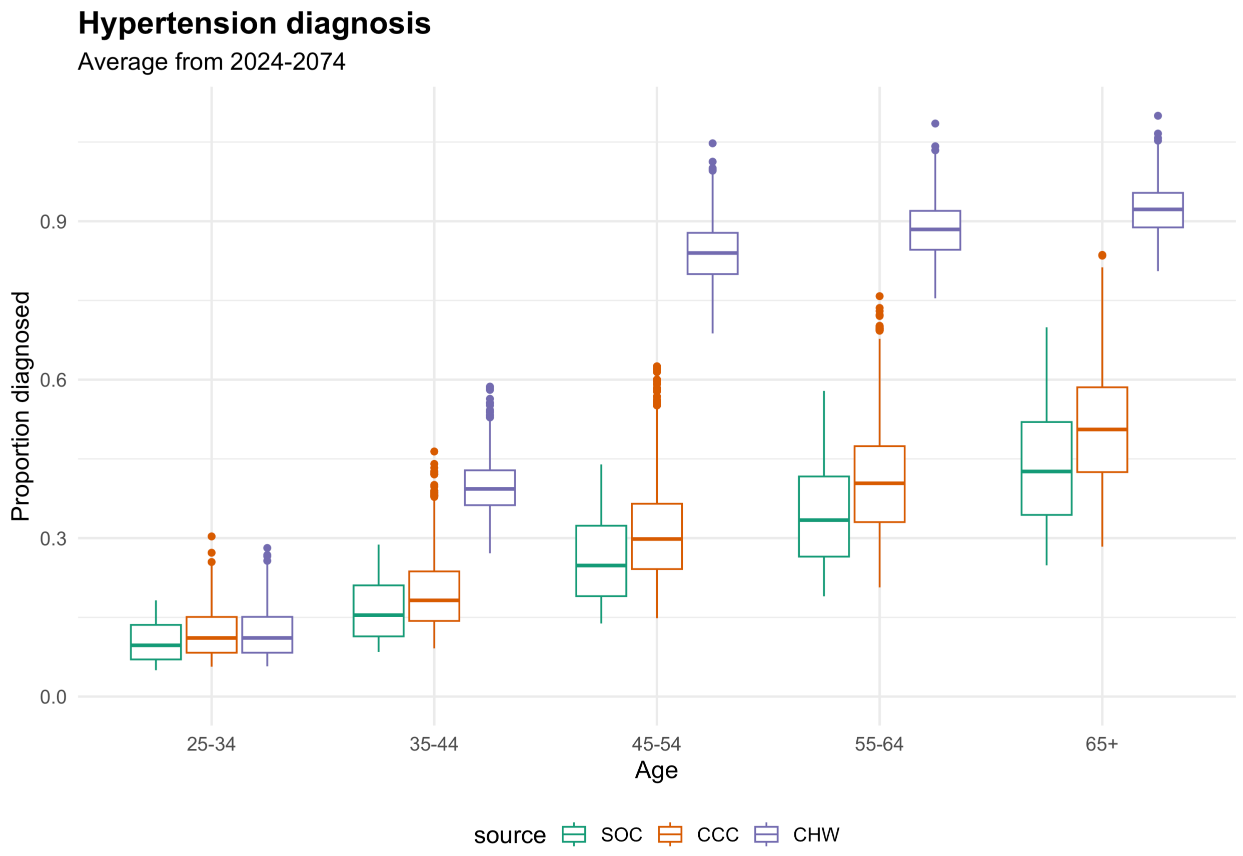

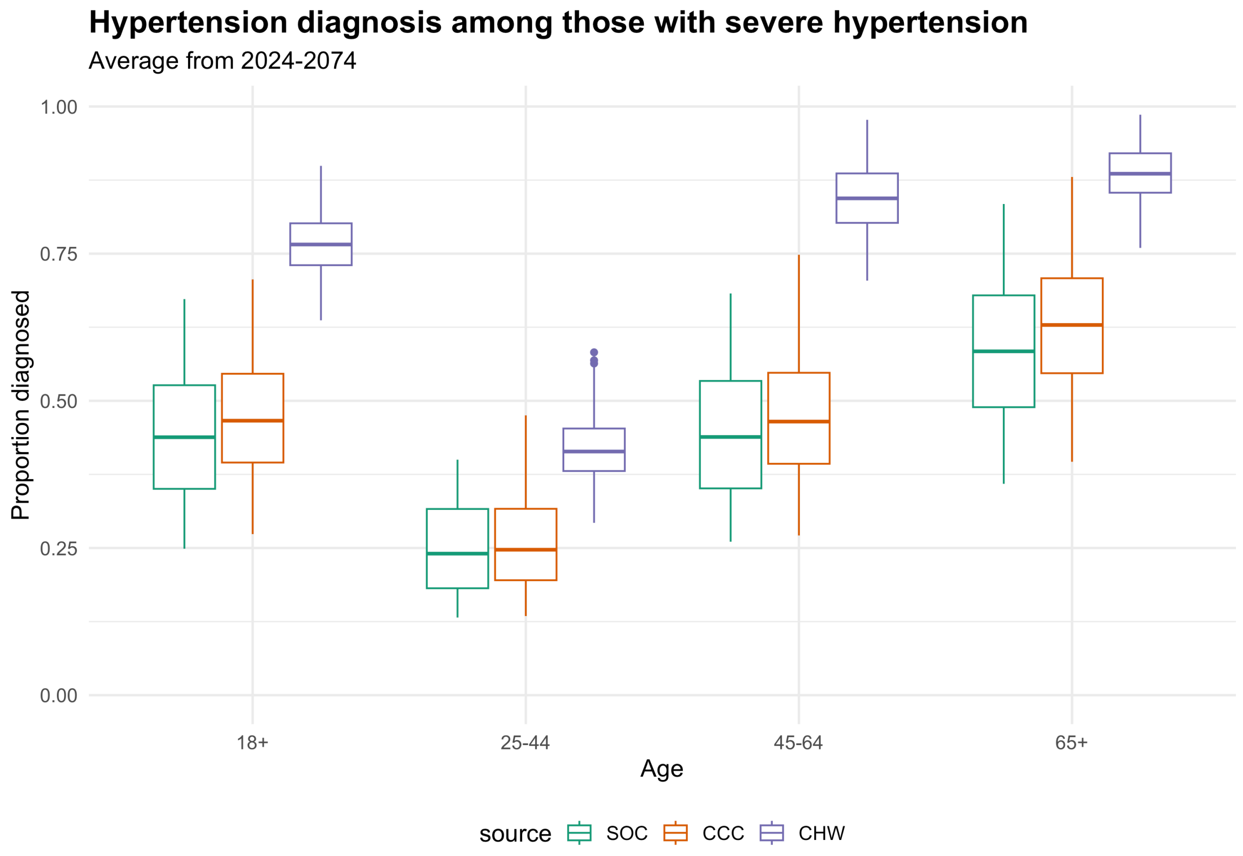

**Note:** Diagnosis can be >100% because the denominator is the proportion with true hypertension (SBP >=140) and the numerator is all with diagnosed hypertension including those who were overdiagnosed (measured BP >=140 but true BP <140). See additional graphs for overdiagnosis below.

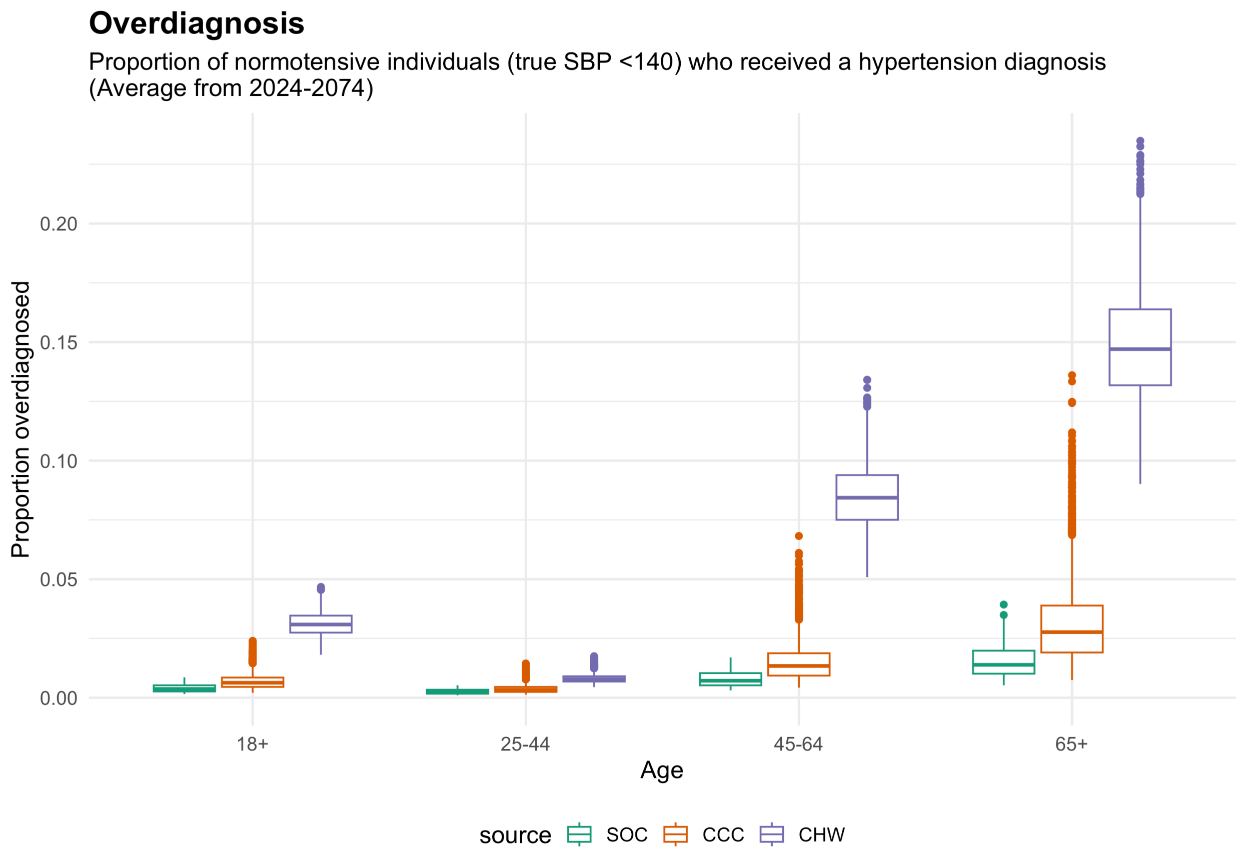
Overdiagnosis
Proportion of normotensive individuals (true SBP <140) who received a hypertension diagnosis
(Average from 2024-2074)
Proportion overdiagnosed
0.00
0.05
0.10
0.15
0.20
18+
25-44
45-64
65+
Age
source
SOC
CCC
CHW

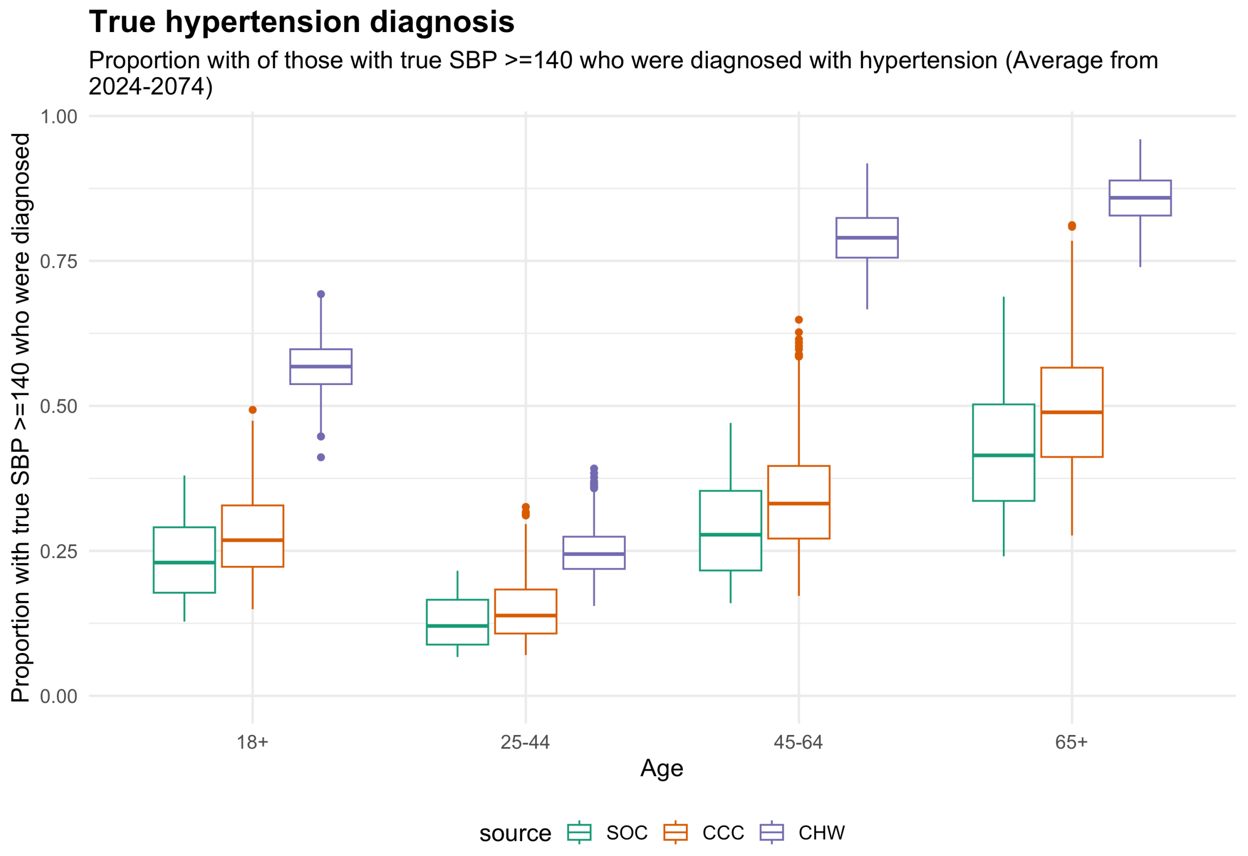
True hypertension diagnosis
Proportion with of those with true SBP >=140 who were diagnosed with hypertension (Average from
2024-2074)
Proportion with true SBP >=140 who were diagnosed
0.00
0.25
0.50
0.75
1.00
18+
25-44
45-64
65+
Age
source
SOC
CCC
CHW

### Hypertension Treatment

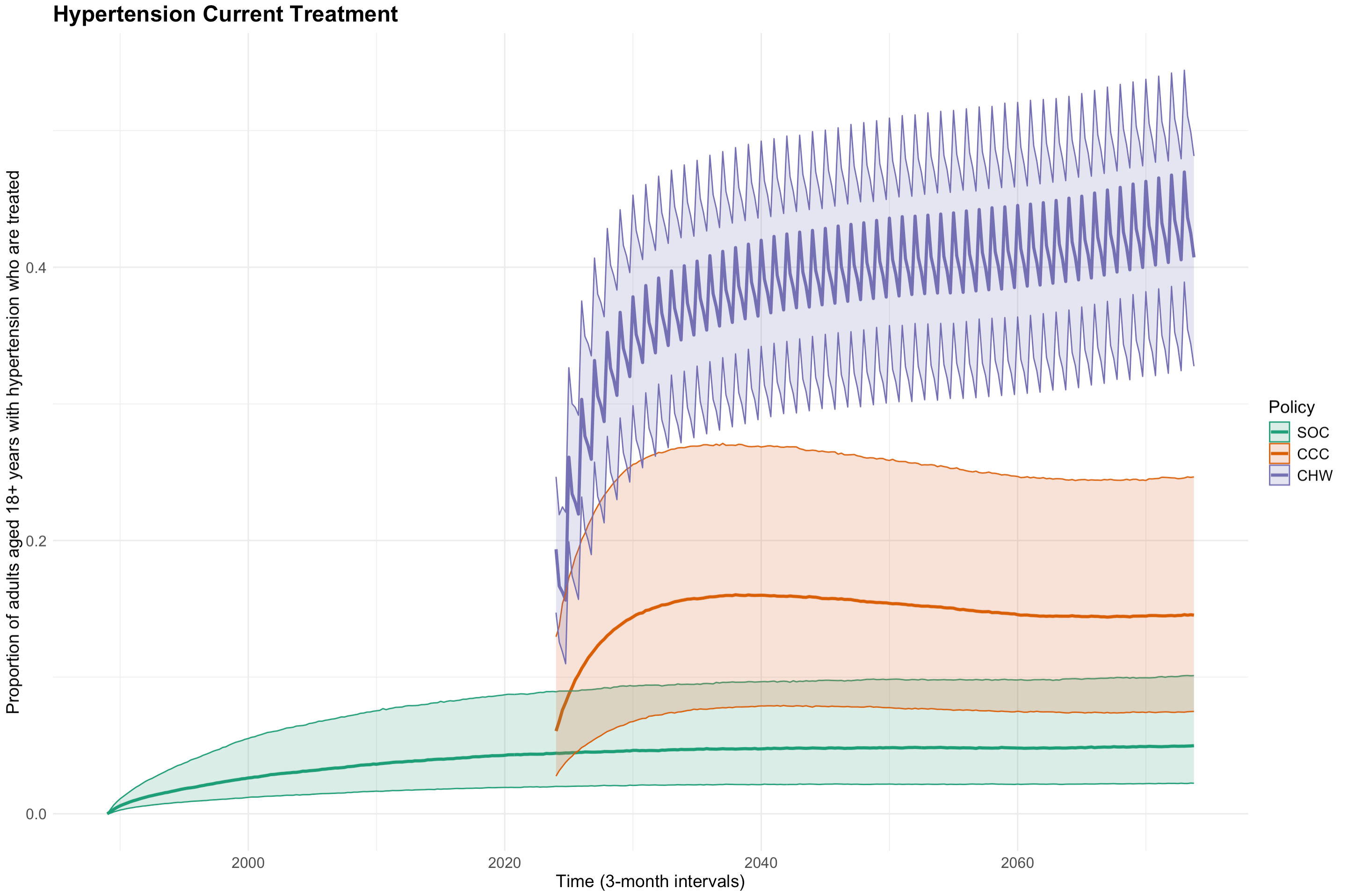

Median and 90% range of current hypertension treatment across setting-scenarios (n=3000), plotted in 3-month intervals.

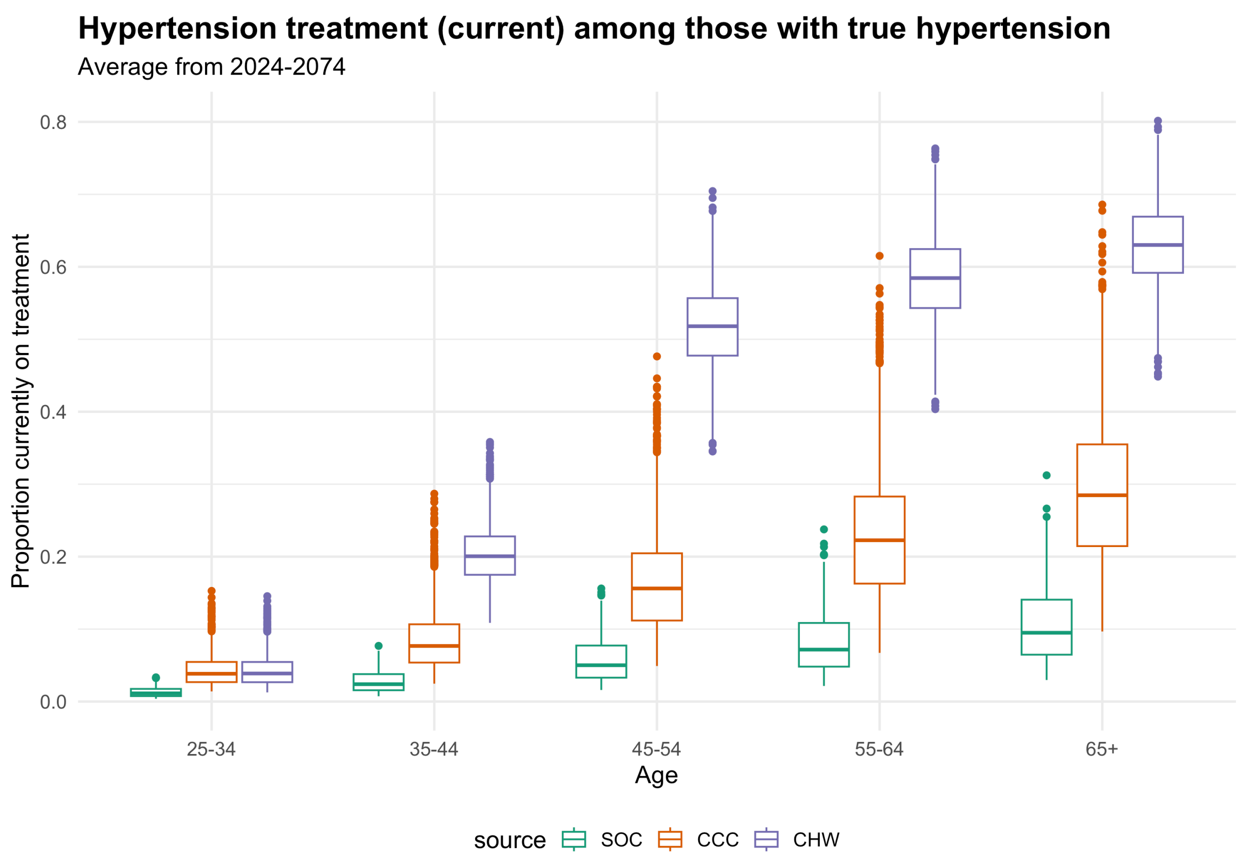

Hypertension treatment (current) among those with true hypertension
Average from 2024-2074
Proportion currently on treatment
0.0
0.2
0.4
0.6
0.8
25-34
35-44
45-54
55-64
65+
Age
source
SOC
CCC
CHW

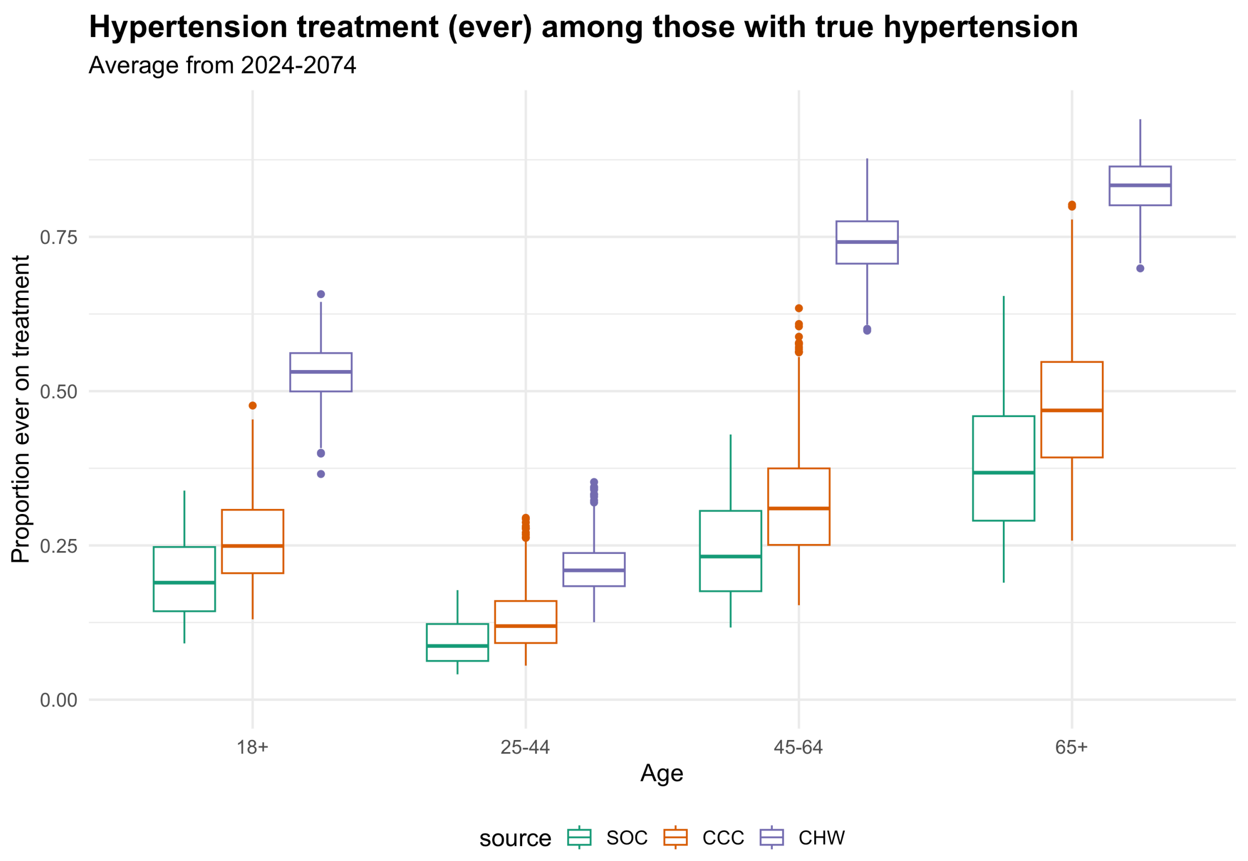

Hypertension treatment (ever) among those with true hypertension
Average from 2024-2074
Proportion ever on treatment
0.00
0.25
0.50
0.75
18+
25-44
45-64
65+
Age
source
SOC
CCC
CHW

#### Overtreatment

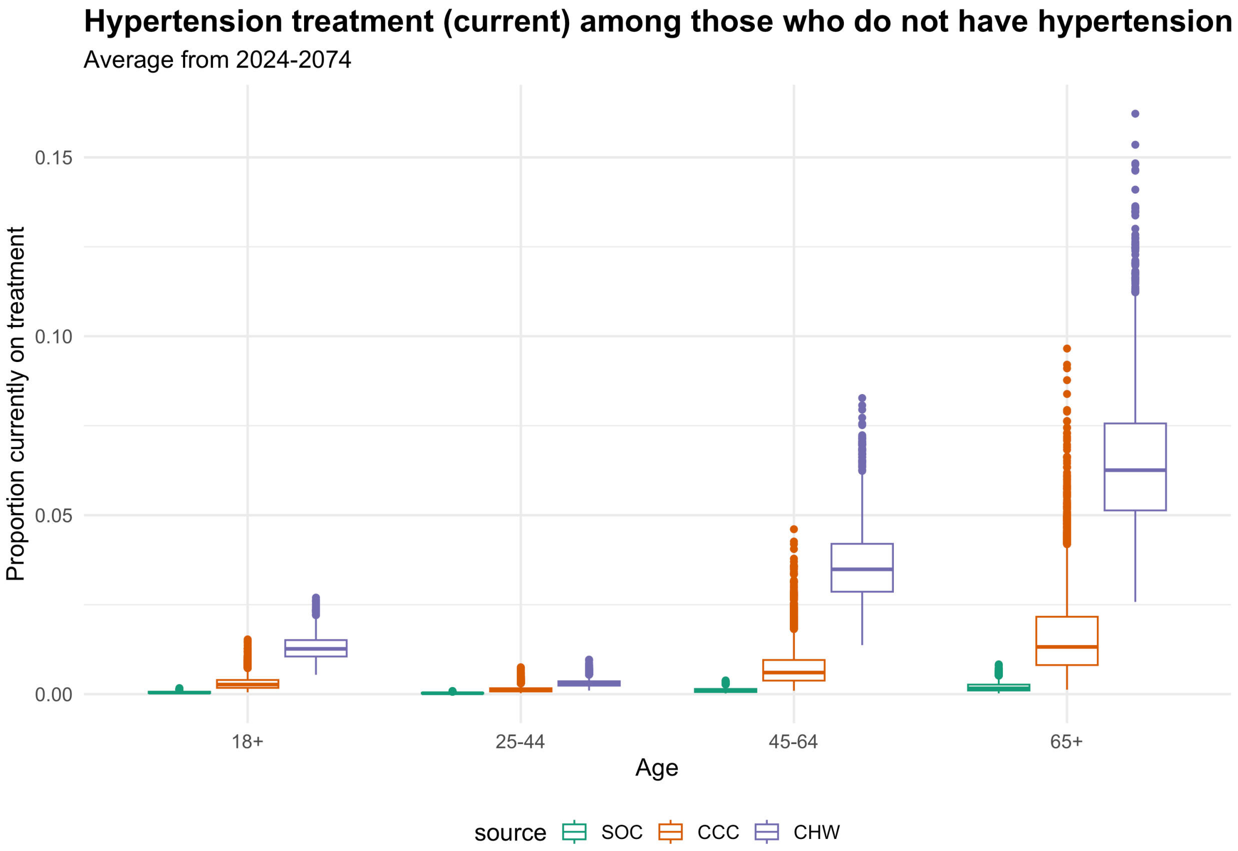

### Hypertension control

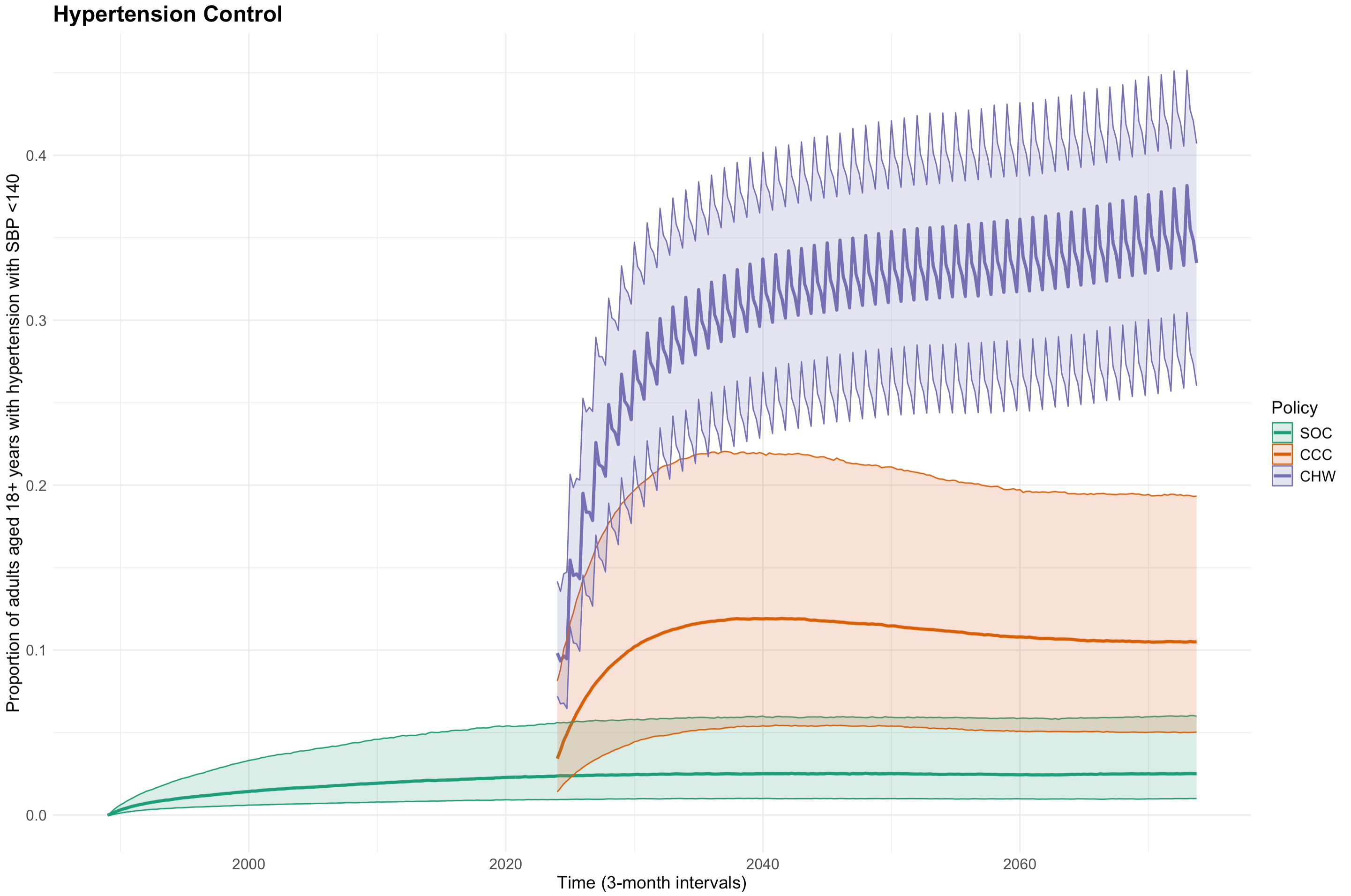

Median and 90% range of hypertension control (SBP <140 mmHg) among individuals with hypertension across setting-scenarios (n=3000), plotted in 3-month intervals.

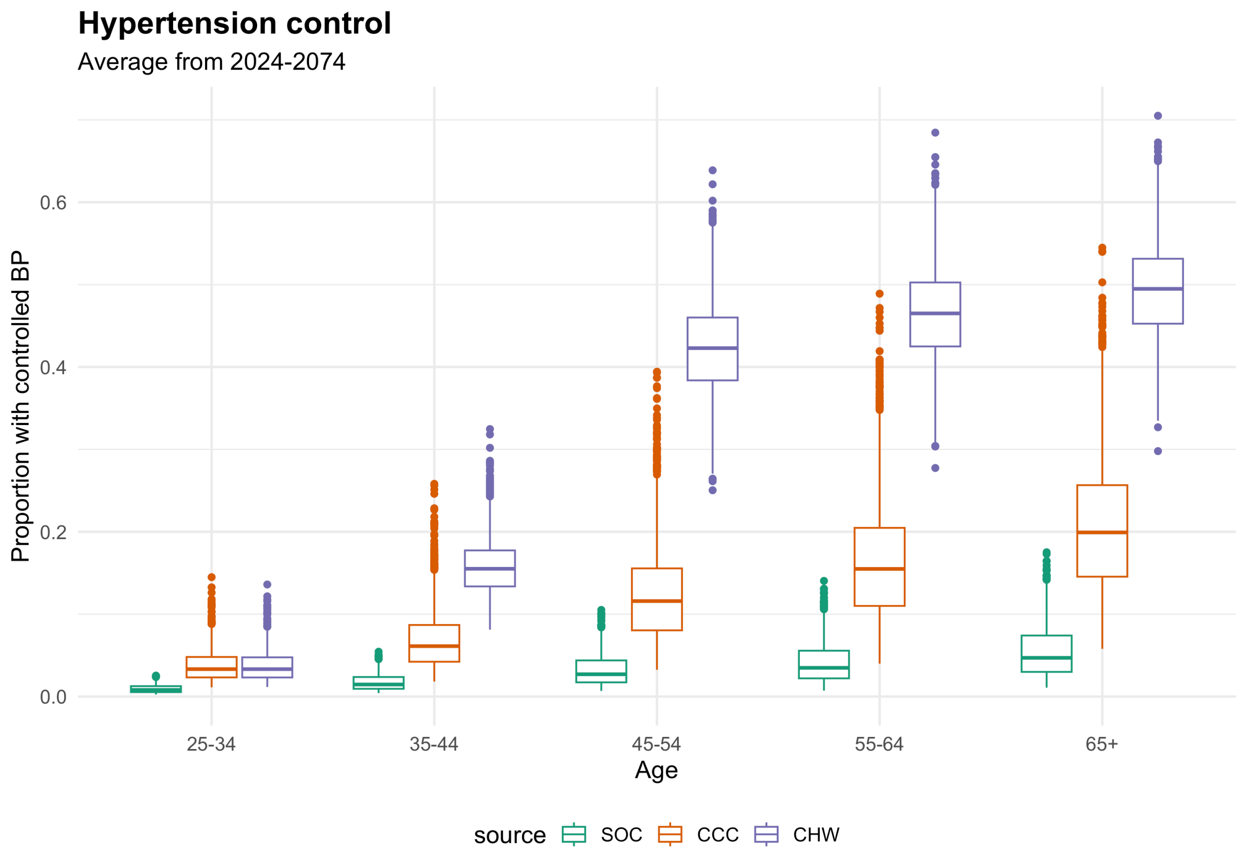
Hypertension control
Average from 2024-2074
Proportion with controlled BP
0.0
0.2
0.4
0.6
25-34
35-44
45-54
55-64
65+
Age
source
SOC
CCC
CHW

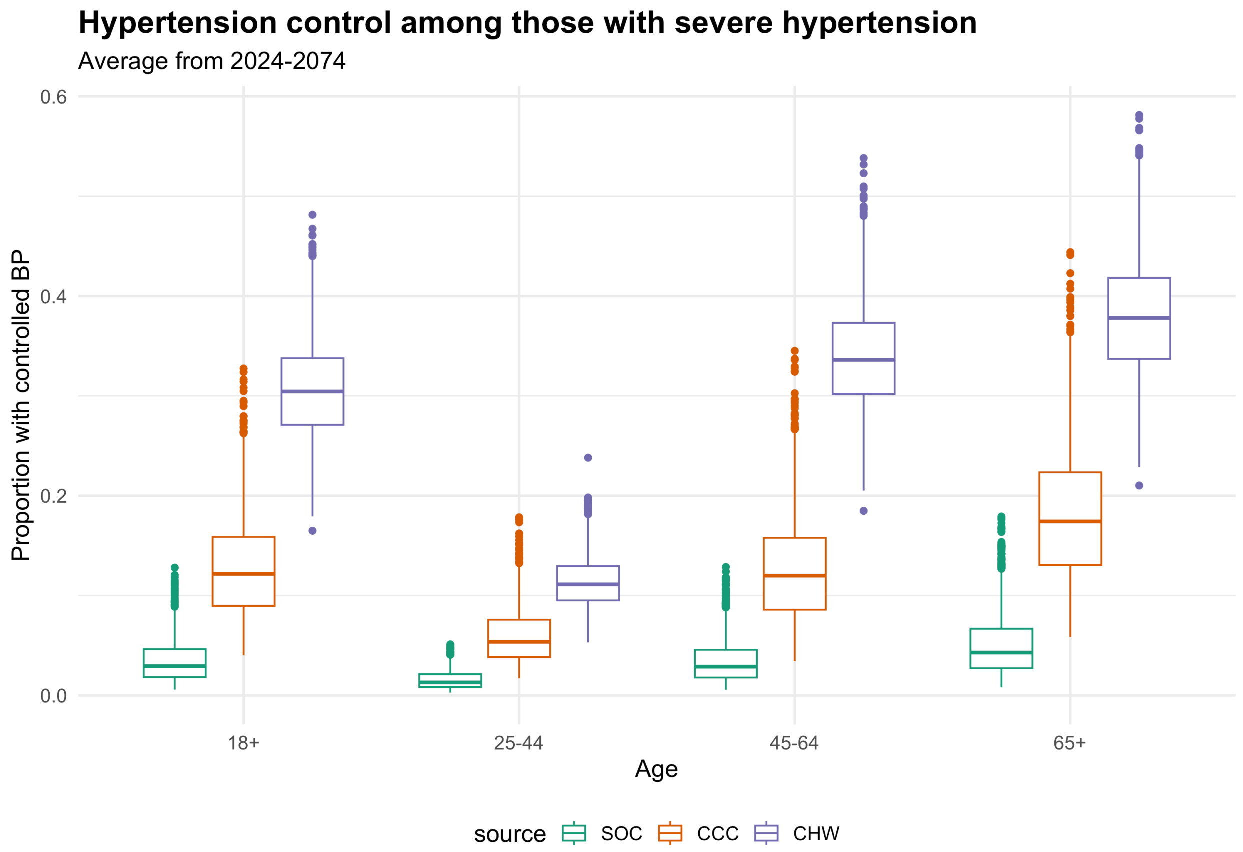
Hypertension control among those with severe hypertension
Average from 2024-2074
Proportion with controlled BP
0.0
0.2
0.4
0.6
18+
25-44
45-64
65+
Age
source
SOC
CCC
CHW

#### SBP Increase with Age

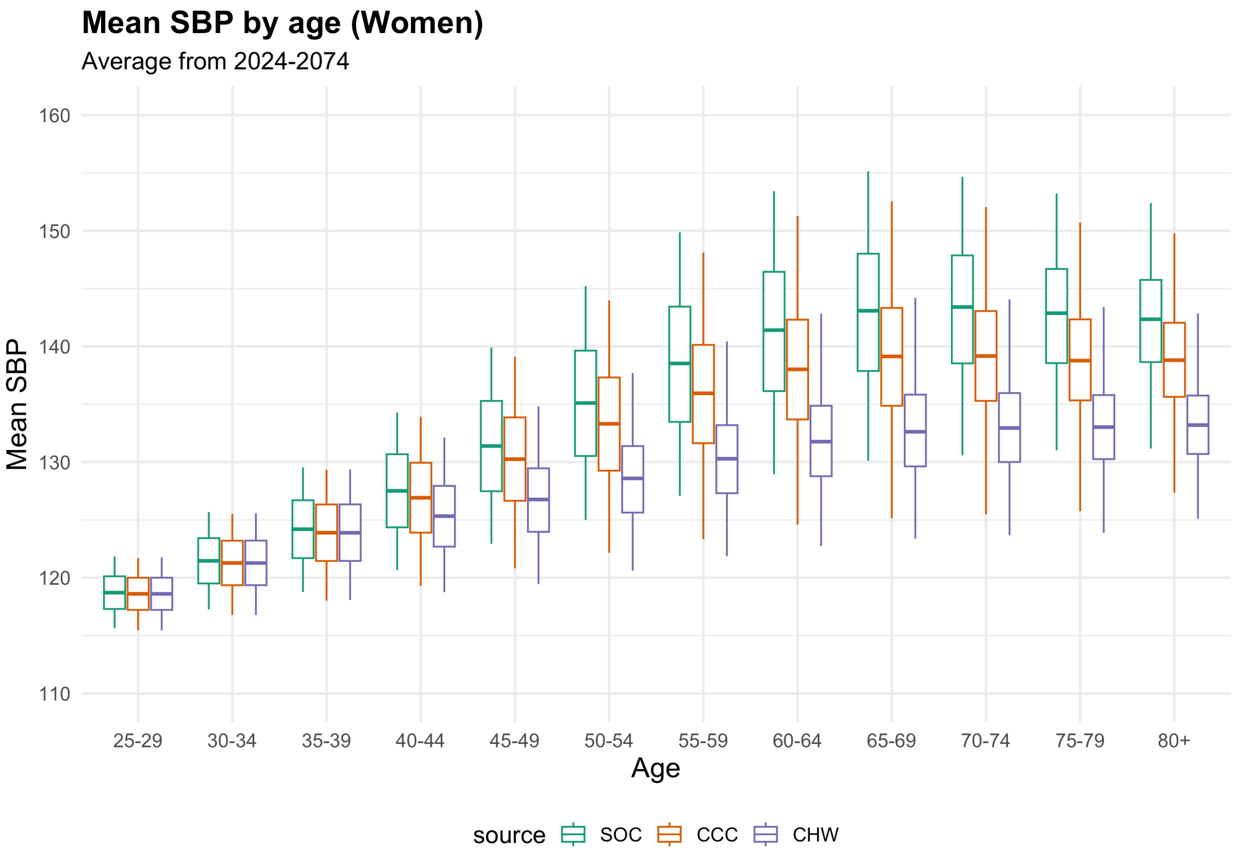

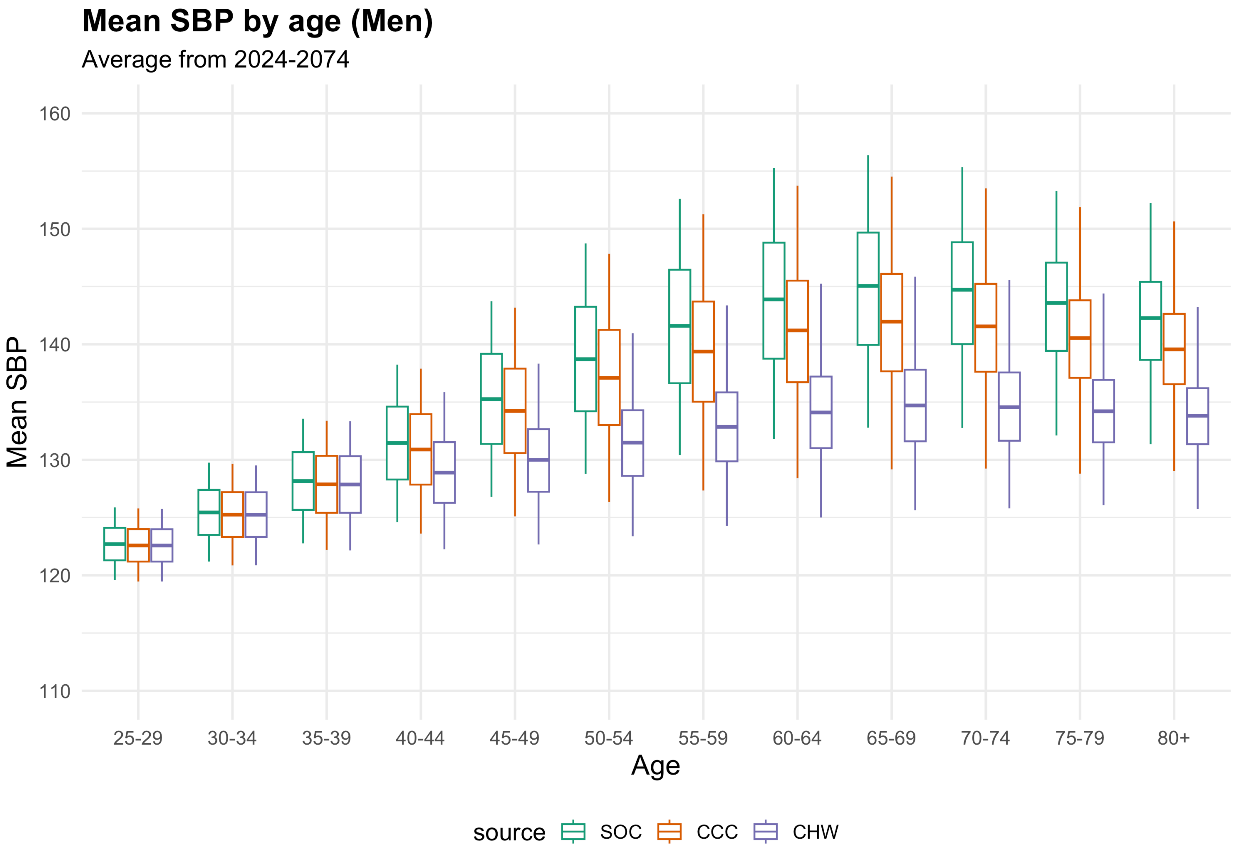

#### CVD Event incidence

*Incident Ischemic Heart Disease Events*

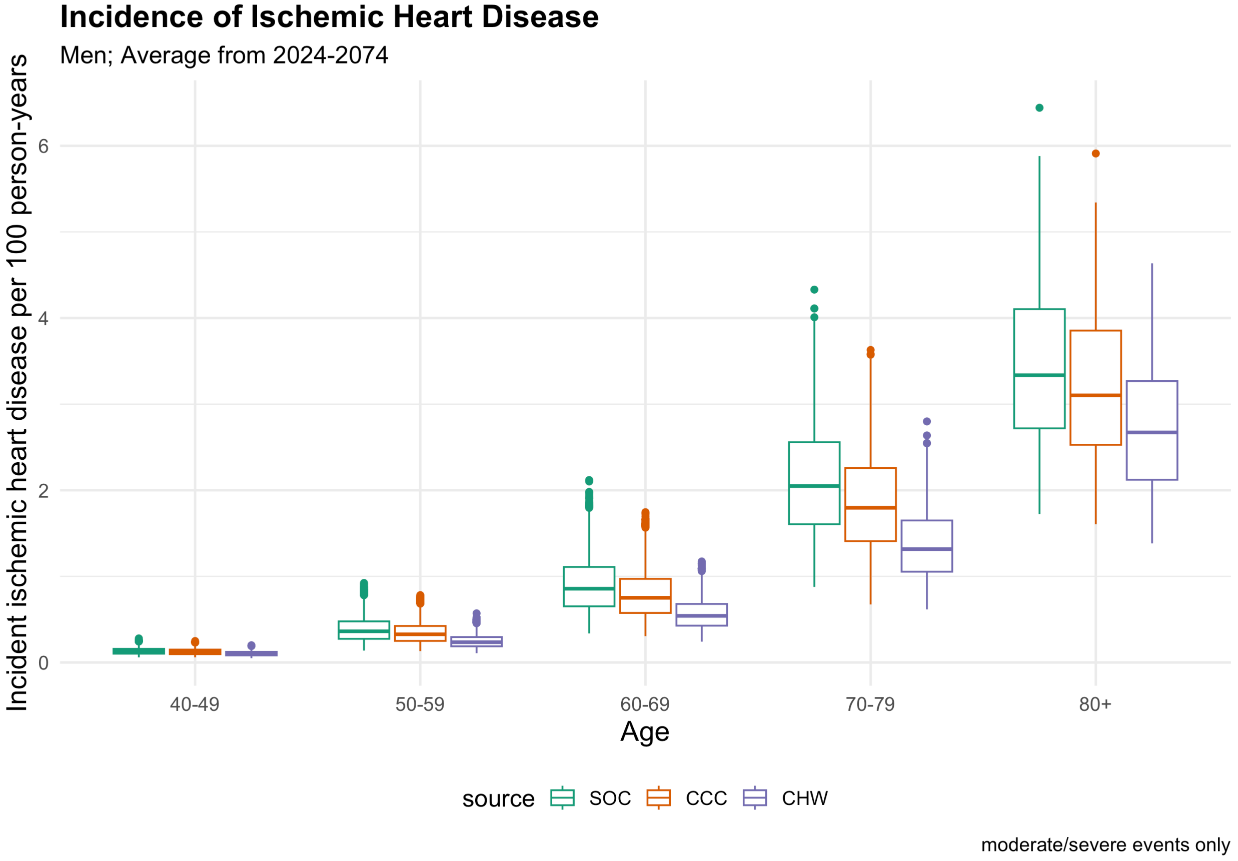

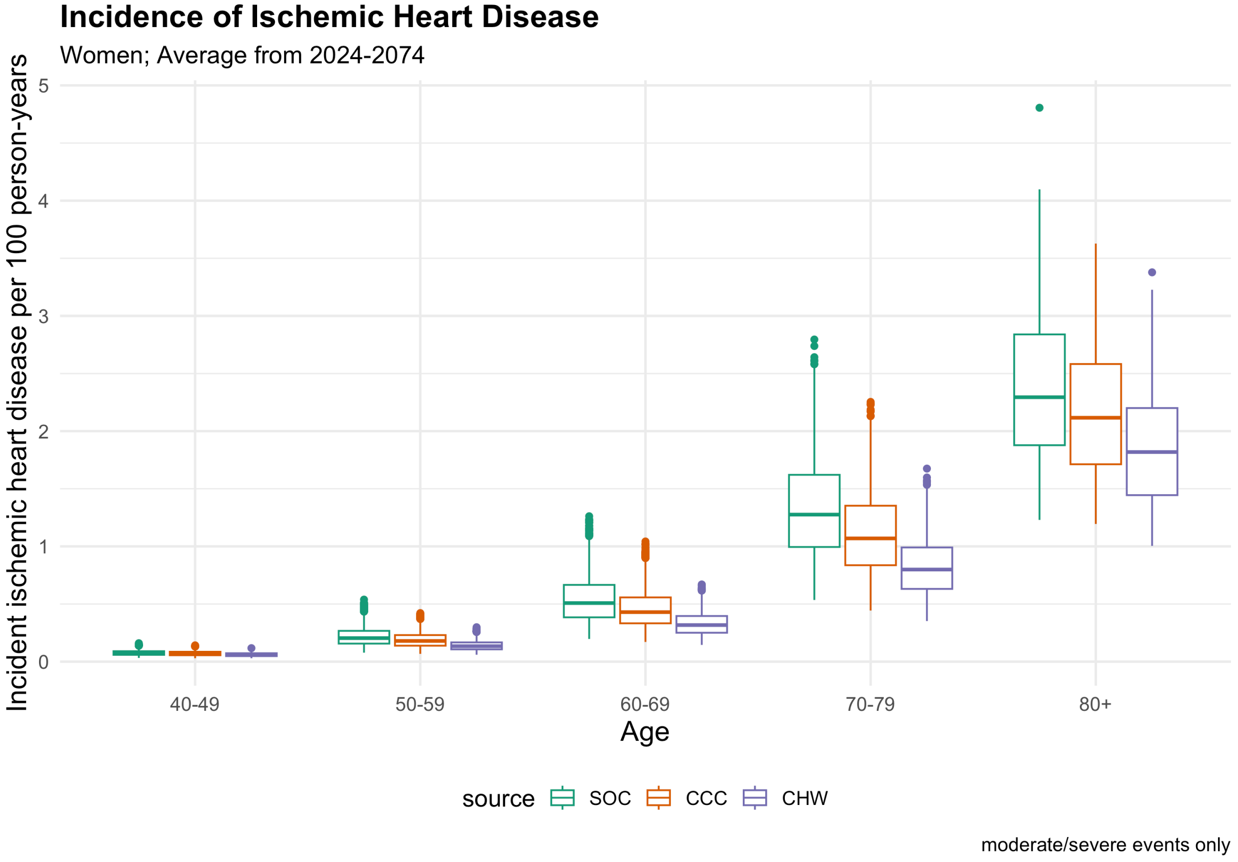

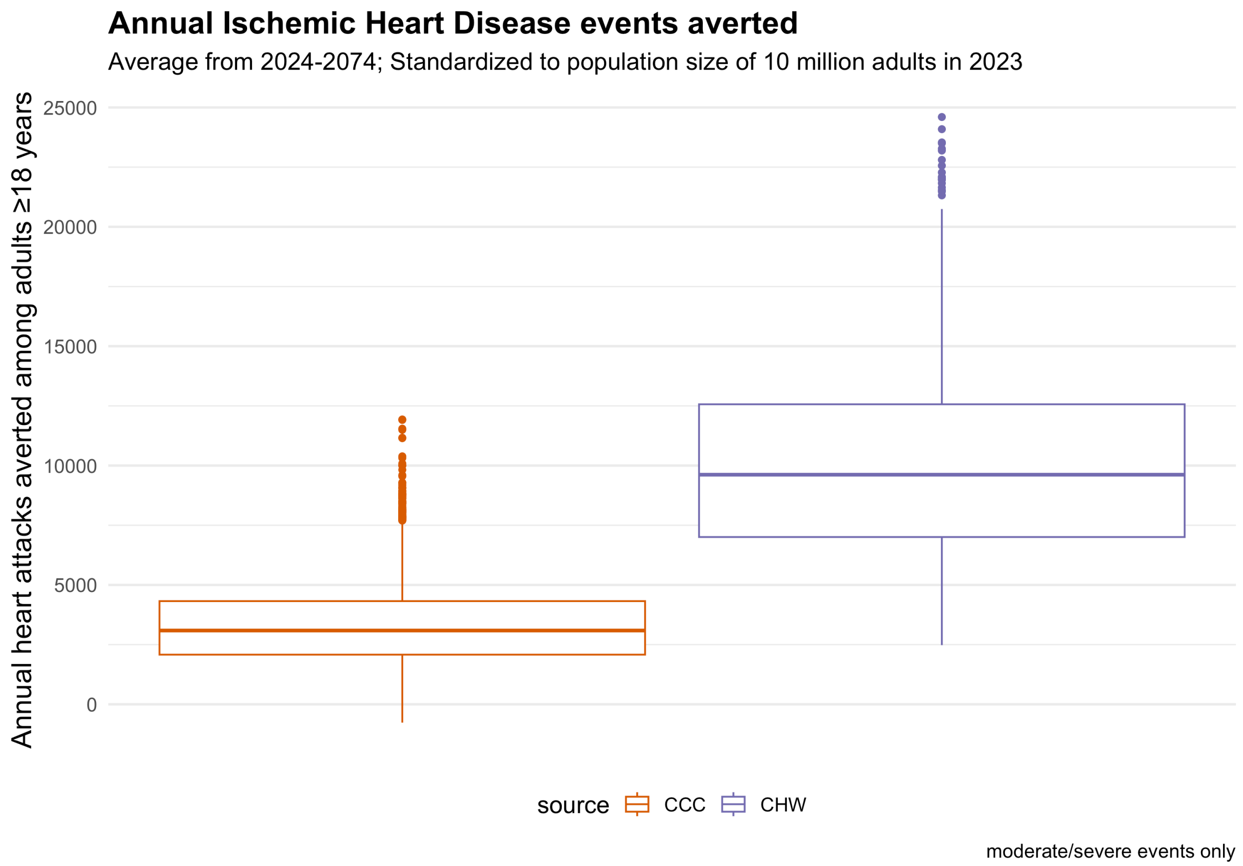

*Incident Stroke*

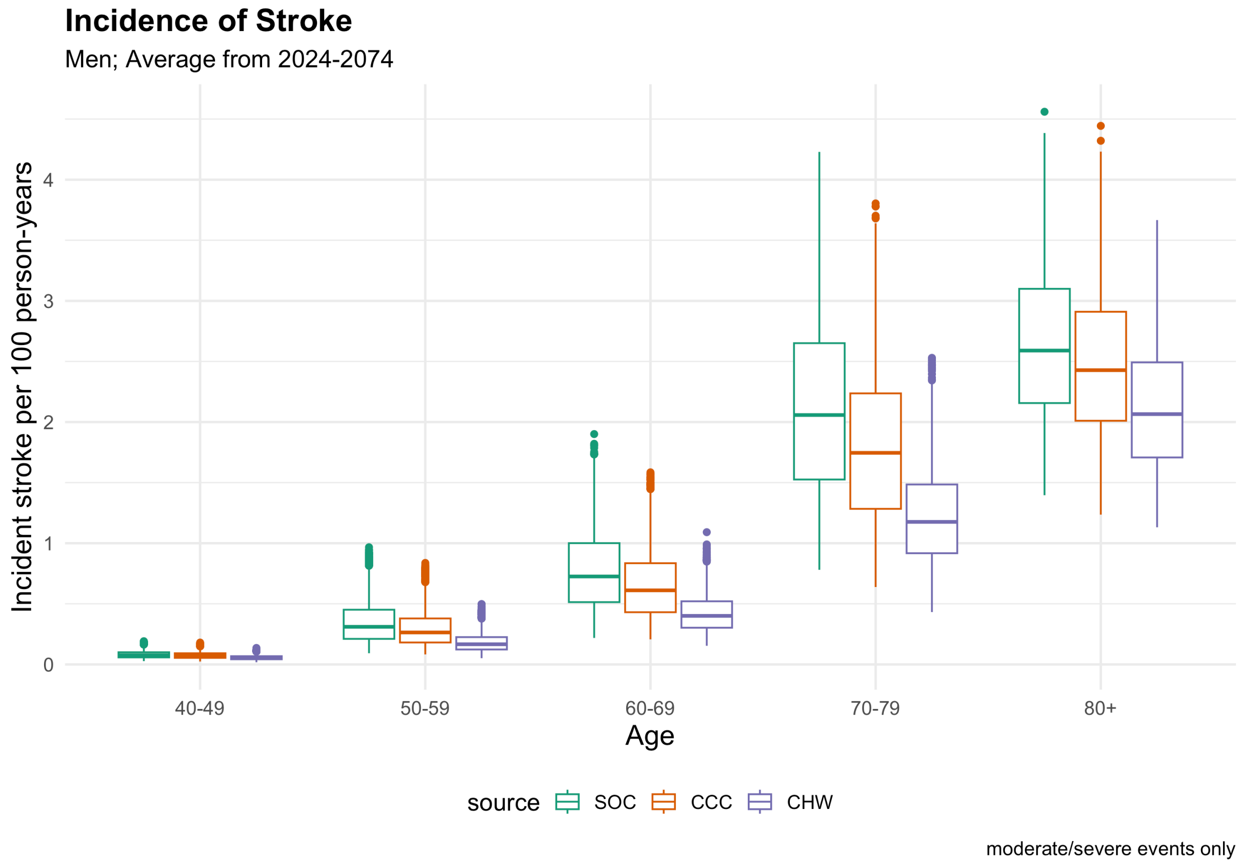

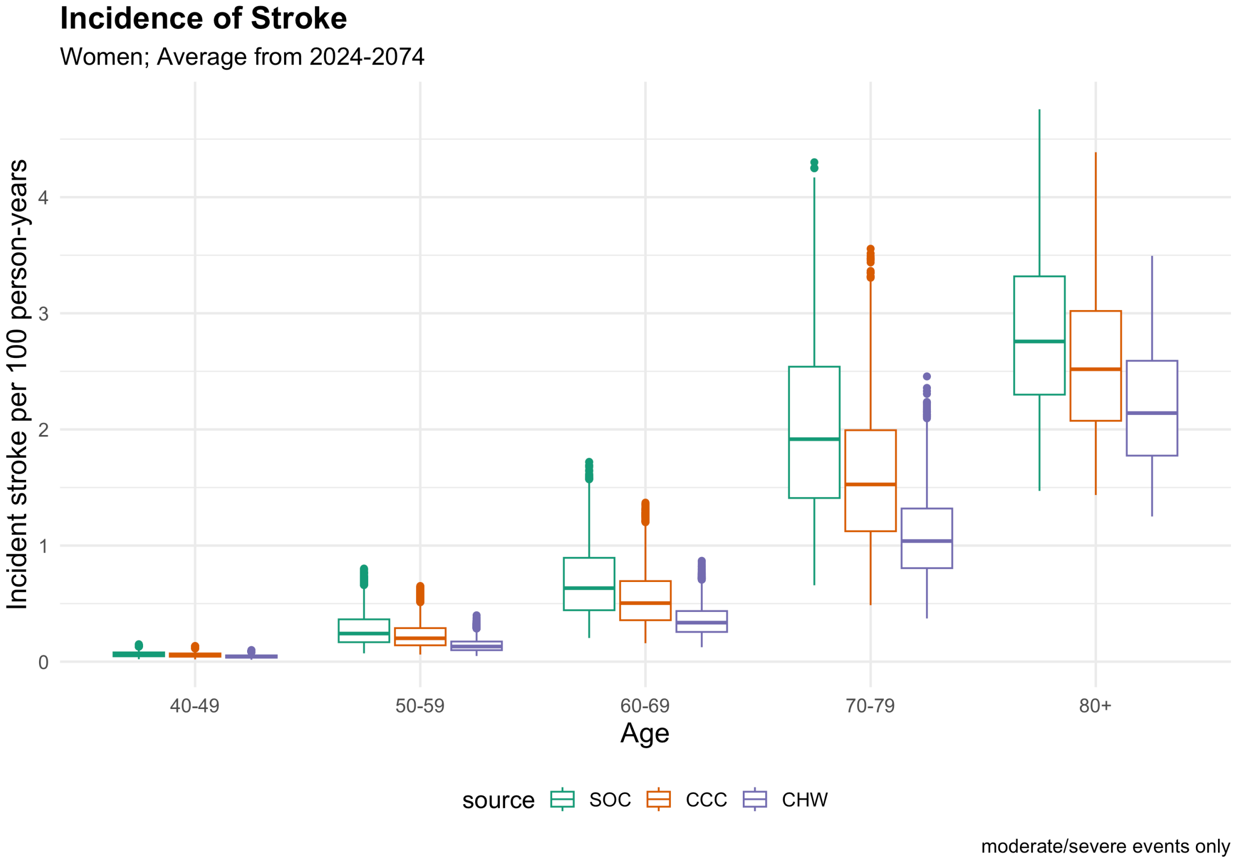
Incidence of Stroke
Women; Average from 2024-2074
Incident stroke per 100 person-years
0
1
2
3
4
40-49
50-59
60-69
70-79
80+
Age
source
SOC
CCC
CHW
moderate/severe events only

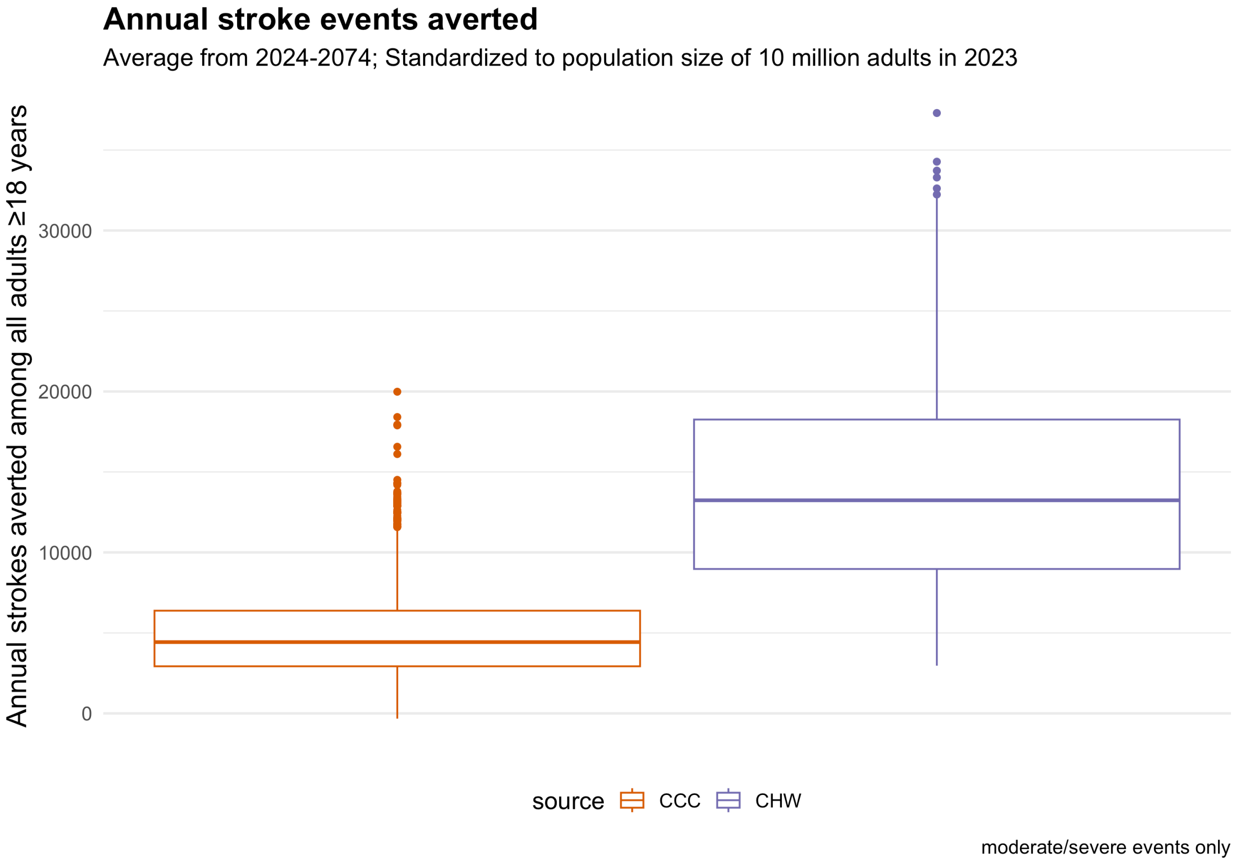
Annual stroke events averted
Average from 2024-2074; Standardized to population size of 10 million adults in 2023
Annual strokes averted among all adults ≥18 years
0
10000
20000
30000
source
CCC
CHW
moderate/severe events only

#### CVD Mortality

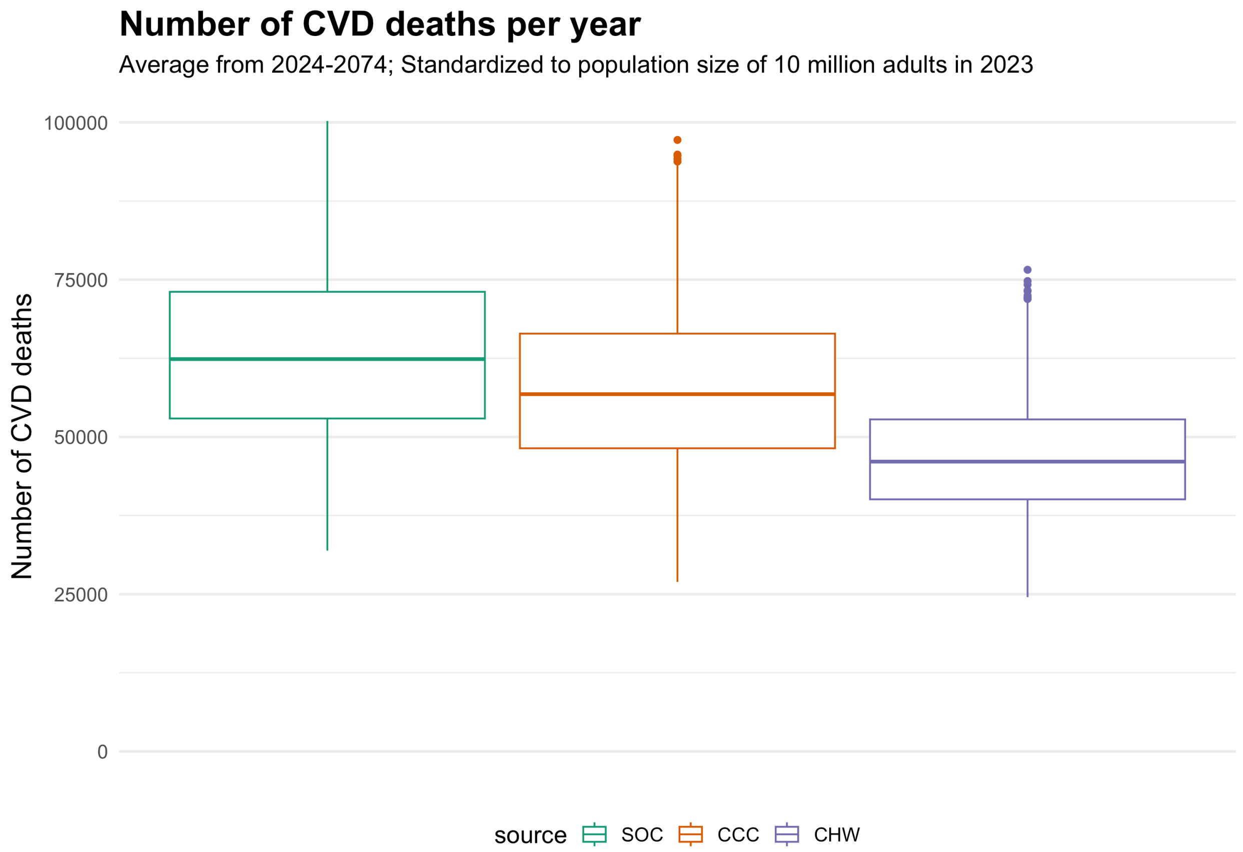

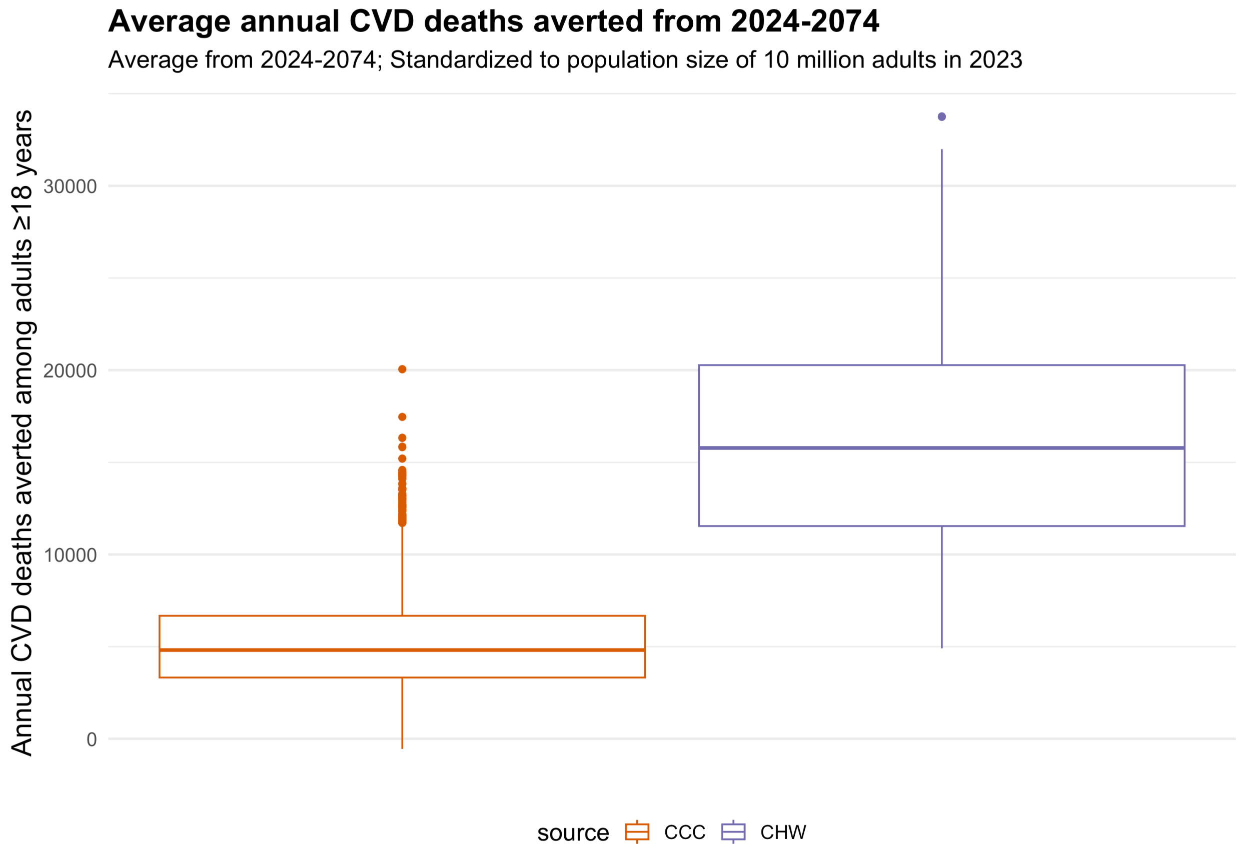

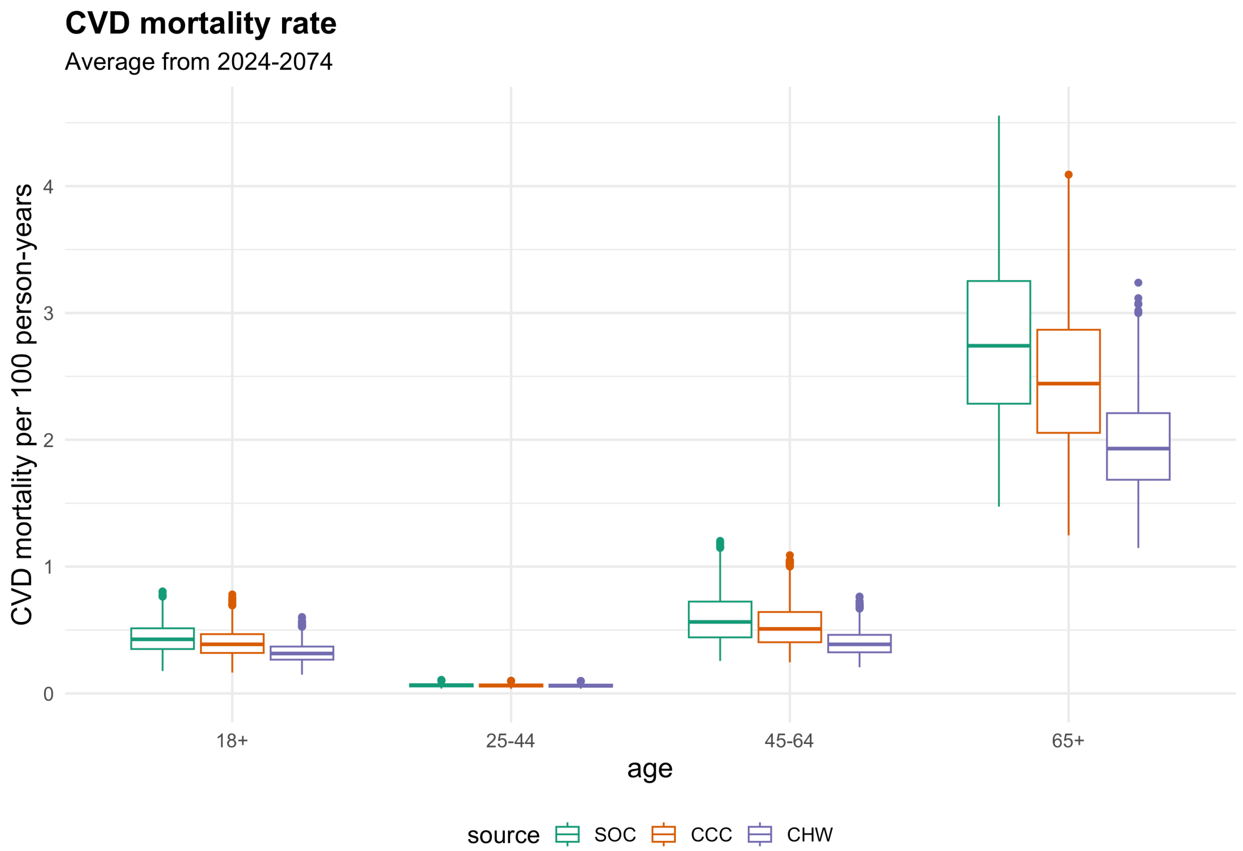

CVD mortality rate
Average from 2024-2074
CVD mortality per 100 person-years
0
1
2
3
4
18+
25-44
45-64
65+
age
source
SOC
CCC
CHW

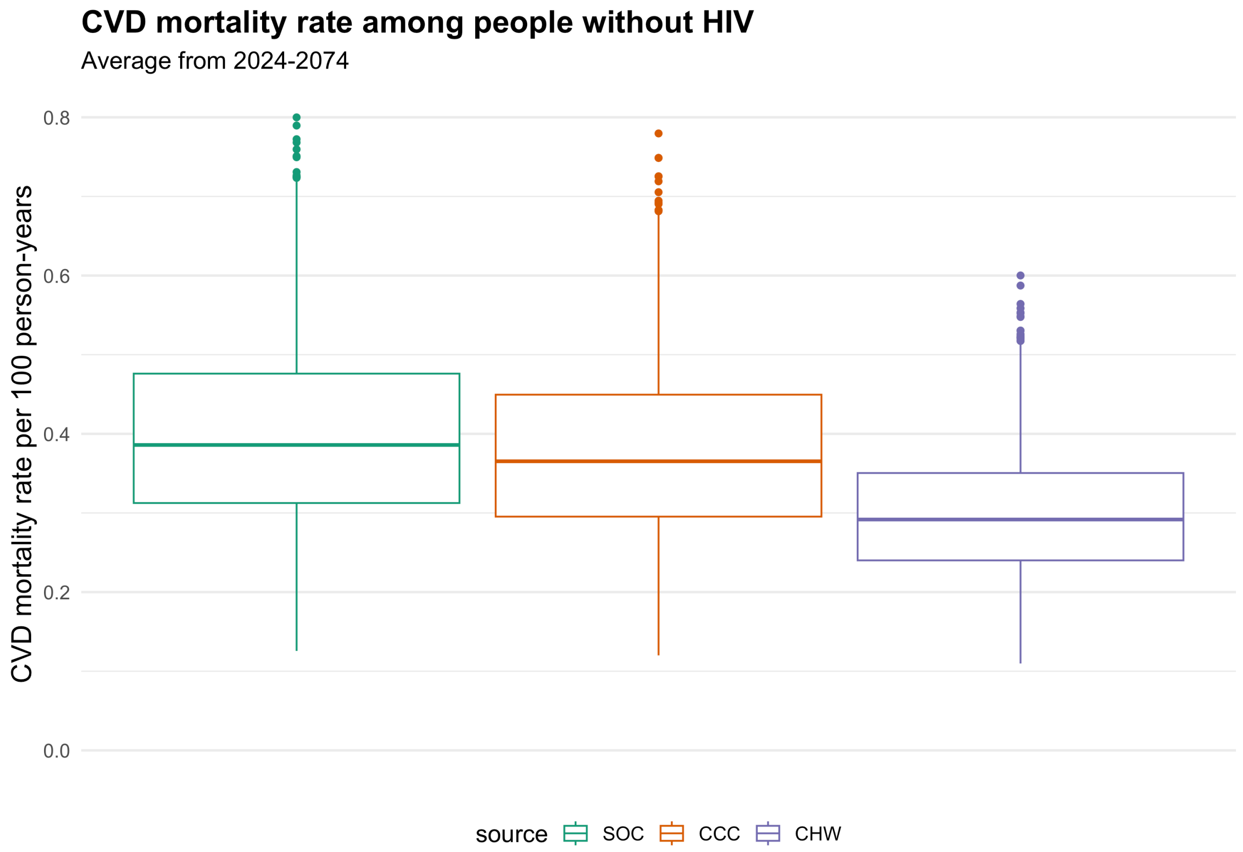

CVD mortality rate among people without HIV
Average from 2024-2074
CVD mortality rate per 100 person-years
0.0
0.2
0.4
0.6
0.8
source
SOC
CCC
CHW

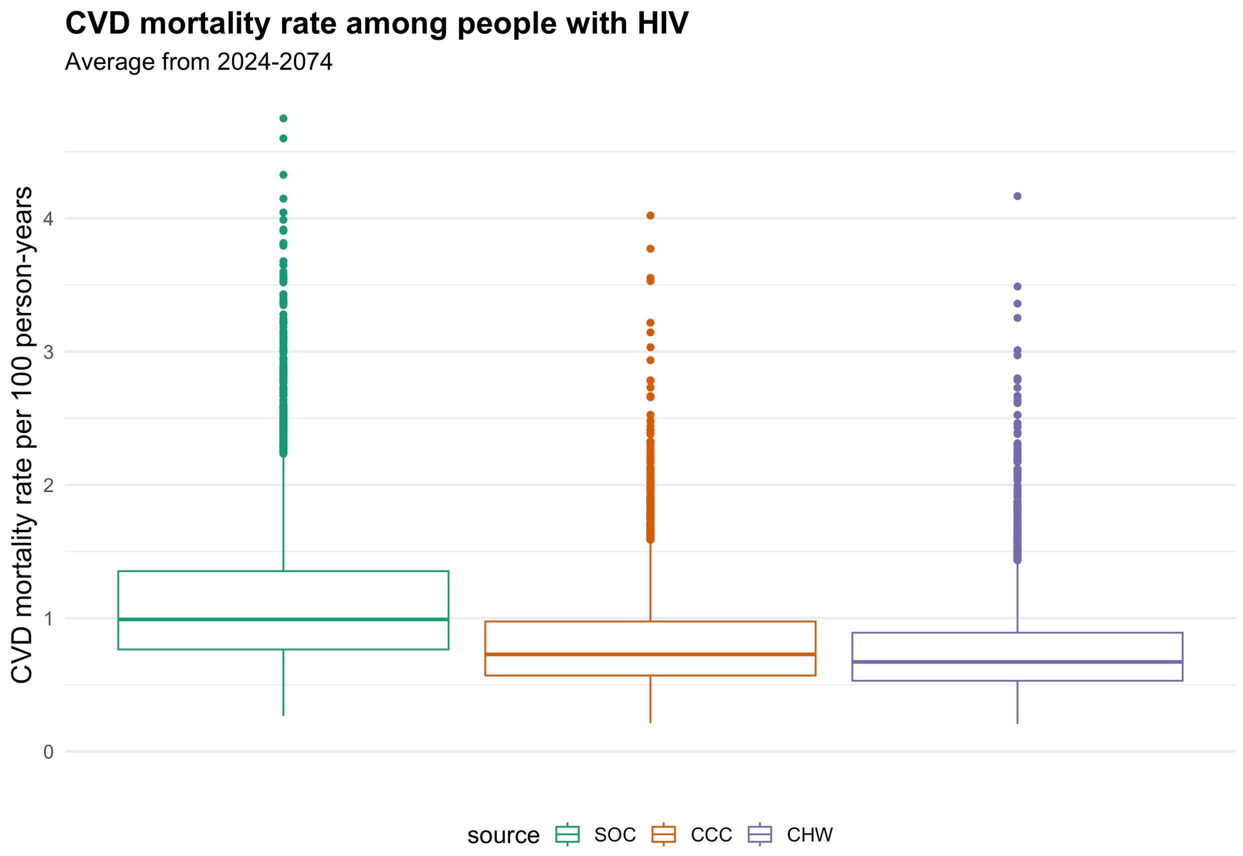
CVD mortality rate among people with HIV
Average from 2024-2074
CVD mortality rate per 100 person-years
0
1
2
3
4
source
SOC
CCC
CHW

#### All-Cause Mortality

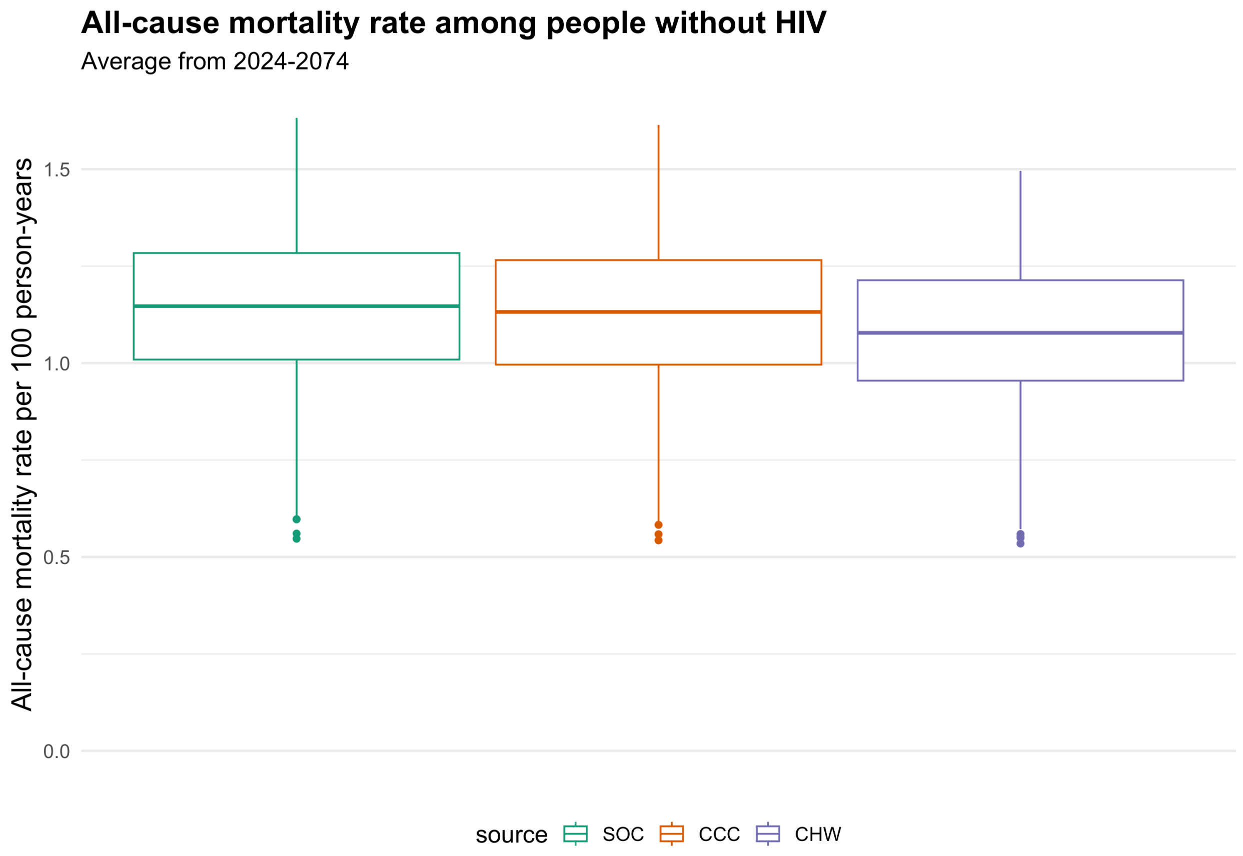

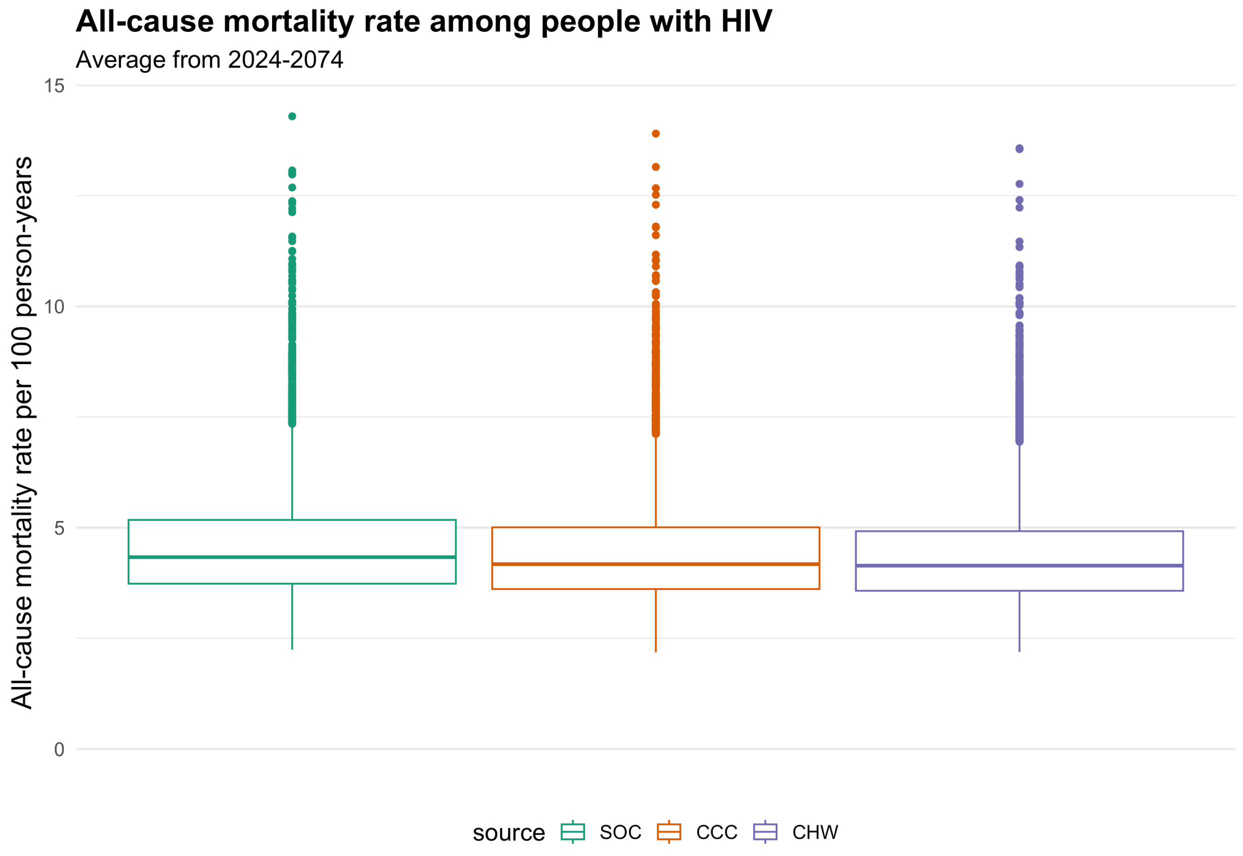

#### Cost

Cost assumptions (in USD):

- Community health worker screening: $3/person screened (estimates from SEARCH)
- Transportation voucher: $5/person linked (Hickey 2022)
- Clinic visits: $20/visit if uncontrolled and $10/visit if controlled (Shade 2021, Shiri 2021)
- Fixed clinic training costs for CCC and CHW policies: $0.96 million USD per 10 million adult population
- HTN medications (90-day supply) (Resolve to Save Lives 2022)
  - Med 1 & 2: $1.5
  - Med 3: $3
- Acute MI/CVA treatment (Subramanian 2019)
  - MI: $2270
  - CVA: $2420
  - cost of acute care that is not effective (poor-quality):
    - MI: $567.5 / $1135 / $2270
    - CVA: $605 / $1210 / $2420
    - we assume that some poor-quality care may be due in part to limited staffing, diagnostic equipment, and medications - and therefore may also cost less. In other cases, similar interventions/diagnostic tests may be undertaken, though the effectiveness will be lower due to delays in care. We assume that such care for acute MI/CVA may cost lower than the cost of effective acute care (sampled from 25%, 50%, and 100% of the cost of effective care)

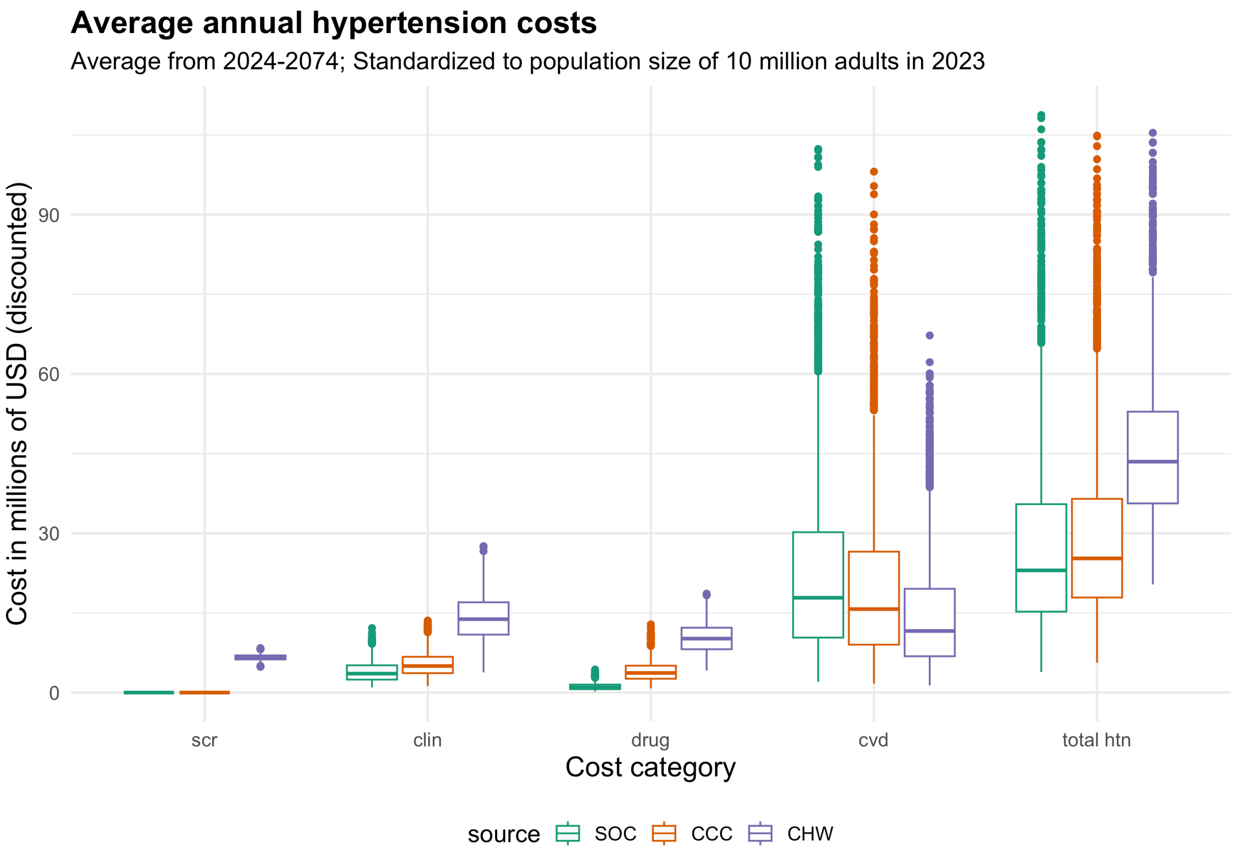

Note that CVD costs vary widely because we sample from a range of CVD care utilization and cost in the main model, in addition to sensitivity analyses with different cost scenarios (Table 5 in the main text).

#### Cost-effectiveness

##### Cost-effectiveness frontier

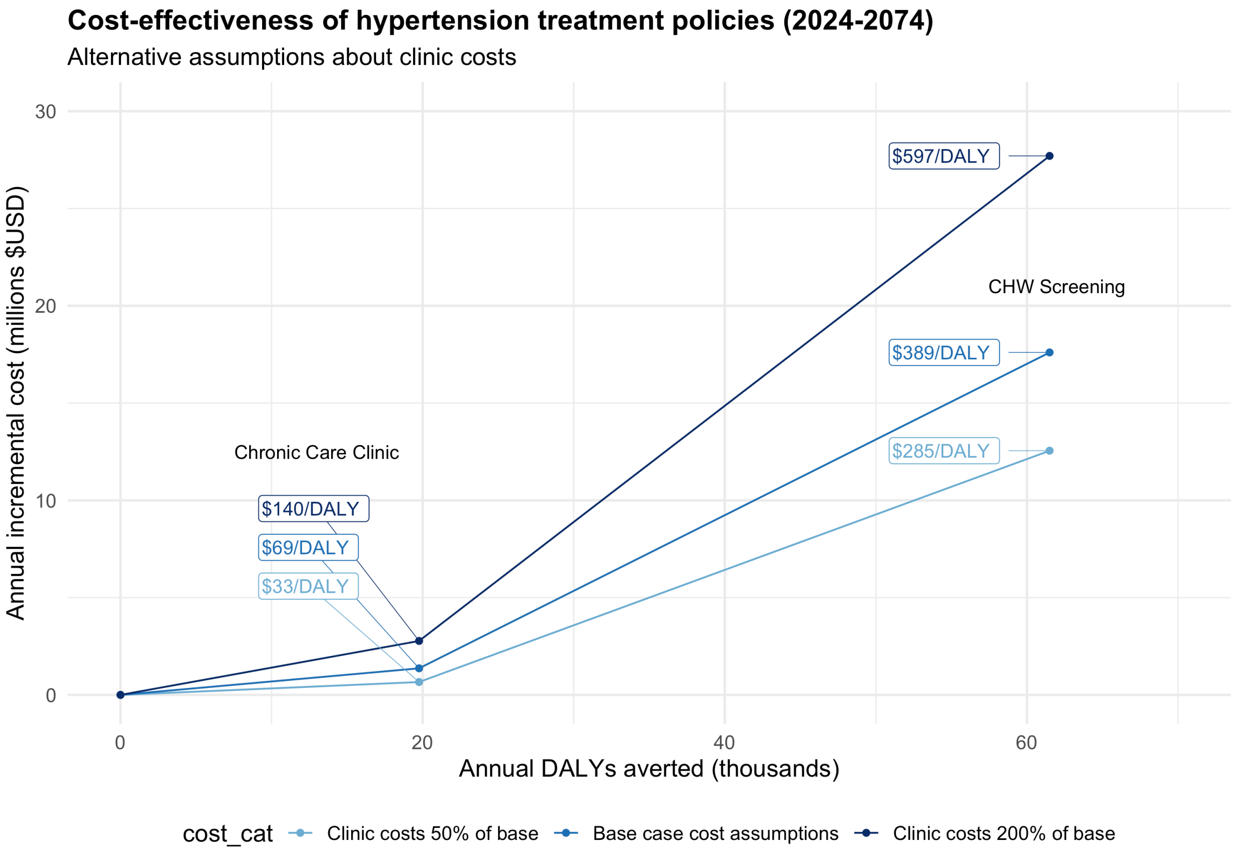

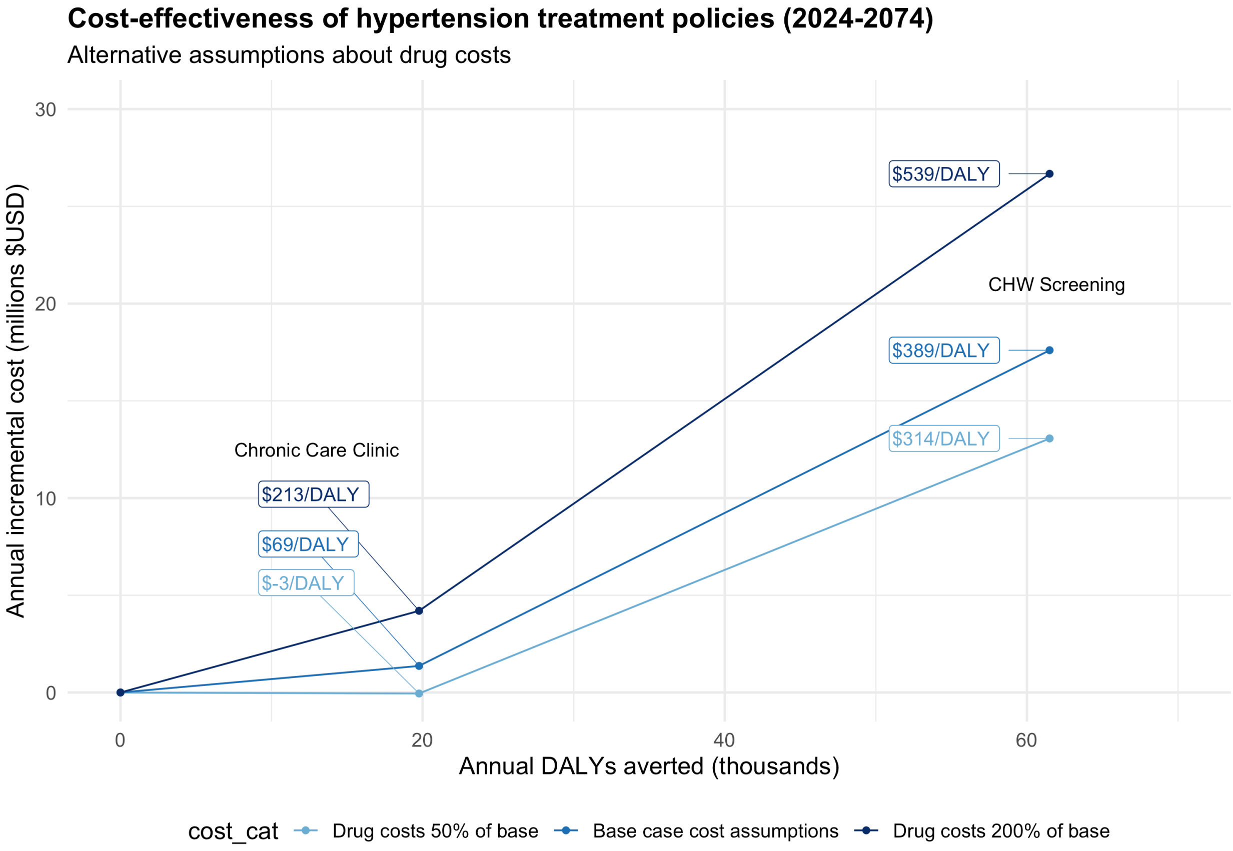

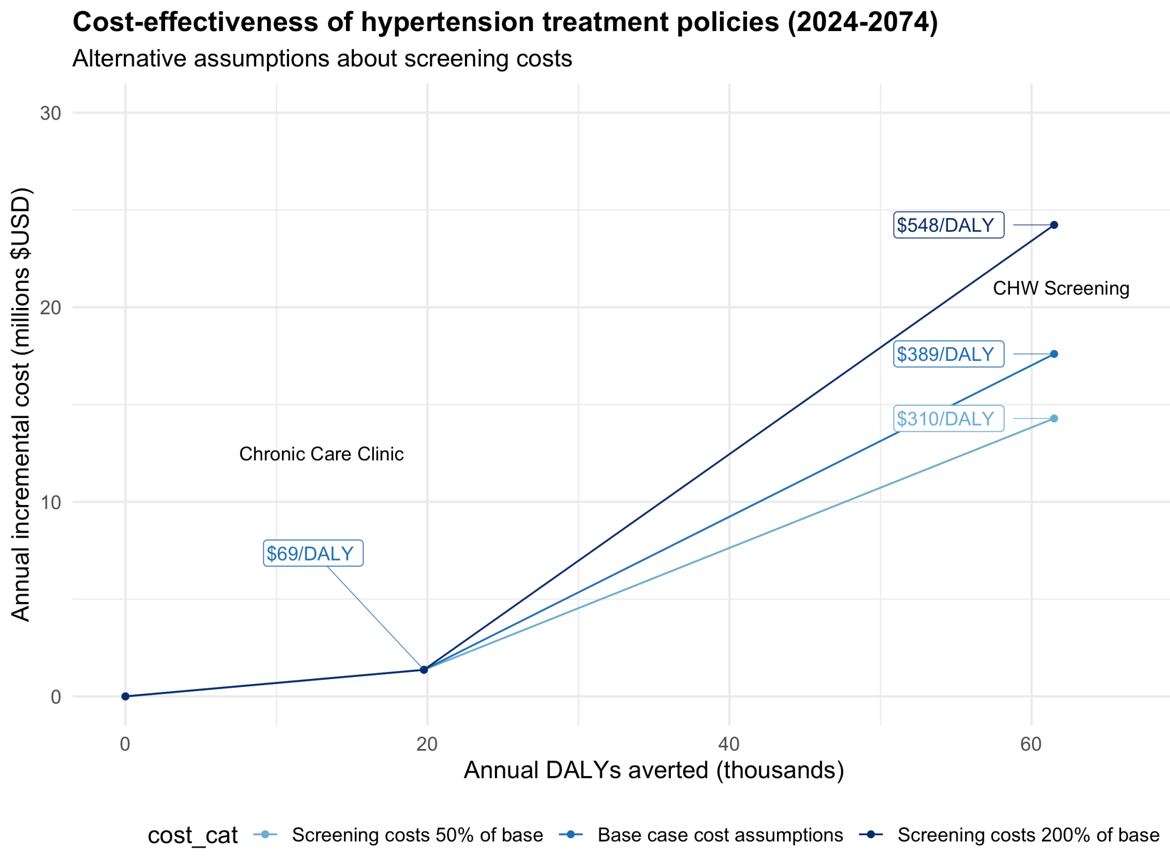

Cost-effectiveness of hypertension treatment policies (2024-2074)
Alternative assumptions about screening costs
$548/DALY
CHW Screening
$389/DALY
$310/DALY
Chronic Care Clinic
$69/DALY
Annual incremental cost (millions $USD)
30
20
10
0
0
20
40
60
Annual DALYs averted (thousands)
cost_cat
Screening costs 50% of base
Base case cost assumptions
Screening costs 200% of base

### Section 6. Five-Year Budget Impact Analysis

Tables in this section depict hypertension care cascade outcomes and undiscounted annual costs over the first five years of implementation (2024-2029).

##### Table S11. Hypertension care cascade outcomes of hypertension policy scenarios over 5 years (2024-2029)

| | Age | SOC | CCC | CHW |
|---|---|---|---|---|
| Hypertension prevalence* | ≥18 | 27% (19% to 35%) | 27% (19% to 35%) | 27% (19% to 35%) |
| | 25-44 | 43% (28% to 59%) | 43% (28% to 59%) | 43% (28% to 59%) |
| | 45-64 | 55% (41% to 67%) | 55% (41% to 67%) | 56% (41% to 67%) |
| | ≥65 | 22% (14% to 31%) | 24% (15% to 35%) | 43% (36% to 51%) |
| Diagnosis | ≥18 | 29% (19% to 41%) | 33% (20% to 48%) | 65% (57% to 73%) |
| | 25-44 | 41% (28% to 55%) | 43% (29% to 58%) | 71% (63% to 79%) |
| | 45-64 | 5% (2% to 9%) | 11% (5% to 20%) | 26% (20% to 33%) |
| | ≥65 | 7% (3% to 14%) | 17% (7% to 32%) | 42% (33% to 52%) |
| Overdiagnosis | ≥18 | 10% (4% to 18%) | 20% (9% to 34%) | 46% (36% to 55%) |
| | 25-44 | 3% (1% to 6%) | 8% (3% to 14%) | 18% (13% to 23%) |
| | 45-64 | 4% (1% to 8%) | 11% (4% to 23%) | 28% (21% to 38%) |
| | ≥65 | 5% (2% to 11%) | 12% (5% to 22%) | 29% (21% to 38%) |
| Current treatment | ≥18 | 27% (19% to 35%) | 27% (19% to 35%) | 27% (19% to 35%) |
| | 25-44 | 43% (28% to 59%) | 43% (28% to 59%) | 43% (28% to 59%) |
| | 45-64 | 55% (41% to 67%) | 55% (41% to 67%) | 56% (41% to 67%) |
| | ≥65 | 22% (14% to 31%) | 24% (15% to 35%) | 43% (36% to 51%) |

##### Table S12. Undiscounted costs of each policy over first 5 years (Budget Impact; 2024-2029)

| Annual undiscounted Costs (millions USD) | SOC | CCC | CHW |
|---|---|---|---|
| Screening | NA | NA | 7.7 (5.9 to 9.4) |
| Clinic | 4.8 (2 to 9.4) | 5.9 (2.6 to 10.6) | 14.9 (7.5 to 23.7) |
| Drug | 1.4 (0.5 to 2.9) | 3.4 (1.3 to 6.6) | 7.9 (4.4 to 12) |
| CVD care | 25.5 (5.9 to 64.3) | 23.5 (5.3 to 60.9) | 19.7 (4.6 to 49.7) |
| Total hypertension costs | 31.7 (10.5 to 71.8) | 32.8 (12.3 to 71.6) | 50.1 (27.1 to 86) |
| Incremental cost | NA | 1.1 (-4.7 to 5.8) | 18.4 (7.6 to 29.1) |

Scaled to population size of 10 million people aged ≥15 years

SOC = current standard of care, CCC = chronic care clinic, CHW = community health worker population-level screening (age ≥40). NA = not applicable. Data shown as means with 90% ranges. Population estimates among all adults persons 15+.

### Section 7. Sensitivity analyses

#### 7.1 CHW screening among age ≥50

We conducted a sensitivity analysis (n=1000 model runs), evaluating health effects and cost-effectiveness of increasing the screening age for CHW hypertension screening to 50 years and older (compared to 40 years and older in the primary analysis).

##### Table S13. Hypertension care cascade and cardiovascular disease outcomes of hypertension policy scenarios over 50 years (2024-2074) in sensitivity analysis with CHW screening beginning at age 50

| | Age | SOC | CCC | CHW (age ≥50) |
|---|---|---|---|---|
| Mean SBP | 25-44 | 125 (120 to 129) | 125 (120 to 129) | 125 (120 to 129) |
| | 45-64 | 138 (128 to 146) | 136 (127 to 144) | 133 (125 to 140) |
| | ≥65 | 144 (133 to 152) | 140 (131 to 149) | 134 (127 to 140) |
| Hypertension prevalence* | ≥18 | 30% (20% to 39%) | 30% (20% to 39%) | 30% (20% to 40%) |
| | 25-44 | 22% (15% to 29%) | 22% (15% to 29%) | 22% (15% to 29%) |
| | 45-64 | 45% (28% to 60%) | 45% (28% to 60%) | 45% (28% to 60%) |
| | ≥65 | 57% (38% to 70%) | 58% (39% to 71%) | 59% (39% to 73%) |
| Diagnosis | ≥18 | 23% (15% to 33%) | 27% (18% to 38%) | 46% (39% to 53%) |
| | 25-44 | 13% (7.6% to 19%) | 15% (8.5% to 22%) | 15% (8.6% to 22%) |
| | 45-64 | 28% (19% to 40%) | 34% (21% to 47%) | 61% (53% to 69%) |
| | ≥65 | 42% (29% to 57%) | 49% (33% to 65%) | 85% (79% to 92%) |
| Overdiagnosis | ≥18 | 0.4% (0.2% to 0.6%) | 0.7% (0.3% to 1.3%) | 2% (1.4% to 2.7%) |
| | 25-44 | 0.3% (0.1% to 0.4%) | 0.4% (0.2% to 0.7%) | 0.4% (0.2% to 0.7%) |
| | 45-64 | 0.8% (0.4% to 1.2%) | 1.5% (0.6% to 3.1%) | 4.9% (3.6% to 6.5%) |
| | ≥65 | 1.5% (0.8% to 2.5%) | 3.2% (1.2% to 6.7%) | 14% (11% to 18%) |
| Current treatment | ≥18 | 5.2% (2.2% to 9.6%) | 15% (7.4% to 25%) | 31% (24% to 38%) |
| | 25-44 | 2.1% (0.8% to 3.9%) | 6.6% (2.8% to 12%) | 6.6% (2.8% to 12%) |
| | 45-64 | 6.7% (2.8% to 12%) | 20% (9% to 33%) | 41% (32% to 50%) |
| | ≥65 | 10% (4.4% to 19%) | 29% (15% to 46%) | 63% (54% to 72%) |
| Ever treatment | ≥18 | 19% (12% to 29%) | 26% (16% to 36%) | 43% (35% to 50%) |
| | 25-44 | 9.2% (5.1% to 15%) | 13% (7.1% to 19%) | 13% (7.1% to 19%) |
| | 45-64 | 24% (15% to 35%) | 32% (19% to 45%) | 56% (48% to 65%) |
| | ≥65 | 38% (24% to 53%) | 47% (31% to 63%) | 83% (75% to 90%) |
| Over-treatment (proportion of normotensive individuals on treatment) | ≥18 | 0% (0% to 0.1%) | 0.3% (0.1% to 0.7%) | 0.9% (0.5% to 1.3%) |
| | 25-44 | 0% (0% to 0.1%) | 0.1% (0% to 0.3%) | 0.1% (0% to 0.3%) |
| | 45-64 | 0.1% (0% to 0.2%) | 0.8% (0.2% to 1.9%) | 2.1% (1.2% to 3.4%) |
| | ≥65 | 0.2% (0.1% to 0.4%) | 1.7% (0.4% to 4.2%) | 6.4% (3.9% to 9.7%) |
| Hypertension control | ≥18 | 2.8% (1% to 5.8%) | 11% (5% to 20%) | 23% (17% to 30%) |
| | 25-44 | 1.3% (0.5% to 2.7%) | 5.5% (2.2% to 10%) | 5.5% (2.2% to 10%) |
| | 45-64 | 3.5% (1.1% to 7.2%) | 14% (6% to 26%) | 30% (22% to 40%) |
| | ≥65 | 5.3% (1.7% to 11%) | 21% (9.3% to 35%) | 48% (38% to 58%) |
| IHD incidence (per 100 person-years)† | ≥18 | 0.25 (0.14 to 0.4) | 0.23 (0.13 to 0.36) | 0.19 (0.11 to 0.29) |
| | 25-44 | 0.04 (0.03 to 0.06) | 0.04 (0.03 to 0.06) | 0.04 (0.03 to 0.06) |
| | 45-64 | 0.34 (0.16 to 0.6) | 0.3 (0.15 to 0.5) | 0.24 (0.13 to 0.39) |
| | ≥65 | 1.63 (0.9 to 2.49) | 1.43 (0.82 to 2.18) | 1.11 (0.69 to 1.66) |
| Relative reduction in IHD incidence† | ≥18 | NA | 0.9 (0.83 to 0.97) | 0.75 (0.67 to 0.83) |
| | 25-44 | NA | 0.98 (0.86 to 1.11) | 0.98 (0.85 to 1.11) |
| | 45-64 | NA | 0.89 (0.79 to 0.98) | 0.72 (0.62 to 0.84) |
| | ≥65 | NA | 0.88 (0.78 to 0.96) | 0.69 (0.6 to 0.79) |
| CVA incidence (per 100 person-years)† | ≥18 | 0.27 (0.13 to 0.45) | 0.23 (0.11 to 0.4) | 0.18 (0.09 to 0.28) |
| | 25-44 | 0.02 (0.01 to 0.03) | 0.02 (0.01 to 0.03) | 0.02 (0.01 to 0.03) |
| | 45-64 | 0.34 (0.13 to 0.72) | 0.29 (0.11 to 0.57) | 0.21 (0.09 to 0.4) |
| | ≥65 | 1.91 (0.93 to 3.15) | 1.61 (0.83 to 2.61) | 1.18 (0.68 to 1.84) |
| Relative reduction in CVA incidence† | ≥18 | NA | 0.87 (0.77 to 0.95) | 0.67 (0.57 to 0.77) |
| | 25-44 | NA | 0.97 (0.8 to 1.15) | 0.97 (0.8 to 1.15) |
| | 45-64 | NA | 0.85 (0.73 to 0.95) | 0.63 (0.52 to 0.78) |
| | ≥65 | NA | 0.85 (0.74 to 0.94) | 0.63 (0.53 to 0.75) |
| CVD Mortality rate (per 100 person-years) | ≥18 | 0.44 (0.27 to 0.64) | 0.4 (0.25 to 0.6) | 0.34 (0.22 to 0.49) |
| | 25-44 | 0.06 (0.05 to 0.09) | 0.06 (0.05 to 0.08) | 0.06 (0.05 to 0.08) |
| | 45-64 | 0.6 (0.34 to 0.96) | 0.54 (0.31 to 0.86) | 0.45 (0.28 to 0.67) |
| | ≥65 | 2.81 (1.85 to 3.88) | 2.49 (1.66 to 3.53) | 2 (1.4 to 2.68) |
| Relative reduction of CVD mortality | ≥18 | NA | 0.91 (0.84 to 0.97) | 0.78 (0.7 to 0.85) |
| | 25-44 | NA | 0.98 (0.89 to 1.07) | 0.98 (0.89 to 1.08) |

| | | | | |
|---|---|---|---|---|
| | 45-64 | NA | 0.9 (0.81 to 0.97) | 0.76 (0.66 to 0.86) |
| | ≥65 | NA | 0.89 (0.8 to 0.96) | 0.72 (0.64 to 0.81) |
| CVD mortality rate (people with HIV; per 100 person-years) | ≥18 | 1.15 (0.52 to 2.38) | 0.83 (0.38 to 1.64) | 0.8 (0.36 to 1.58) |
| Relative risk of CVD mortality (people with HIV) | ≥18 | NA | 0.74 (0.59 to 0.89) | 0.71 (0.55 to 0.88) |
| All-cause mortality rate (per 100 person-years) | ≥18 | 1.36 (1.12 to 1.6) | 1.34 (1.1 to 1.57) | 1.31 (1.08 to 1.52) |
| Relative risk of all-cause mortality | ≥18 | NA | 0.98 (0.97 to 1) | 0.96 (0.93 to 0.98) |
| All-cause mortality rate (people with HIV; per 100 person-years) | ≥18 | 4.77 (3.02 to 8.58) | 4.59 (2.95 to 7.89) | 4.55 (2.91 to 7.76) |
| Relative risk reduction of all-cause mortality (people with HIV) | ≥18 | NA | 0.97 (0.86 to 1.09) | 0.96 (0.84 to 1.08) |

SOC = current standard of care, CCC = chronic care clinic, CHW = community health worker population-level screening (age ≥50), NA = not applicable. Data shown as means/proportions with 90% ranges

* True underlying current or pre-treatment SBP ≥140 mmHg (regardless of whether measured)

† Overdiagnosis indicates the proportion of normotensive individuals erroneously given a diagnosis of hypertension

‡ Overtreatment indicates the proportion of normotensive individuals diagnosed and treated for hypertension

§ Moderate/severe events only. Mild events assumed to be asymptomatic and therefore under-ascertained in existing data from the literature used for model calibration.

#### Table S14. Cost and cost-effectiveness of hypertension policies per 10 million adults over 50 years (2024-2074) in sensitivity analysis with CHW screening beginning at age 50

| | SOC | CCC | CHW (age ≥50) |
|---|---|---|---|
| DALYs Averted (thousands) | NA | 20.2 (-1.7 to 47.2) | 50.6 (14.5 to 92.3) |
| Costs (millions USD) | | | |
| Screening | NA | NA | 3.8 (3.2 to 4.4) |
| Clinic | 4 (1.5 to 7.8) | 5.5 (2.5 to 10.1) | 11.4 (6.3 to 17.2) |
| Drug | 1.2 (0.4 to 2.5) | 4.1 (1.6 to 7.4) | 8.1 (4.6 to 11.7) |
| CVD care | 23.5 (5.7 to 59.4) | 20.5 (5 to 52.4) | 15.9 (4.1 to 40.2) |
| Total HTN cost | 28.7 (9.3 to 64.5) | 30 (11.6 to 63.2) | 39.2 (21.5 to 66.9) |
| Incremental cost | NA | 1.3 (-3.6 to 4.8) | 10.5 (-1.1 to 19) |
| Incremental cost-effectiveness ratio (ICER; $USD / DALY averted)* | NA | $66 | $302 |
| Net DALYs averted (thousands)† | NA | 17.5 (-5 to 48) | 29.6 (-5.7 to 73.5) |
| Proportion of setting-scenarios where strategy is most cost-effective (considering all 3 policies)† | 4% | 23% | 72% |

Scaled to population size of 10 million people aged ≥15 years

SOC = current standard of care, CCC = chronic care clinic, CHW = community health worker population-level screening (age ≥40). NA = not applicable. Data shown as means with 90% ranges. Population estimates among all adults persons 15+.

* For the CHW policy, the ICER is calculated as the incremental cost-effectiveness of adding CHW screening to CCC (the next most effective policy in terms of health benefit).

† Assuming cost-effectiveness threshold of $500 USD per DALY averted. Net DALYs calculated by summing total average annual DALYs and average annual cost / cost effectiveness threshold of $500 USD.

#### Table S15. Proportion of setting-scenarios where each hypertension policy is the most cost-effective over next 50 years (2024-2074), assuming a cost-effectiveness threshold of $500 USD per DALY averted, in sensitivity analysis with CHW screening beginning at age 50

| Setting Scenario in 2023 | | SOC | CCC | CHW |
|---|---|---|---|---|
| Mean SBP among adults aged 45-64 | <132 | 16% | 51% | 34% |
| | 132 to <136 | 5% | 38% | 57% |
| | 136 to <140 | 2% | 16% | 82% |
| | >=140 | 0% | 6% | 94% |
| Hypertension prevalence among adults aged 45-64 | <35% | 15% | 46% | 40% |
| | 35 to <43% | 4% | 34% | 62% |
| | 43 to <50% | 2% | 18% | 80% |
| | >=50% | 0% | 6% | 94% |
| Hypertension diagnosis among adults aged 45-64 | <22% | 4% | 15% | 81% |
| | 22 to <28% | 5% | 18% | 77% |
| | 28 to <36% | 5% | 27% | 68% |
| | >=36% | 3% | 32% | 65% |
| Hypertension treatment among adults aged 45-64 | <4% | 6% | 17% | 78% |
| | 4 to <6% | 4% | 16% | 80% |
| | 6 to <9% | 6% | 27% | 68% |
| | >=9% | 2% | 32% | 66% |
| Hypertension control among adults aged 45-64 | <2% | 4% | 12% | 84% |
| | 2 to <4% | 4% | 19% | 77% |
| | 4 to <6% | 6% | 27% | 67% |
| | >=6% | 3% | 44% | 53% |
| CVD mortality rate | <400 per 100,000 | 10% | 42% | 47% |
| | 400 to <500 per 100,000 | 8% | 31% | 62% |
| | 500 to <600 per 100,000 | 1% | 23% | 76% |
| | >=600 per 100,000 | 0% | 8% | 92% |
| Population-level mean SBP rise per decade after 2015 | 0 mmHg | 4% | 21% | 75% |
| | 0.5 mmHg | 3% | 26% | 71% |
| | 1 mmHg | 6% | 23% | 71% |
| Proportion of acute CVD events receiving emergency care | 20% | 7% | 31% | 62% |
| | 40% | 5% | 24% | 72% |
| | 80% | 2% | 15% | 83% |
| Proportion of emergency CVD care that is effective* | 12.5% | 4% | 22% | 74% |
| | 25% | 4% | 24% | 72% |
| | 50% | 5% | 24% | 71% |
| Cost of ineffective emergency CVD care compared to effective care | 25% | 5% | 28% | 67% |
| | 50% | 3% | 24% | 72% |
| | 100% | 5% | 17% | 78% |
| HIV prevalence among adults aged 15-49 | <5% | 5% | 17% | 78% |
| | 5 to <10% | 5% | 21% | 74% |
| | 10 to <15% | 4% | 24% | 72% |
| | >=15% | 3% | 32% | 65% |
| Cost scenarios | Total costs 50% of base | 1% | 11% | 88% |
| | Acute CVD costs 25% of base | 8% | 33% | 60% |
| | Acute CVD costs 50% of base | 6% | 29% | 65% |
| | Drug costs 50% of base | 2% | 18% | 80% |
| | Clinic costs 50% of base | 2% | 16% | 82% |
| | Screening costs 50% of base | 3% | 17% | 81% |
| | Base case cost assumptions | 4% | 23% | 72% |
| | Screening costs 200% of base | 6% | 39% | 55% |
| | Clinic costs 200% of base | 11% | 43% | 45% |
| | Drug costs 200% of base | 12% | 35% | 53% |
| | Acute CVD costs 200% of base | 2% | 16% | 82% |
| | Acute CVD costs 400% of base | 1% | 10% | 89% |
| | Total costs 200% of base | 16% | 54% | 30% |

Table depicts the proportion of settings where a given policy is the most cost-effective (greatest number of net DALYs averted [DALYs averted – incremental costs / cost-effectiveness threshold], using a cost-effectiveness threshold of $500 USD/DALY averted). Row percentages total to 100%
SOC = current standard of care, CCC = chronic care clinic, CHW = community health worker population-level screening (age ≥50)
* Effective care represents emergency care that includes timely delivery of evidence-based interventions that result in acute mortality risk reduction

#### 7.2 Model estimates using a higher 5% discount rate for future costs and health benefits

We conducted a sensitivity analysis of the original 3000 model runs, evaluating cost effectiveness with a higher 5% annual discount rate for future health effects and costs. In the pr

##### Table S16. Cost and cost-effectiveness of hypertension policies per 10 million adults over 50 years (2024-2074) in sensitivity analysis with 5% discount rate

| | SOC | CCC | CHW |
|---|---|---|---|
| DALYs Averted (thousands) | NA | 11.9 (-1.8 to 27.7) | 36.5 (11.4 to 66) |
| Costs (millions USD) | | | |
| Screening | NA | NA | 4.5 (3.9 to 5.1) |
| Clinic | 2.7 (1.1 to 5.2) | 3.6 (1.6 to 6.5) | 9.4 (5.3 to 14.2) |
| Drug | 0.8 (0.3 to 1.7) | 2.7 (1.1 to 4.9) | 6.7 (3.9 to 9.7) |
| CVD care | 15.3 (3.7 to 38.8) | 13.4 (3.2 to 34) | 10 (2.5 to 24.9) |
| Total HTN cost | 18.8 (6.1 to 42.6) | 19.7 (7.8 to 40.9) | 30.7 (18 to 48.6) |
| Incremental cost | NA | 0.9 (-2 to 3.2) | 11.9 (4 to 18.7) |
| Incremental cost-effectiveness ratio (ICER; $USD / DALY averted)* | NA | $66 | $302 |
| Net DALYs averted (thousands)† | NA | 10.1 (-4.1 to 27.1) | 12.8 (-10.7 to 44.2) |
| Proportion of setting-scenarios where strategy is most cost-effective (considering all 3 policies)† | 9% | 40% | 51% |

Scaled to population size of 10 million people aged ≥15 years
SOC = current standard of care, CCC = chronic care clinic, CHW = community health worker population-level screening (age ≥40). NA = not applicable. Data shown as means with 90% ranges. Population estimates among all adults persons 15+.
* For the CHW policy, the ICER is calculated as the incremental cost-effectiveness of adding CHW screening to CCC (the next most effective policy in terms of health benefit).
† Assuming cost-effectiveness threshold of $500 USD per DALY averted. Net DALYs calculated by summing total average annual DALYs and average annual cost / cost effectiveness threshold of $500 USD.

#### Table S17. Proportion of setting-scenarios where each hypertension policy is the most cost-effective over next 50 years (2024-2074), assuming a cost-effectiveness threshold of $500 USD per DALY averted, in sensitivity analysis with 5% discount rate

| Setting Scenario in 2023 | | SOC | CCC | CHW |
|---|---|---|---|---|
| Mean SBP among adults aged 45-64 | <132 | 23% | 63% | 14% |
| | 132 to <136 | 13% | 57% | 30% |
| | 136 to <140 | 6% | 38% | 56% |
| | >=140 | 1% | 16% | 83% |
| Hypertension prevalence among adults aged 45-64 | <35% | 24% | 59% | 18% |
| | 35 to <43% | 10% | 56% | 34% |
| | 43 to <50% | 7% | 37% | 56% |
| | >=50% | 1% | 17% | 82% |
| Hypertension diagnosis among adults aged 45-64 | <22% | 11% | 30% | 59% |
| | 22 to <28% | 8% | 34% | 58% |
| | 28 to <36% | 10% | 41% | 49% |
| | >=36% | 8% | 54% | 39% |
| Hypertension treatment among adults aged 45-64 | <4% | 11% | 33% | 56% |
| | 4 to <6% | 10% | 33% | 57% |
| | 6 to <9% | 9% | 43% | 47% |
| | >=9% | 7% | 49% | 44% |
| Hypertension control among adults aged 45-64 | <2% | 9% | 27% | 64% |
| | 2 to <4% | 9% | 35% | 56% |
| | 4 to <6% | 9% | 50% | 41% |
| | >=6% | 9% | 58% | 33% |
| CVD mortality rate | <400 per 100,000 | 20% | 62% | 18% |
| | 400 to <500 per 100,000 | 14% | 47% | 39% |
| | 500 to <600 per 100,000 | 6% | 43% | 51% |
| | >=600 per 100,000 | 1% | 20% | 79% |
| Population-level mean SBP rise per decade after 2015 | 0 mmHg | 8% | 40% | 51% |
| | 0·5 mmHg | 10% | 42% | 48% |
| | 1 mmHg | 9% | 38% | 53% |
| Proportion of acute CVD events receiving emergency care | 20% | 12% | 48% | 40% |
| | 40% | 9% | 42% | 50% |
| | 80% | 6% | 31% | 63% |
| Proportion of emergency CVD care that is effective* | 12·5% | 10% | 41% | 49% |
| | 25% | 9% | 41% | 50% |
| | 50% | 8% | 38% | 54% |
| Cost of ineffective emergency CVD care compared to effective care | 25% | 11% | 44% | 45% |
| | 50% | 9% | 42% | 49% |
| | 100% | 8% | 35% | 58% |
| HIV prevalence among adults aged 15-49 | <5% | 11% | 34% | 56% |
| | 5 to <10% | 11% | 38% | 51% |
| | 10 to <15% | 6% | 45% | 49% |
| | >=15% | 7% | 47% | 45% |
| Cost scenarios | Total costs 50% of base | 2% | 12% | 86% |
| | Acute CVD costs 25% of base | 14% | 50% | 35% |
| | Acute CVD costs 50% of base | 12% | 47% | 41% |
| | Drug costs 50% of base | 5% | 30% | 65% |
| | Clinic costs 50% of base | 5% | 26% | 69% |
| | Screening costs 50% of base | 7% | 26% | 67% |
| | Base case cost assumptions | 9% | 40% | 51% |
| | Screening costs 200% of base | 12% | 62% | 26% |
| | Clinic costs 200% of base | 18% | 66% | 17% |
| | Drug costs 200% of base | 20% | 54% | 25% |
| | Acute CVD costs 200% of base | 5% | 29% | 66% |
| | Acute CVD costs 400% of base | 2% | 16% | 82% |
| | Total costs 200% of base | 22% | 69% | 9% |

Table depicts the proportion of settings where a given policy is the most cost-effective (greatest number of net DALYs averted [DALYs averted – incremental costs / cost-effectiveness threshold], using a cost-effectiveness threshold of $500 USD/DALY averted). Row percentages total to 100%
SOC = current standard of care, CCC = chronic care clinic, CHW = community health worker population-level screening (age ≥50)
* Effective care represents emergency care that includes timely delivery of evidence-based interventions that result in acute mortality risk reduction
